# Vibration of effects resulting from network geometry in mixed-treatment comparisons: a case study on network meta-analyses of antidepressants in major depressive disorder

**DOI:** 10.1101/2023.10.10.23296749

**Authors:** Constant Vinatier, Clement Palpacuer, Alexandre Scanff, Florian Naudet

## Abstract

**Objective:** It is frequent to find overlapping network meta-analyses (NMAs) on the same topic with differences in terms of both treatments included and effect estimates. We aimed to evaluate the impact on effect estimates of selecting different treatment combinations (i.e. network geometries) for inclusion in NMAs.

**Design:** Multiverse analysis, covering all possible NMAs on different combinations of treatments.

**Setting:** Data from a previously published NMA exploring the comparative effectiveness of 22 treatments (21 antidepressants and a placebo) for the treatment of acute major depressive disorder.

**Participants:** Cipriani et al (2018) explored a dataset of 116 477 patients included in 522 randomized controlled trials.

**Main outcome measures:** For each possible network geometry, we performed a NMA to estimate comparative effectiveness on treatment response and treatment discontinuation for the treatments included (231 between-treatment comparisons). The distribution of effect estimates of between-treatment comparisons across NMAs was computed, and the direction, magnitude, and statistical significance of the 1^st^ and 99^th^ percentiles were compared.

**Results:** 4 116 254 different NMAs concerned treatment response. Among possible network geometries, 172/ 231 (74%) pairwise comparisons exhibited opposite effects between the 1^st^ and 99^th^ percentiles, 57/231 (25%) comparisons exhibited statistically significant results in opposite directions, 118 of 231 (51%) comparisons derived non-robust results in terms of statistical significance at 5% risk and 56/231 (24%) treatment pairs obtained robust results across meta-analyses. Comparisons based on indirect evidence only were associated with greater variability in effect estimates. Comparisons with small absolute values observed in the complete NMA more frequently obtained statistically significant results in opposite directions. Similar results were observed for treatment discontinuation.

**Conclusion:** In this case study we observed that the selection of treatments to be included in a NMA could have considerable consequences on treatment effect estimations.

**Registration:** https://osf.io/mb5dy

## Introduction

Network meta-analyses (NMAs) are influential evidence synthesis tools often considered to dominate the hierarchy of evidence supporting clinical decision-making (1). By evaluating connected networks of RCTs, NMAs draw inferences on the comparative effectiveness of many interventions that may or may not have been compared directly. NMAs provide some answers to practical questions in day to day clinical practice, for instance which treatment should be prioritized when many treatments are available for the same condition (2). This information is all the more important in fields such as psychopharmacology where “blockbuster” drugs (e.g. fluoxetine for majors depressive disorders) co-exist with “me-too” drugs, marketed despite uncertain added value. Direct evidence for comparative effectiveness is indeed all too often lacking from regulatory approvals (3). For these reasons NMAs have become very popular tools in Evidence-Based Medicine.

NMAs are however victims of their own success, as their number is rapidly expanding with extensive overlap and potential redundancy. Too often, NMAs present an incomplete and fragmented picture of the total available evidence, with certain potential reproducibility issues. It has been observed that conclusions on comparative effectiveness can vary across overlapping NMAs on the same topic (4), suggesting that NMAs are prone to vibration of effects (VoE), which measures how far an effect estimate can vary across multiple distinct analyses (4). Several case studies have highlighted how VoE resulting from different methodological and analytical choices can lead to divergent and antagonistic conclusions in meta-analyses, e.g. for pairwise meta-analyses (5,6), for pooled analyses of individual patient data (7) and for indirect comparisons (6). Similar reproducibility issues are expected with NMAs since they rely on strong assumptions - e.g. transitivity (similarity across the studies included) and consistency (homogeneity between direct and indirect evidence) - which are quite difficult to ascertain (8). Because of the numerous interventions compared in NMAs, they are also prone to multiplicity issues (8). Even basic choices such as the consideration of eligible nodes to be included in a NMA can yield different effect estimates and treatment ranking (9). We aimed to quantify and visualize VoE arising from all possible network geometries i.e. all possible combination of treatments included in a network meta-analysis. For this purpose, we based our investigation on a widely known NMA by Cipriani et al. exploring the comparative effectiveness and acceptability of 21 antidepressant drugs and placebo for the treatment of adults with acute major depressive disorder (10).

## Methods

### Protocol, registration and reporting

The protocol was registered on August 3, 2020, on the Open Science Framework (OSF) before the start of the study (available at: https://osf.io/mb5dy). The results are presented according to the PRISMA checklist (Preferred Reporting Items for Systematic Reviews and Meta-Analysis) (11) and its extension for network meta-analyses (12).

### Data retrieval and study selection

We re-used the dataset used in Cipriani et al. NMA which is openly shared on Mendeley (available at: https://data.mendeley.com/datasets/83rthbp8ys/2). Data collection has been comprehensively detailed previously (10). Briefly, this dataset was collected up to Jan 8, 2016 and includes published and unpublished placebo-controlled and head-to-head double blind RCTs on 21 antidepressants (agomelatine, amitriptyline, bupropion, citalopram, clomipramine desvenlafaxine, duloxetine, escitalopram, fluoxetine, fluvoxamine, levomilnacipran, milnacipran, mirtazapine, nefazodone, paroxetine, reboxetine, sertraline, trazodone venlafaxine, vilazodone, and vortioxetine) used for the acute treatment of adults with major depressive disorder. Quasi-randomised trials, incomplete trials or trials that included 20% or more participants with bipolar disorder, psychotic depression, or treatment-resistant depression, or patients with a serious concomitant medical condition were not included. The dataset includes 522 RCTs involving 116 477 patients in 1199 different study arms, conducted between 1979 and 2016. All study arms evaluating the efficacy of antidepressants within the licensed dose range and the accepted/recommended dose range in the main clinical guidelines) (10) were considered.

### Study outcome

We explored VoE for the two different outcomes used in the NMA by Cipriani et al. The primary outcome was efficacy assessed using the response rate (treatment response defined by a reduction of ≥50% in the total score on a standardized observer-rated scale for depression). The secondary outcome was treatment discontinuation measured by the proportion of patients who withdrew for any reason. These outcomes were recorded as close to 8 weeks after initiation of treatment as possible and computed for all randomized patients. The response rate was imputed for 292 (24.3%) study arms, and dropouts were imputed as non-responders. In the case of multi-arm studies evaluating several doses of the same treatment for which the outcome was available, these arms were pooled.

### Assessment of vibration of effects

NMAs were performed for each possible network geometry derived from the 21 antidepressants and placebo (i.e. we constructed all possible networks with 2, 3, etc. up to 22 treatments). Among these possible networks, combinations that led to non-connected networks were excluded.

For all networks included, NMAs were performed. We collected network geometry (names of treatments, number of comparisons, number of participants treated), treatment comparisons (odds ratio and p-value) and two other metrics (Cochran’s Q and I² index). We computed the distribution of point estimates by effect sizes (ESs) and their corresponding p-values under the various analytical scenarios defined by the different network geometries. Comparisons were considered statistically significant if the ES was associated with a p-value < 0.05. For each comparison pair, the presence of a “Janus effect” was investigated by calculating the 1st and 99th percentiles of the distribution of the ES (13). A Janus effect is defined as an ES that is in the opposite direction between the 1st and 99th percentiles of the meta-analysis. It demonstrates the presence of substantial VoE. In addition, we computed the distribution of the I^2^ indices, and the p-values on Cochran’s Q test calculated for each network meta-analysis. Heterogeneity was considered statistically significant if the p-value for the Q test was < 0.10.

The network meta-analyses were performed with R software [version 4.2.2 (2022-10-31)] (14) netmeta package, which uses a frequentist method to perform NMAs (15), the doParallel package (16) and the tydiverse language (17). A random-effect model was considered for all NMAs.

### Changes to the initial protocol

In addition to the Janus effect, we described two additional parameters in order to have a more comprehensive understanding of VoE in this dataset: 1/ an extreme form of the Janus effect where the two extremes exhibit statistically significant results and 2/ the relative Odds Ratios (ROR) as described by Patel et al. (13), calculated as the ratio of ORs at the 1^st^ and 99^th^ percentile, which enables quantification of variations in point estimates, even when no Janus effect is observed. A ROR value of 1 suggests the absence of VoE, whereas higher ROR values indicate a more pronounced level of vibration (18). We explored the correlation between the ROR for treatment response and the ROR for treatment discontinuation using Spearman’s rank correlation.

As an exploratory analysis using our assessment of VoE for all treatment comparisons, we decided to investigate, using either a logistic or a linear model, the associations for 1/ the Janus Effect, 2/ the existence of statistically significant results in opposite directions and, 3/ the RORs with the following explanatory variables considered as possible sources of VoE in NMAs: 1/ a categorial variable describing the type of available evidence for the comparison in the full network (presence of direct comparisons without inconsistency, presence of direct comparisons with inconsistency, indirect comparisons only). A threshold for the p-value <0.10 was used to define inconsistency, from a two-sided z test comparing direct and indirect evidence determined on the most complete network(15)) and 2/ the effect size of the comparison in question (absolute value of the log Odds Ratio of the most complete meta-analysis). With this last parameter we aimed to explore whether null results were more likely to induce a Janus effect. Because of the lack of normality of residuals in the linear model for ROR, a log transformation was applied.

### Patient and public involvement

Patients and the public were not involved in the design, conduct, reporting, or dissemination plans for this research. This was a methodological study, and we had no established contacts with specific patient groups who might be involved in this project.

## Results

### Primary outcome: treatment response

Among the 4 194 281 possible NMAs, 78 027 (2%) non-connected networks were excluded, resulting in a total of 4 116 254 NMAs (see **e-Table 1**). The percentage of non-connected networks decreased as the number of treatments per network increased, falling from 57% for networks of 2 treatments to 0% for networks with 18 to 22 treatments. **Figure 1** and **e-Table 2** summarize the distribution of the network geometries observed for all 4 116 254 NMAs included. All treatments except milnacipran had direct comparisons with placebo which was the most widely represented arm (with 35 721 patients). The most frequent direct comparisons were those for paroxetine versus placebo (46 studies) and fluoxetine versus placebo (40 studies). Levomilnacipran was the only treatment represented in the network by a single comparison (versus placebo). Among the 231 pairs across the 22 treatments, 99 had direct evidence and 132 relied only on indirect evidence.

**Table 1:** Association between vibration of effects indices and various characteristics of treatment comparisons. † effect sizes are expressed as absolute values of the log odds-ratio estimated in the most complete network meta-analysis

| Characteristics of treatment comparisons | Janus effect |  | Statistically significant results in opposite directions |  | Log(Relative Odds Ratio) |  |
| --- | --- | --- | --- | --- | --- | --- |
| | OR [95% CI] | p-value | OR [95% CI] | p-value | $\beta$ [95% CI] | p-value |
| **Treatment response** |  |  |  |  |  |  |
| Type of available evidence for the comparison (ref = direct evidence without inconsistency) |  |  |  |  |  |  |
| - Direct evidence with inconsistency | 0.393 [0.149 ; 1.016] | 0.055 | 0.596 [0.126 ; 2.117] | 0.458 | 0.041 [-0.109 ; 0.190] | 0.593 |
| - Only indirect evidence | 3.176 [1.587 ; 6.492] | 0.001* | 2.225 [1.109 ; 4.699] | 0.029* | 0.406 [0.313 ; 0.498] | <0.001* |
| Effect Size† | 0.037 [0.004 ; 0.373] | 0.005* | 0.004 [0.000 ; 0.092] | 0.002* | -0.072 [0.396 ; 0.252] | 0.660 |
| **Treatment discontinuation** |  |  |  |  |  |  |
| Type of available evidence for the comparison (ref = direct evidence without inconsistency) |  |  |  |  |  |  |
| - Direct evidence with inconsistency | 0.643 [0.221 ; 1.943] | 0.4217 | 0.906 [0.128 ; 4.113] | 0.907 | 1.134 [0.941 ; 1.366] | 0.184 |
| - Only indirect evidence | 2.773 [1.281 ; 6.141] | 0.0103* | 3.641 [1.600 ; 9.433] | 0.004* | 1.632 [1.456 ; 1.828] | <0.001* |
| Effect Size† | $3.25 \cdot 10^{-6}$ [$2.68 \cdot 10^{-8}$ ; $2.56 \cdot 10^{-4}$] | <0.001* | 0.063 [0.001 ; 4.329] | 0.214 | 0.667 [0.350 ; 1.272] | 0.217 |

**Figure 1:**
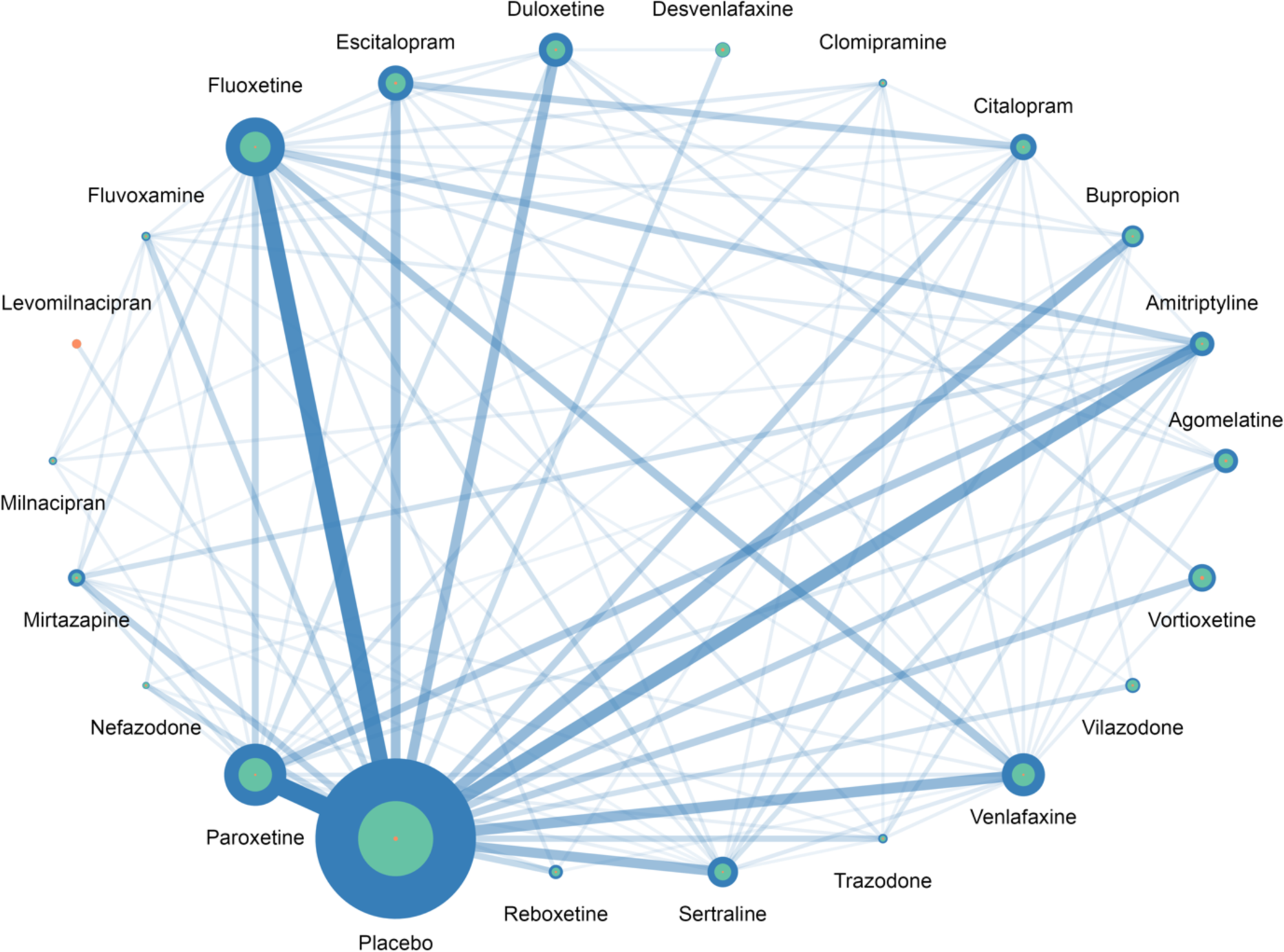
Distribution of network geometry for NMAs on treatment response. The size of each dot represents the number of patients allocated to the respective treatments. For each treatment, the blue circles indicate the NMAs with the largest number of patients included, the green circles represent the NMAs with the median number of patients included, and the orange circles show the NMAs with the smallest number of patients included s. The width of the lines is proportional to the number of trials comparing pairs of treatments in the complete NMA.

**Figure 2a** summarizes VoE observed across the 231 treatments comparisons. After computing the 4 116 254 NMAs, we observed the presence of the Janus effect in 172/231 (74%) treatment comparisons. There were statistically significant results pointing in opposite directions for 57/231 (25%) of the comparisons. 56/231 (24%) comparisons were robust in identifying either a statistically significant difference or no difference across NMAs and 118/231 (51%) comparisons had non-robust results for the existence or absence of statistical significance at 5% risk across NMAs. RORs ranged from 1.01 to 5.96 with a median ROR of 1.72 (interquartile range, IQR: 1.03-4.83).

**Figure 2:**
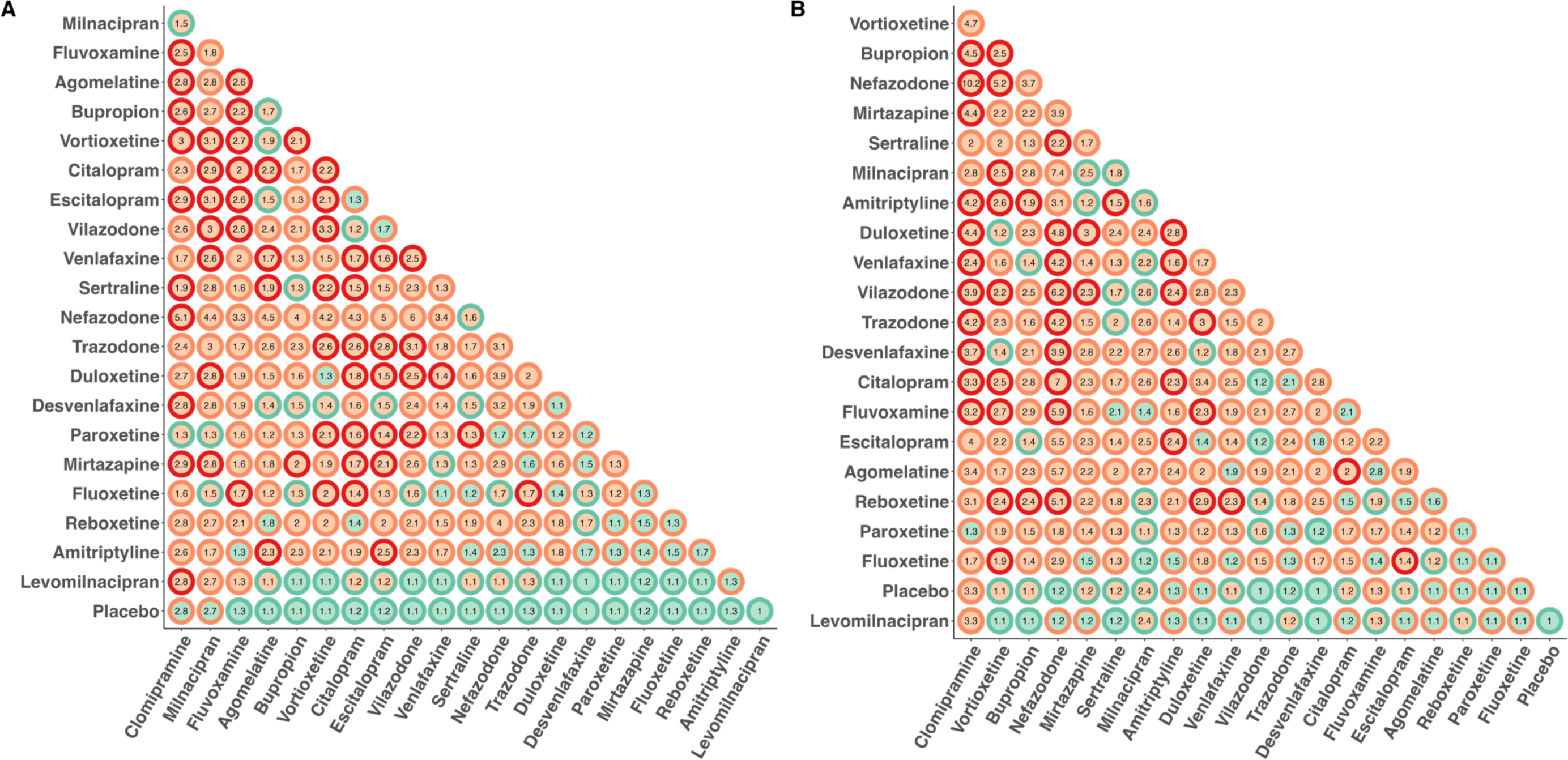
VoE in the 231 comparisons across the 22 treatments, classified according to their degree of VoE. Panel A: For treatment response. Panel B: For treatment discontinuation. For each dot, the center indicates the existence of a Janus Effect (green=no, orange=yes), the outline indicates the existence of statistically significant results in two opposite directions (green= robust identification of either a statistically significant difference or no difference, orange: non-robust results for the existence or absence of statistical significance, red: significant results observed in opposite directions). Numbers correspond to the Relative Odd Ratios.

Clomipramine (**Figure 3**) was the treatment with the highest level of VoE with a Janus effect present in all comparisons except the comparison with placebo. NMAs showing statistically significant results in opposite directions were present for 10 different comparisons.

**Figure 3:**
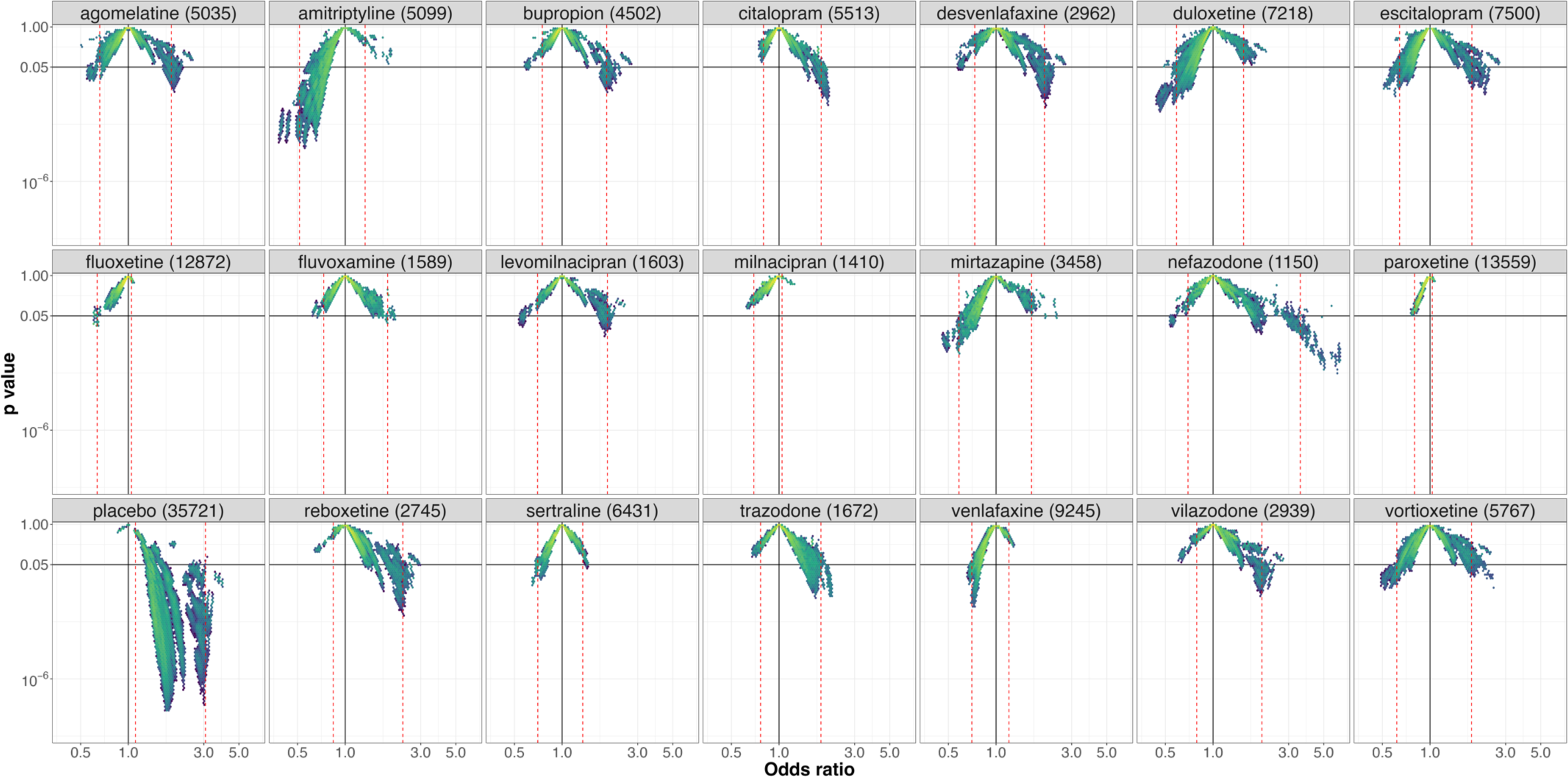
Vibration of effects for treatment response for the comparisons of clomipramine with the 20 remaining antidepressants and placebo (with the number of patients included in the most complete network for this comparison). An Odd ratio >1 favors clomipramine. The colors indicate the log densities of network meta-analyses (yellow: high, green: moderate, blue: low). Dotted red lines show the 1^st^ and 99^th^ percentiles.

Placebo (**Figure 4**) was the treatment with the lowest level of VoE. No Janus effect was identified for any comparisons. All NMAs identified a statistically significant superiority of antidepressants over placebo, except for clomipramine and milnacipran for which 16% and 11% of the NMAs respectively failed to identify statistically significant results.

**Figure 4:**
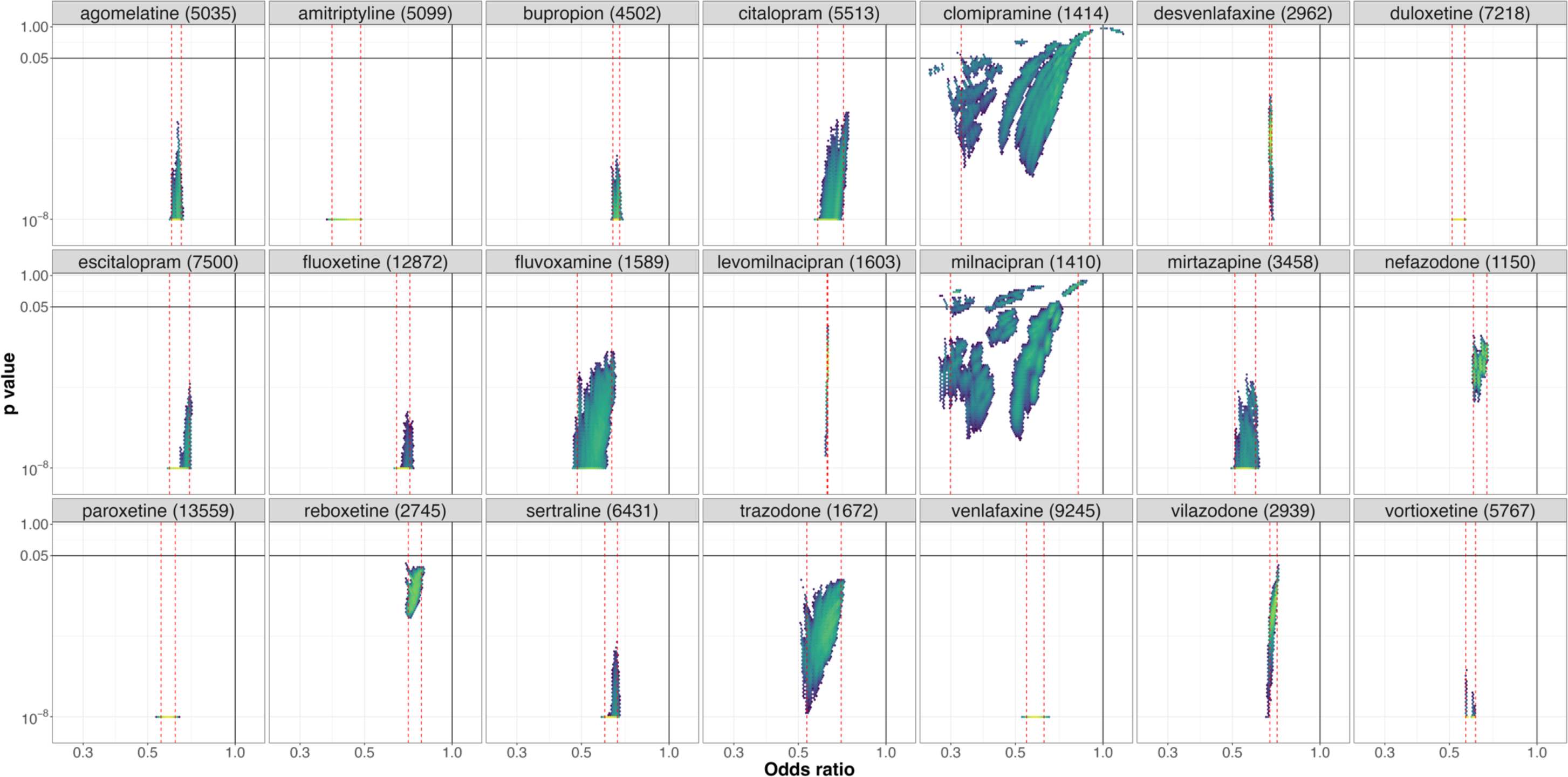
Vibration of effects for treatment response for the comparisons of placebo with the 21 antidepressants (with the number of patients included in the most complete network for this comparison). An Odds ratio >1 favors placebo. The colors indicate the log densities of network meta-analyses (yellow: high, green: moderate, blue: low). Dotted red lines show the 1^st^ and 99^th^ percentiles.

Results for other treatments are presented in supplementary **e-Figures** 1-20.

Across all NMAs assessing treatment response, the median I^2^ was 31% (IQR = 22–36%) and the p-value for Cochran’s Q test was < 0.10 for 3 353 881/4 116 254 (81%) of the NMAs. **e-Figure 21** details vibration for these 2 parameters.

### Secondary outcome: treatment discontinuation

For these analyses we had to exclude 6 studies corresponding to 145 patients randomized in 7 arms because of absence of event. Of the 4 194 281 possible NMAs, 72 691 (2%) non-connected networks were not included, resulting in a total of 4 121 590 NMAs (see **e-Table 1**) i.e. 5 336 more than for treatment response. **E-Figure 22 and e-Table 2** summarize the distribution of the network geometries observed for the 4 121 590 NMAs included. Four direct comparisons that were available for treatment response were missing for treatment discontinuation (clomipramine vs milnacipran, clomipramine vs trazodone, fluoxetine vs vilazodone and reboxetine vs venlafaxine). Conversely, there were two direct comparisons for treatment discontinuation that were absent for treatment response (amitriptyline vs bupropion and clomipramine vs placebo). Among the 231 comparisons of the 22 treatments, 97 had direct evidence and 134 relied only on indirect evidence. **Figure 2b** summarizes VoE observed across the 231 treatment comparisons. After computing the 4 194 281 NMAs, we observed a Janus effect in 180/231 (74%) treatment comparisons. We also observed statistically significant results pointing in opposite directions for 45/231 (19%) of the comparisons. 46/231 (20%) of the comparisons were able to identify a robust or statistically significant difference or absence of difference, and 140/231 (61%) comparisons had non-robust results for the existence or absence of statistical significance. RORs ranged from 1.01 to 10.17 with a median ROR of 1.95 (interquartile range, IQR: 1.33-2.50). Results observed for all treatments are detailed in **e-Figures 23-44**. Among the NMAs assessing treatment discontinuation, the median I^2^ was 24% (IQR = 11–30%) and the p-value on Cochran’s Q test was < 0.10 for 2 539 033/4 121 590 (61%) of the NMAs. **e-Figure 45** details vibration for these 2 parameters. RORs observed for treatment discontinuation were correlated with RORs observed for treatment response (Spearman’s ρ=0.86, p value <0.001, **Figure 5**).

**Figure 5:**
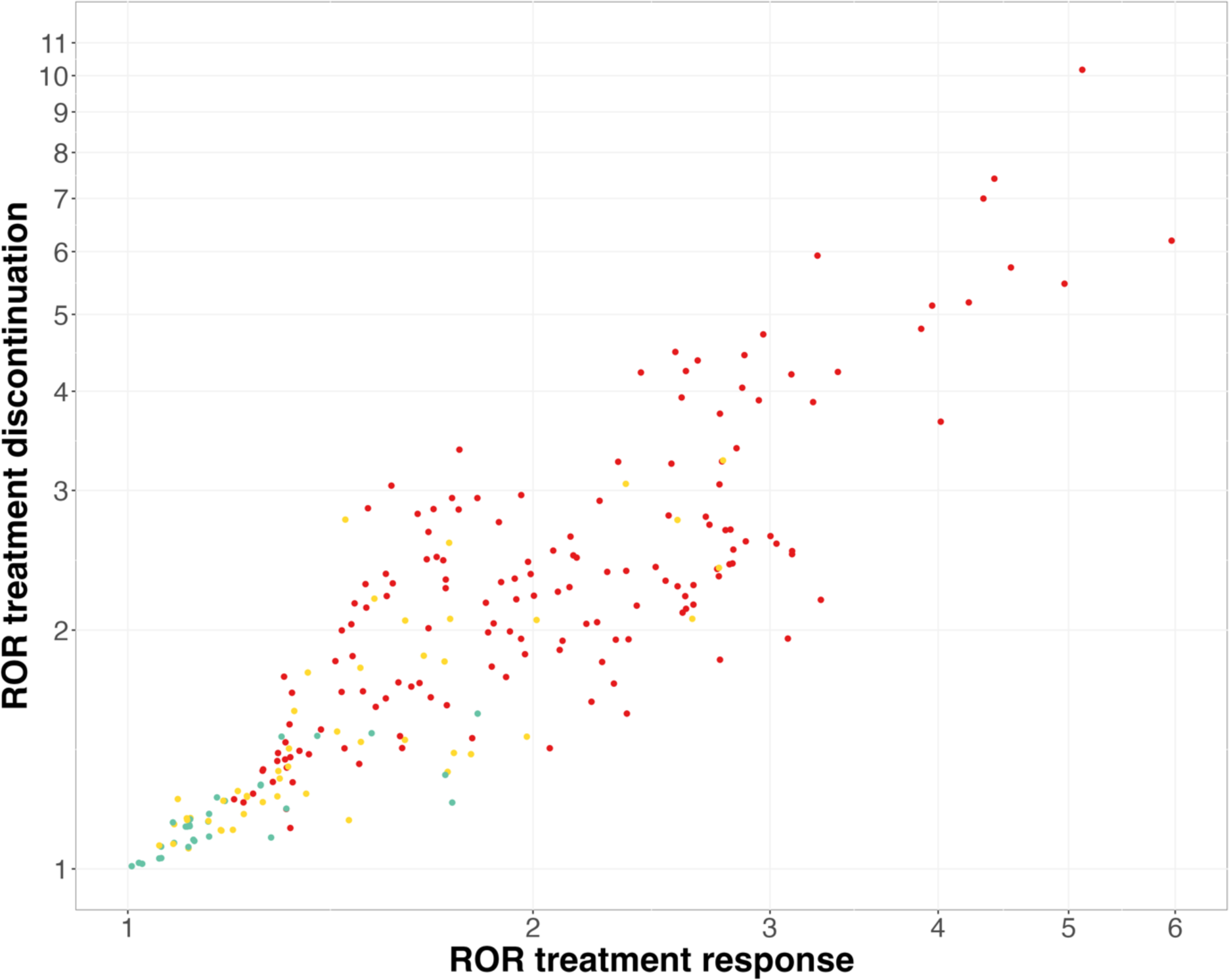
RORs for treatment response and treatment discontinuation. The color of the dots indicates the presence of a Janus effect for both outcomes (green: no Janus effect, yellow: Janus effect for one of the outcomes and red: Janus effect for both).

### Exploratory Analysis of characteristics associated with VoE

Levomilnacipran was only studied against placebo, making it impossible to provide indirect evidence, which is why this comparison was left out for the exploratory analysis. The results based on the remaining 230 comparisons are presented in **Table 1**. Regarding treatment response, indirect evidence was associated with a more frequent Janus effect, more results in opposite directions and greater RORs, while the effect size observed in the most comprehensive meta-analysis (expressed as an absolute value of the log odds-ratios) was only found to be associated with the Janus effect and statistically significant results in opposite directions. Quite similar results were observed for treatment discontinuation.

## Discussion

### Statement of principal findings

In this case study we performed 4 116 254 NMAs evaluating the comparative treatment response of 21 antidepressants and placebo. Depending on network geometry, we identified substantial VoE with the presence of a Janus effect in 172/231 (74%) comparisons. For 57/231 comparisons (27%) VoE yielded statistically significant results in opposite directions. Similar results were observed among the 4 121 590 NMAs evaluating treatment discontinuation. RORs for treatment response and treatment discontinuation were highly correlated. Comparisons relying on indirect evidence alone were associated with all three indices of VoE (Janus effect, significant results in opposite directions and RORs). Having an effect size close to zero (as assessed in the most comprehensive meta-analysis), was associated with the Janus effect, with significant results in opposite directions, but not with RORs.

In other terms, variations in estimated effects are greater for comparisons relying on indirect evidence only. When the actual differences between treatments are small, this can lead to effect estimates in opposite directions, and occasionally to statistically significant results in both directions. It is not surprising to see these results in this very specific case study focused on antidepressants. Many of the drugs studied are me-too drugs from a few therapeutic classes, resulting in small difference between treatments. In addition, VoE could be expected in this corpus, as a previous re-analysis of the Cipriani et al. dataset was able to identify differences among antidepressant placebos although all are composed of sucrose (19), to some extent suggesting violations of the main assumption of NMAs.

### Strengths and weaknesses of the study

We used a well-known NMA with 22 different treatments (including placebo), making it possible to study a large number of network geometries. As this was a case study, different results could be observed in a different context, e.g. for networks of different size or in different fields. In addition, estimating VoE related to network geometry could be difficult to conduct for NMAs exploring smaller networks of RCTs. Smaller networks could be less prone to VoE because of the network geometry, as the contribution of indirect comparisons is associated with the number of treatments included in the NMA (20). On the other hand, studies of VoE related to network geometry can be challenging in larger networks, since performing a large number of NMAs requires a lot of computing time. It took us almost 3 months on a personal computer to compute the near 8 million NMAs needed for this specific case-study (around 4 million for treatment response and same number for treatment discontinuation).

In addition, we considered only network geometry as a source of VoE for this study. Although it seems to be a relevant choice, as differences in network geometry are frequently observed for overlapping NMAs on the same topic(4), complementary methodological choices could have been made, for example the exhaustiveness of the evidence base (related both to the selection criteria and to the quality of the literature searches) or the risk of bias in the RCTs included. In addition, for network geometry, additional VoE could be related to decisions made to merge or not to merge different doses of the same treatment in a given node.

### Strengths and weaknesses in relation to other studies, discussion of important differences in results

After our previous case study, which made several methodological choices for indirect comparison meta-analyses to compare nalmefene and naltrexone in the reduction of alcohol consumption (6), this new study, in a more complex network, corroborates VoE arising from indirect comparisons. VoE was also found to influence the results in a head to head meta-analysis in the case of acupuncture for smoking cessation, a domain that is known for its clinical and methodological heterogeneity (21). Similarly, marked VoE was observed in a meta-analysis comparing operative with nonoperative treatments for proximal humerus fractures (5). While the domain of antidepressant research is probably more standardized with less variability in interventions and study designs than acupuncture or surgery, we were still able to evidence VoE. In addition, VoE has been observed in pooled analyses of individual participant data from 12 randomized controlled trials comparing canagliflozin and placebo for type 2 diabetes mellitus (7). All these case studies were useful to investigate reproducibility issues and controversies arising from redundant and overlapping meta-analyses (22). Nevertheless, these studies converge to point to the existence of VoE in meta-analyses, and we recommend further research to systematically explore VoE and its determinants (e.g. effect sizes, heterogeneity, inconsistency, risk of bias in studies included and random sampling) in a large set of meta-analyses before any systematic implementation in routine practice.

### Meaning of the study: possible explanations and implications for clinicians and policymakers

Our results show that effect estimates in NMAs can be impacted by the network structure. In other words, NMAs allow for a certain amount of analytical flexibility, which can lead to divergent results, and NMAs can therefore be easily hijacked to a desired conclusion. This is all the more important since NMAs have particular importance for clinical decision-making: since direct evidence of comparative effectiveness is all too often lacking in regulatory approvals (23), indirect evidence is often required for guideline development (24). Concerning the conduct of NMAs, analytical flexibility can be partly defined by pre-registration in Prospero (25), a practice that is encouraged but not enforced by most journals, as there is no policy for meta-analyses similar to the 2005 ICMJE policy on clinical trials (26). Still, because meta-analyses are almost always retrospective studies that gather existing evidence, the possibility of an a posteriori registration is often very difficult to rule out. The constitution of systematic, permanent, living NMAs could also help to reduce reporting bias of this sort. Regarding interpretation of NMAs, our results highlight the importance of considering uncertainties in NMA results, and corroborate the widespread idea that indirect comparisons can lead to biased conclusions (27,28). This is all the more important since empirical evidence suggests that in NMAs, most of the information often comes from indirect evidence (20). It also raises doubts about the relevance of indirect comparisons as a decision-making tool, and provides empirical support for the GRADE approach for NMAs, which considers the certainty of evidence for all direct, indirect and NMA estimates between interventions included in the network (29), and downgrades certainty of the evidence in case of absence of direct comparisons.

### Unanswered questions and future research

In this case study, we explored the VoE arising from the network geometry in a large NMA on 21 antidepressants and placebo in the treatment of major depressive disorders. We found substantial variations in the magnitude, direction and statistical significance of the effects estimated. These findings suggest that when conducting NMAs on randomized controlled trials, the selection of treatments to be included in the network could have considerable consequences on treatment effect estimations. More comprehensive studies on VoE across the medical literature are needed to gain better understanding of these reproducibility issues and to define safeguards to limit their impact on clinical decision-making.

## Data Availability

All data produced are available online athttps://osf.io/hb7uj/

## Acknowledgements

We thank Angela Verdier for revising the English, and Karima Hammas for her help for the protocol.

## Contributions

CP initiated and designed the study. FN initiated and designed the study, interpreted the results and drafted the manuscript. CV cleaned the data, performed the analysis, interpreted the results and drafted the manuscript. AS contributed to the data analysis. All authors have critically revised the manuscript for important intellectual content and approved the manuscript.

FN acts as guarantor. The corresponding author attests that all listed authors meet authorship criteria and that no others meeting the criteria have been omitted.

## Ethics approval

Not applicable.

## Data-sharing statement

The data and code are openly shared on the OSF (https://osf.io/hb7uj/) and the dataset is shared by the authors on Mendeley (https://data.mendeley.com/datasets/83rthbp8ys/2).

## Dissemination plans

There is at present no specific dissemination plan for these results.

## Transparency

The lead author and the manuscript guarantor state that the manuscript is an honest, accurate, and transparent account of the study reported, that no important aspects of the study have been omitted, and that any divergences from the study as planned have been explaine

