## Supplementary material for "Vibration of effects resulting from network geometry in mixed-treatment comparisons: a case study on network meta-analyses of antidepressants in major depressive disorder": Suplementaries

### Appendices

| Number of treatments in the networks | Theoretical number of possible NMAs | Non-connected networks n (%) | |
|---|---|---|---|
| | | *Treatment response* | *Treatment discontinuation* |
| ***2*** | 231 | 132 (57.1) | 97 (58.0) |
| ***3*** | 1 540 | 414 (26.9) | 435 (28.3) |
| ***4*** | 7 315 | 924 (12.6) | 980 (13.4) |
| ***5*** | 26 334 | 2 030 (7.71) | 2030 (7.71) |
| ***6*** | 74 613 | 4 529 (6.07) | 4333 (5.81) |
| ***7*** | 170 544 | 8 995 (5.27) | 8284 (4.86) |
| ***8*** | 319 770 | 13 779 (4.31) | 12 555 (3.93) |
| ***9*** | 497 420 | 15 946 (3.20) | 14 590 (2.93) |
| ***10*** | 646 646 | 14 068 (2.18) | 13 014 (2.01) |
| ***11*** | 705 432 | 9 548 (1.35) | 8 955 (1.27) |
| ***12*** | 646 646 | 4 990 (0.77) | 4 795 (0.73) |
| ***13*** | 497 420 | 1987 (0.40) | 1 919 (0.39) |
| ***14*** | 319 770 | 585 (0.18) | 575 (0.18) |
| ***15*** | 170 544 | 122 (0.07) | 121 (0.07) |
| ***16*** | 74 613 | 16 (0.02) | 16 (0.02) |
| ***17*** | 26 334 | 1 (0.01) | 1 (0.01) |
| ***18*** | 7 315 | 0 (0) | 0 (0) |
| ***19*** | 1 540 | 0 (0) | 0 (0) |
| ***20*** | 231 | 0 (0) | 0 (0) |
| ***21*** | 22 | 0 (0) | 0 (0) |
| ***22*** | 1 | 0 (0) | 0 (0) |

***e-Table 1:*** *Number of possible NMAs (combinations of 22 treatments) and non-connected networks depending on the number of treatments per network.*

| | **Treatment response** | | | | **Treatment discontinuation** | | | |
|---|---|---|---|---|---|---|---|---|
| | **Number of patients included in NMAs** | | | **Number of direct comparisons** | **Number of patients included in NMAs** | | | **Number of direct comparisons** |
| | ***min*** | ***max*** | ***median*** | | ***min*** | ***max*** | ***median*** | |
| ***Agomelatine*** | 202 | 5 035 | 2 905 | 6 | 202 | 4 153 | 2 848 | 6 |
| ***Amitriptyline*** | 51 | 5 099 | 2 641 | 11 | 41 | 4 270 | 2 465 | 12 |
| ***Bupropion*** | 63 | 4 502 | 2 954 | 7 | 63 | 3 363 | 2 808 | 8 |
| ***Citalopram*** | 57 | 5 513 | 2 795 | 12 | 25 | 4 013 | 2 577 | 12 |
| ***Clomipramine*** | 41 | 1 414 | 876 | 8 | 20 | 1 201 | 799 | 7 |
| ***Desvenlafaxine*** | 315 | 2 962 | 2 647 | 2 | 315 | 2 647 | 2 647 | 2 |
| ***Duloxetine*** | 152 | 7 218 | 3 855 | 8 | 152 | 4 557 | 3 633 | 8 |
| ***Escitalopram*** | 227 | 7 500 | 4 174 | 9 | 227 | 5 726 | 4 082 | 9 |
| ***Fluoxetine*** | 70 | 12 872 | 6 580 | 18 | 70 | 9 443 | 6 178 | 17 |
| ***Fluvoxamine*** | 34 | 1 589 | 914 | 10 | 20 | 1 708 | 1 016 | 10 |
| ***Levomilnacipran*** | 1 603 | 1 603 | 1 603 | 1 | 1 693 | 1 693 | 1 693 | 1 |
| ***Milnacipran*** | 27 | 1 410 | 832 | 6 | 27 | 1 358 | 780 | 5 |
| ***Mirtazapine*** | 137 | 3 458 | 1813 | 9 | 137 | 2 974 | 1 769 | 9 |
| ***Nefazodone*** | 55 | 1 150 | 887 | 5 | 55 | 1 226 | 963 | 5 |
| ***Paroxetine*** | 85 | 13 559 | 7328 | 17 | 85 | 11 329 | 7 484 | 17 |
| ***Placebo*** | 553 | 35 721 | 17 046 | 19 | 18 | 27 225 | 16 371 | 20 |
| ***Reboxetine*** | 80 | 2 745 | 1 478 | 5 | 229 | 1 866 | 1 478 | 4 |
| ***Sertraline*** | 26 | 6 431 | 3 414 | 14 | 26 | 5 268 | 3 416 | 14 |
| ***Trazodone*** | 57 | 1 672 | 892 | 9 | 61 | 1 362 | 840 | 8 |
| ***Venlafaxine*** | 77 | 9 245 | 4 768 | 16 | 28 | 7 454 | 4 505 | 15 |
| ***Vilazodone*** | 104 | 2 939 | 2 009 | 3 | 583 | 1 662 | 1 662 | 2 |
| ***Vortioxetine*** | 423 | 5 767 | 3 921 | 3 | 423 | 4 134 | 3 921 | 3 |

***e-Table 2:*** *Number of patients included in all the NMAs (min, max and median) depending on treatments and the number of existing direct comparisons. The maximum number (max) corresponds to the number of patients included in the Cipriani et al. NMA.*

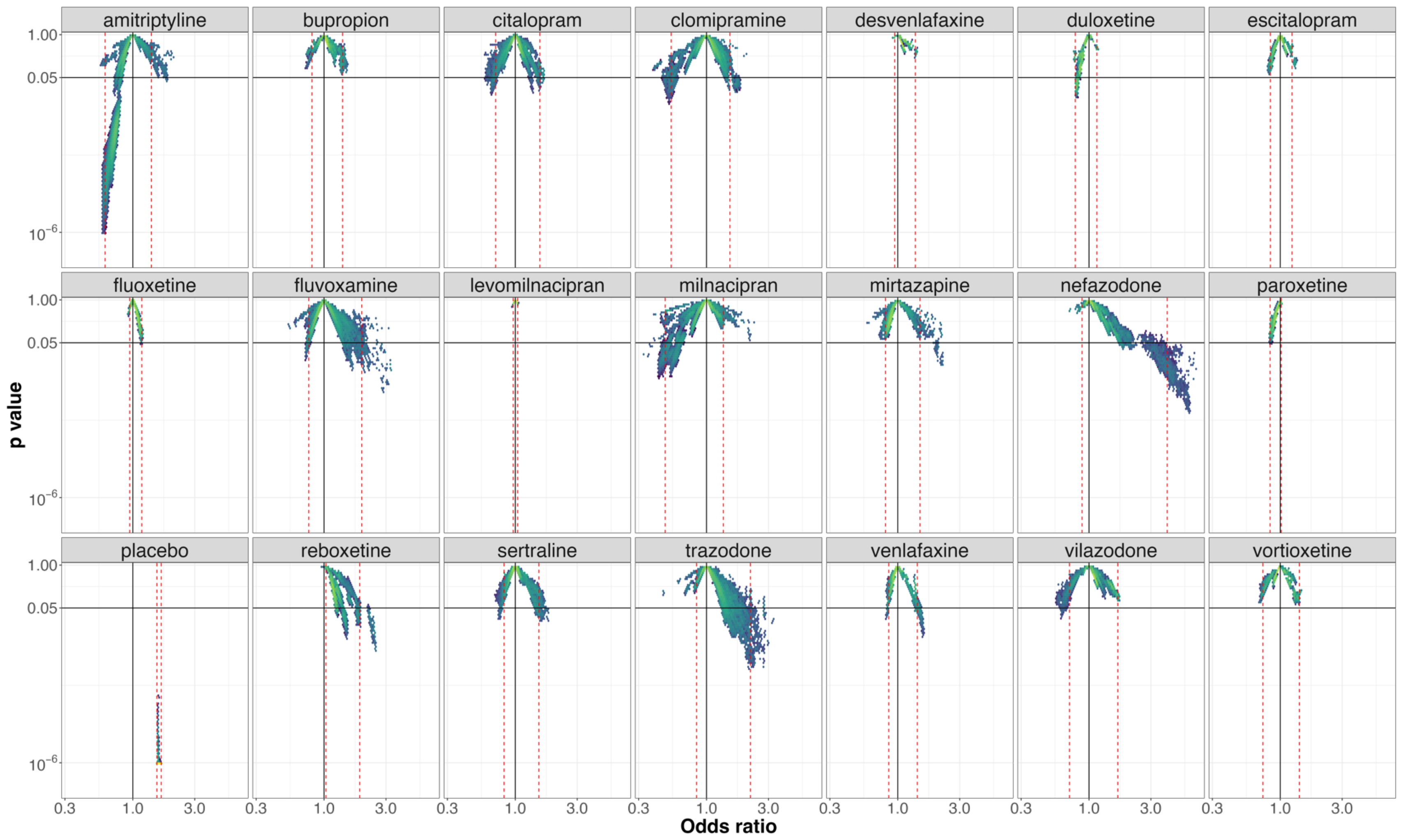


**e-figure 1:** *Vibration of effects for* treatment response *for the comparisons of agomelatine with the 20 remaining antidepressants and placebo (with the number of patients included in the most complete network for this comparison). An Odds ratio >1 favors agomelatine. The Colors indicate the log densities of network meta-analyses (yellow: high, green: moderate, blue: low) Dotted red lines illustrate the 1^st^ and 99^th^ percentiles.*

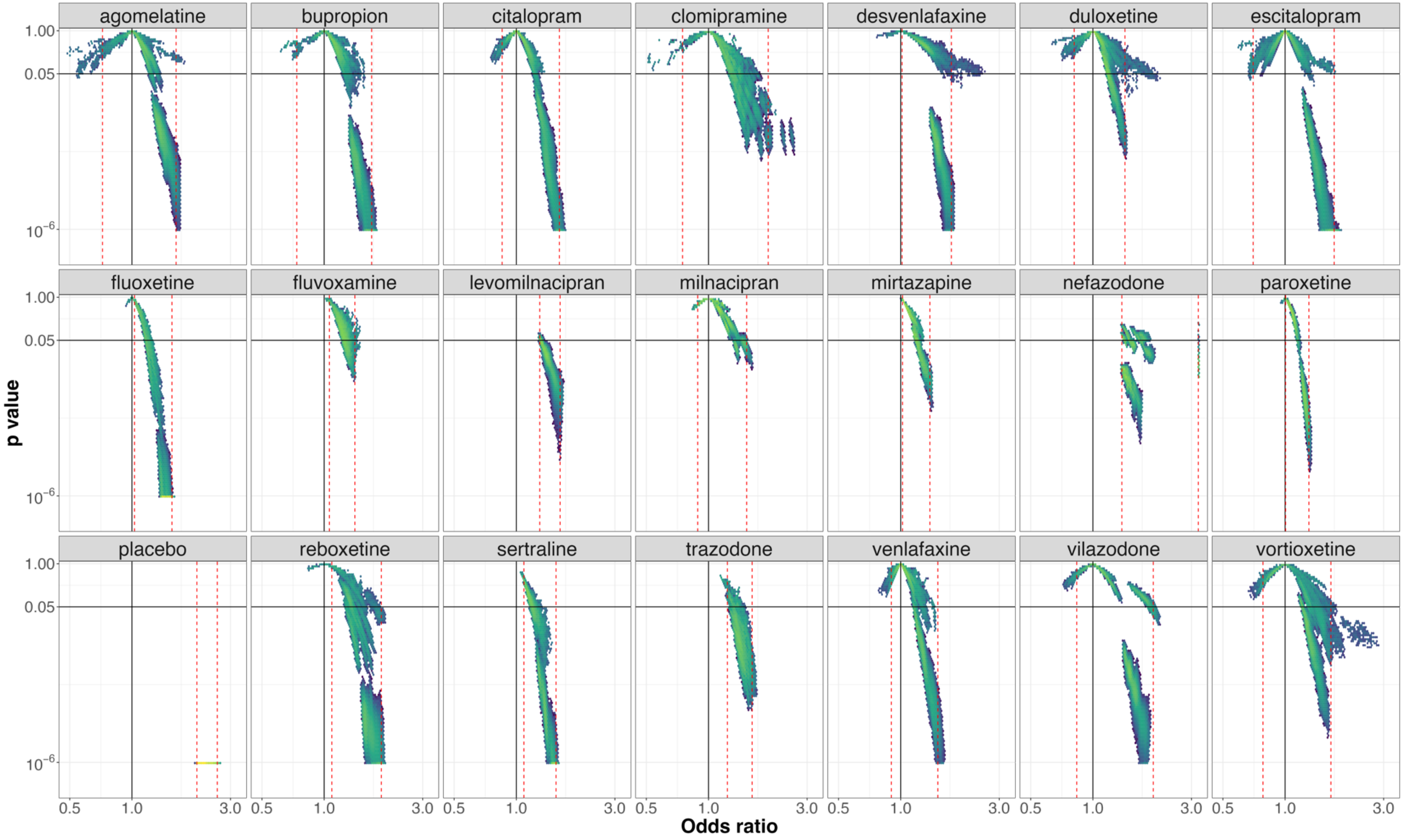


**e-figure 2:** *Vibration of effects for* treatment response *for the comparisons of amitriptyline with the 20 remaining antidepressants and placebo (with the number of patients included in the most complete network for this comparison). An Odds ratio >1 favors amitriptyline. The Colors indicate the log densities of network meta-analyses (yellow: high, green: moderate, blue: low) Dotted red lines illustrate the 1st and 99th percentiles.*

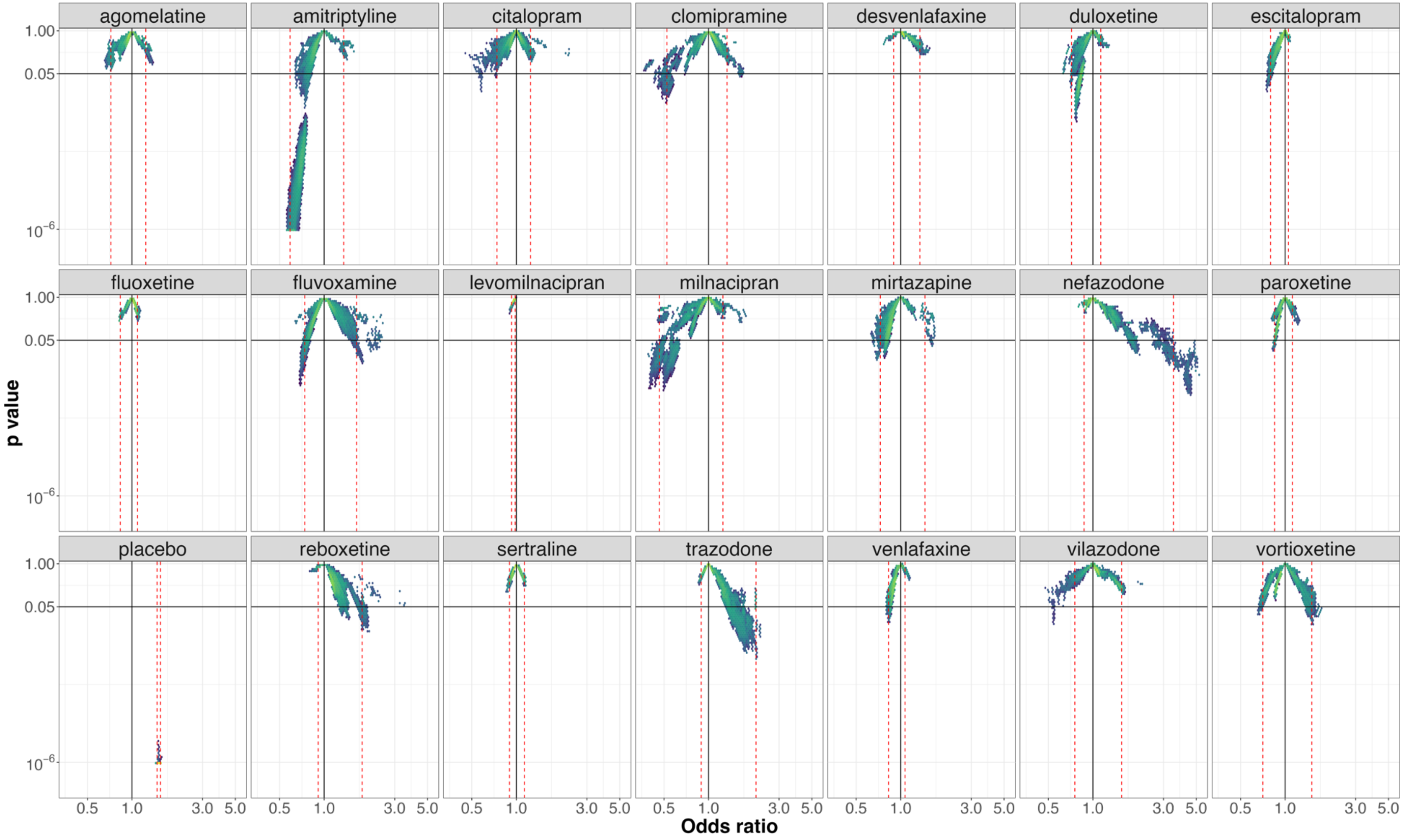


**e-figure 3:** *Vibration of effects for* treatment response *for the comparisons of bupropion with the 20 remaining antidepressants and placebo (with the number of patients included in the most complete network for this comparison). An Odds ratio >1 favors bupropion. The Colors indicate the log densities of network meta-analyses (yellow: high, green: moderate, blue: low) Dotted red lines illustrate the 1st and 99th percentiles.*

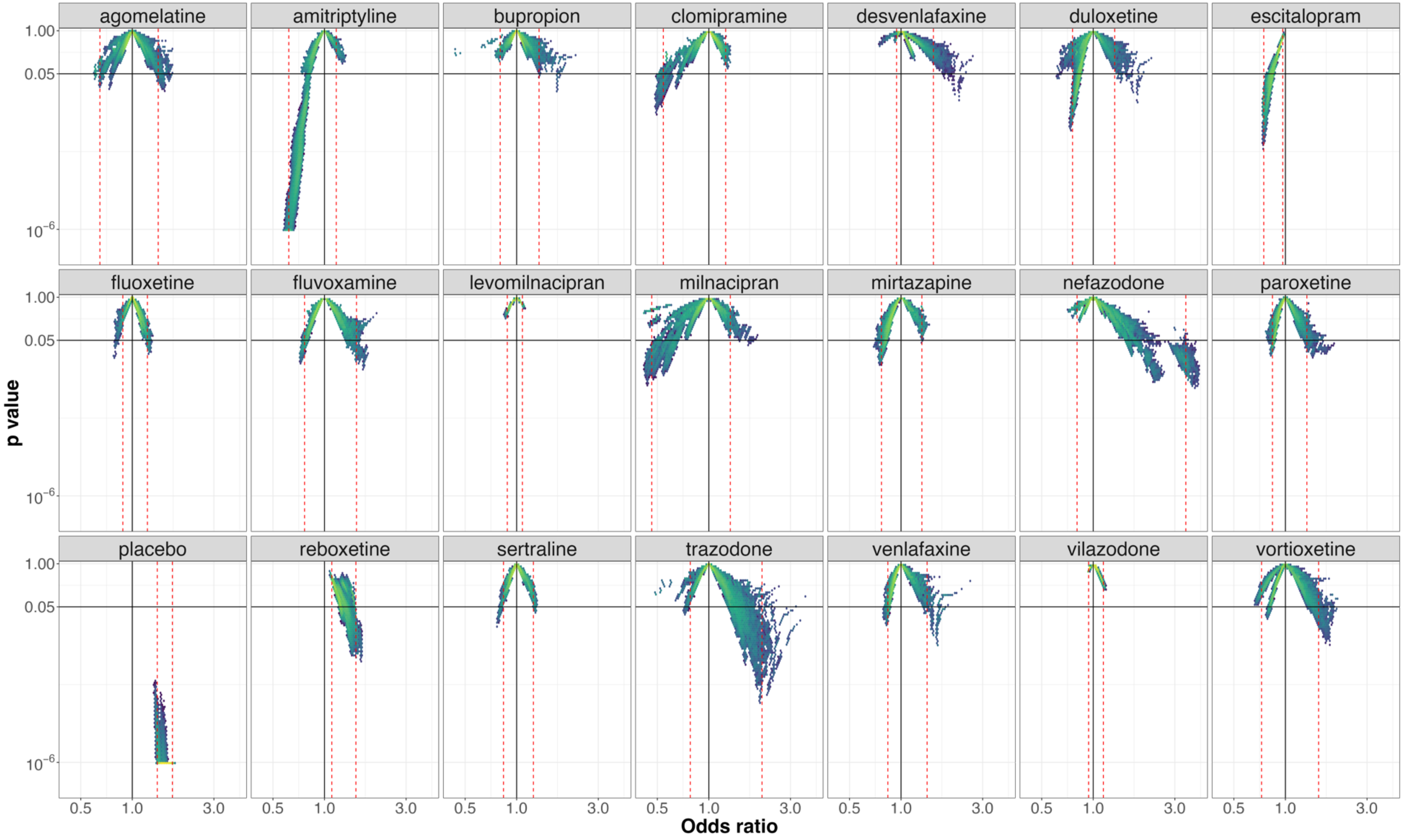


**e-figure 4:** *Vibration of effects for* treatment response *for the comparisons of citalopram with the 20 remaining antidepressants and placebo (with the number of patients included in the most complete network for this comparison). An Odds ratio >1 favors citalopram. The Colors indicate the log densities of network meta-analyses (yellow: high, green: moderate, blue: low) Dotted red lines illustrate the 1$^{st}$ and 99$^{th}$ percentiles.*

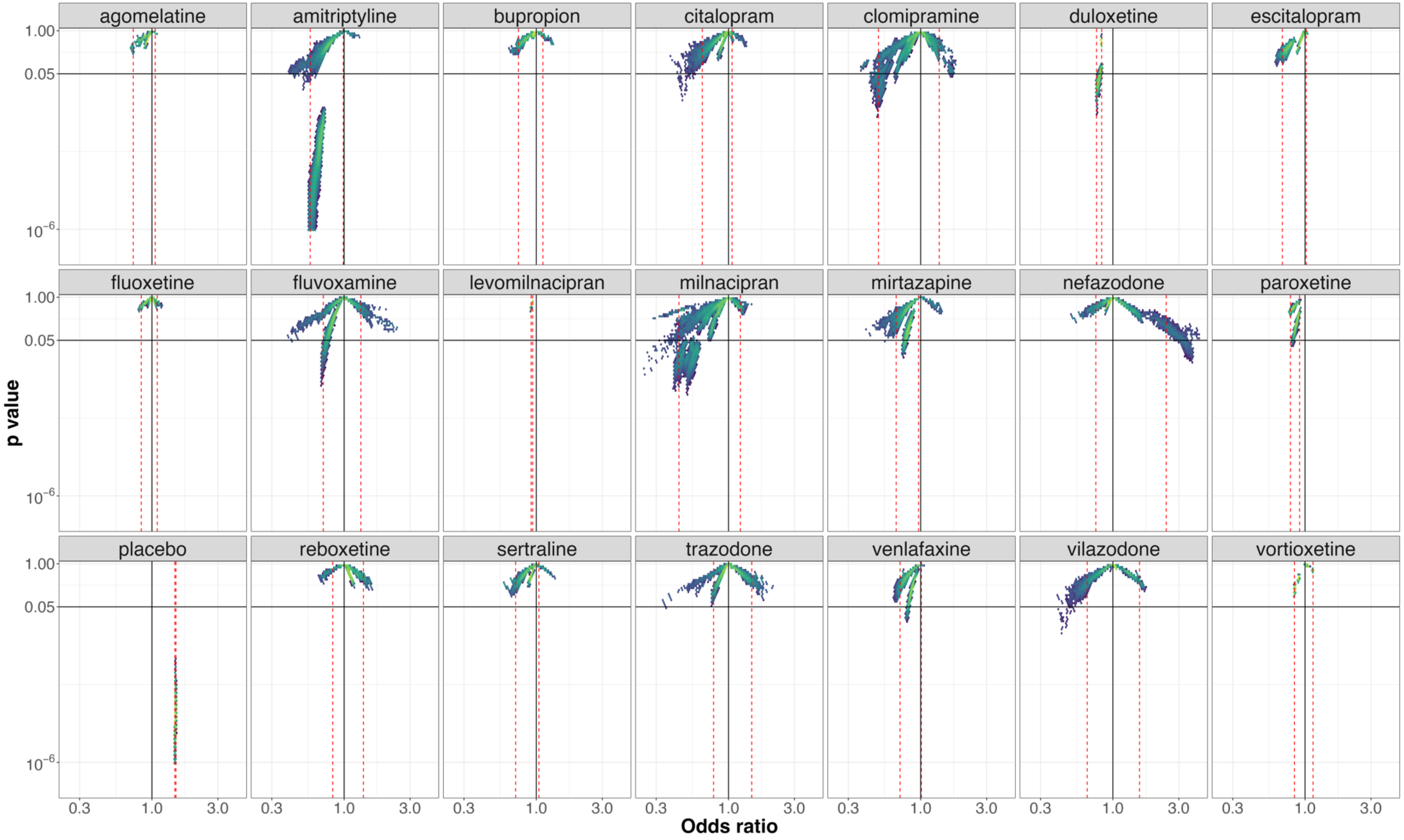


**e-figure 5:** *Vibration of effects for* treatment response *for the comparisons of desvenlafaxine with the 20 remaining antidepressants and placebo (with the number of patients included in the most complete network for this comparison). An Odds ratio >1 favors desvenlafaxine. The Colors indicate the log densities of network meta-analyses (yellow: high, green: moderate, blue: low) Dotted red lines illustrate the 1$^{st}$ and 99$^{th}$ percentiles.*

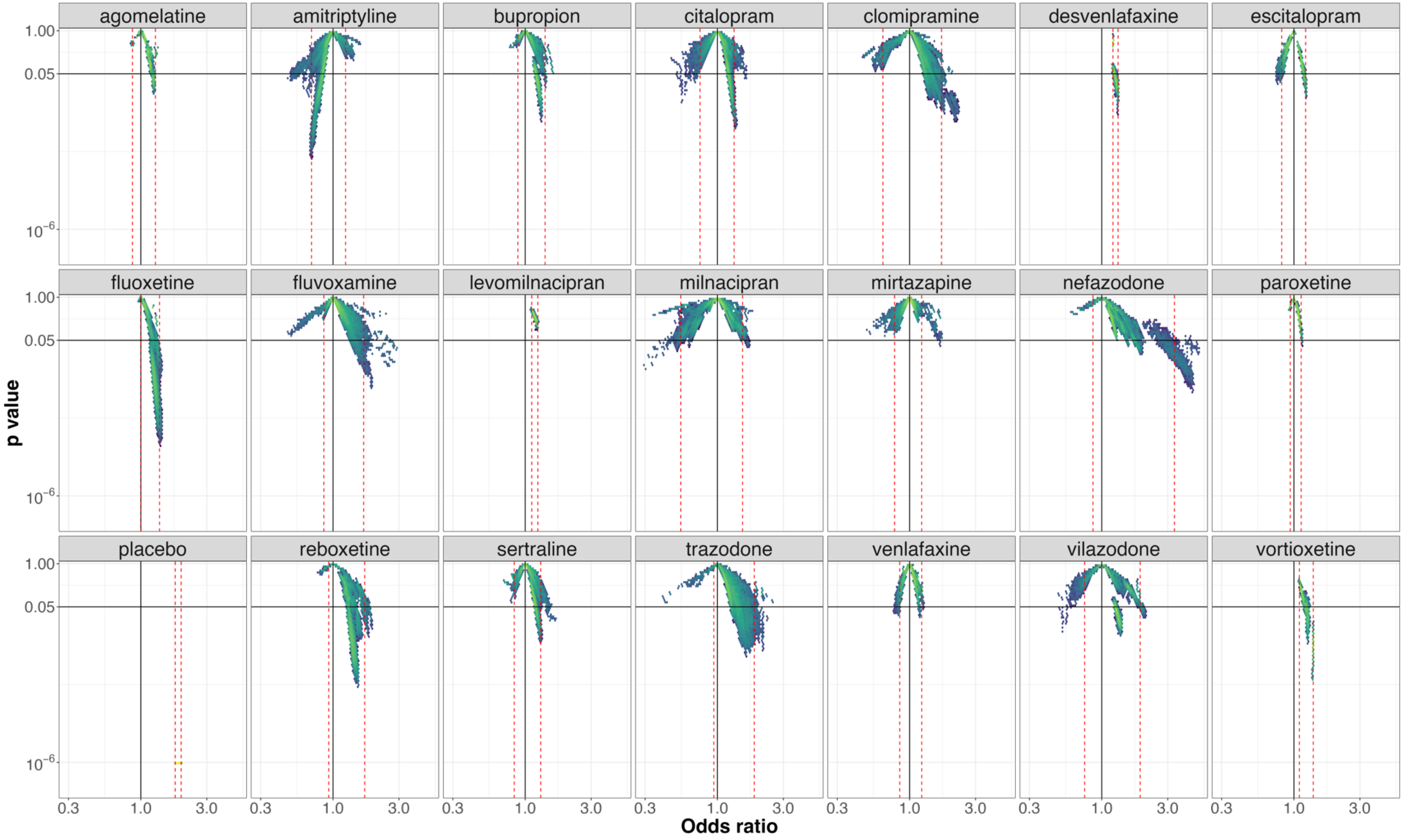


**e-figure 6:** *Vibration of effects for* treatment response *for the comparisons of duloxetine with the 20 remaining antidepressants and placebo (with the number of patients included in the most complete network for this comparison). An Odds ratio >1 favors duloxetine. The Colors indicate the log densities of network meta-analyses (yellow: high, green: moderate, blue: low) Dotted red lines illustrate the 1st and 99th percentiles.*

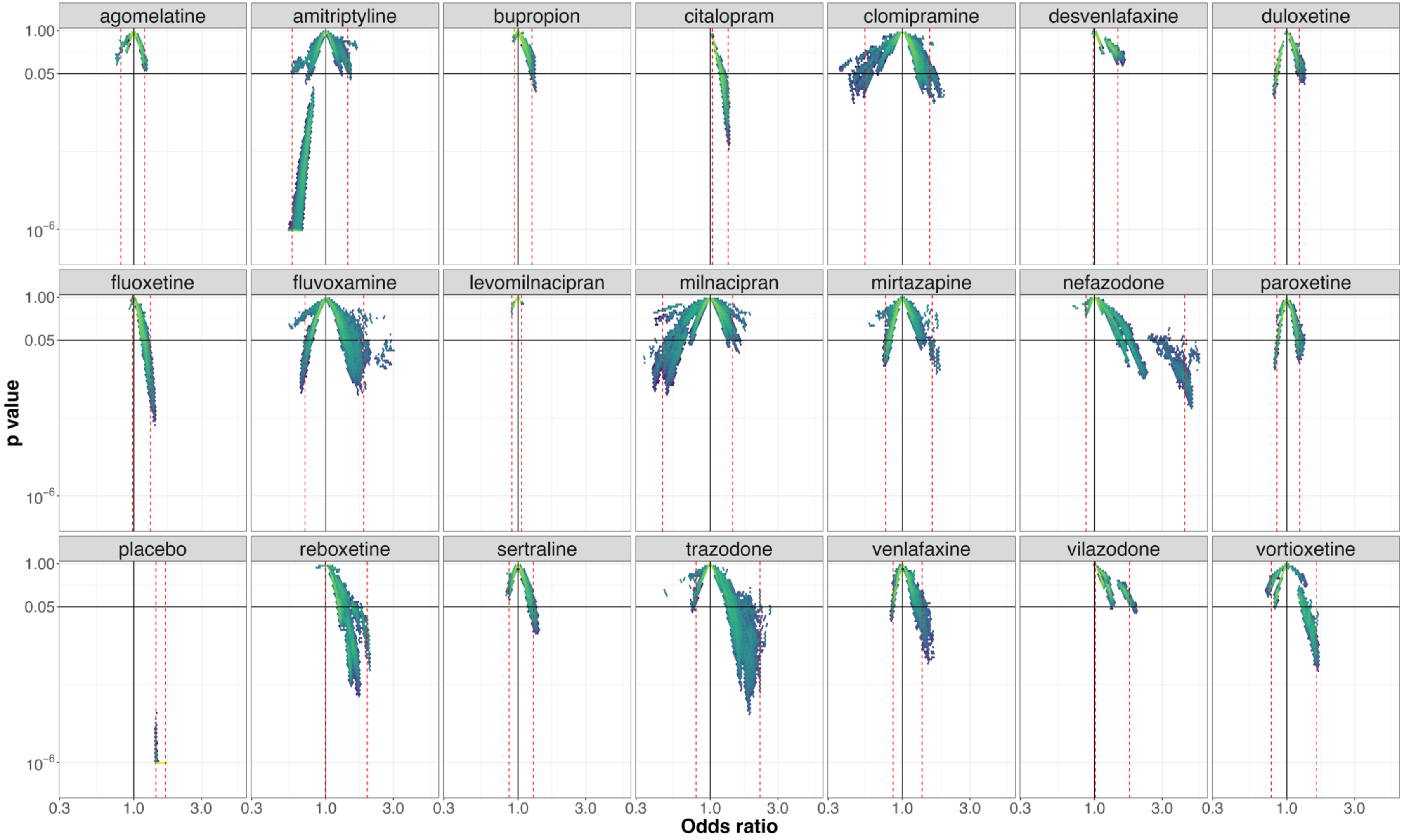


**e-figure 7:** *Vibration of effects for* treatment response *for the comparisons of escitalopram with the 20 remaining antidepressants and placebo (with the number of patients included in the most complete network for this comparison). An Odds ratio >1 favors escitalopram. The Colors indicate the log densities of network meta-analyses (yellow: high, green: moderate, blue: low) Dotted red lines illustrate the 1st and 99th percentiles.*

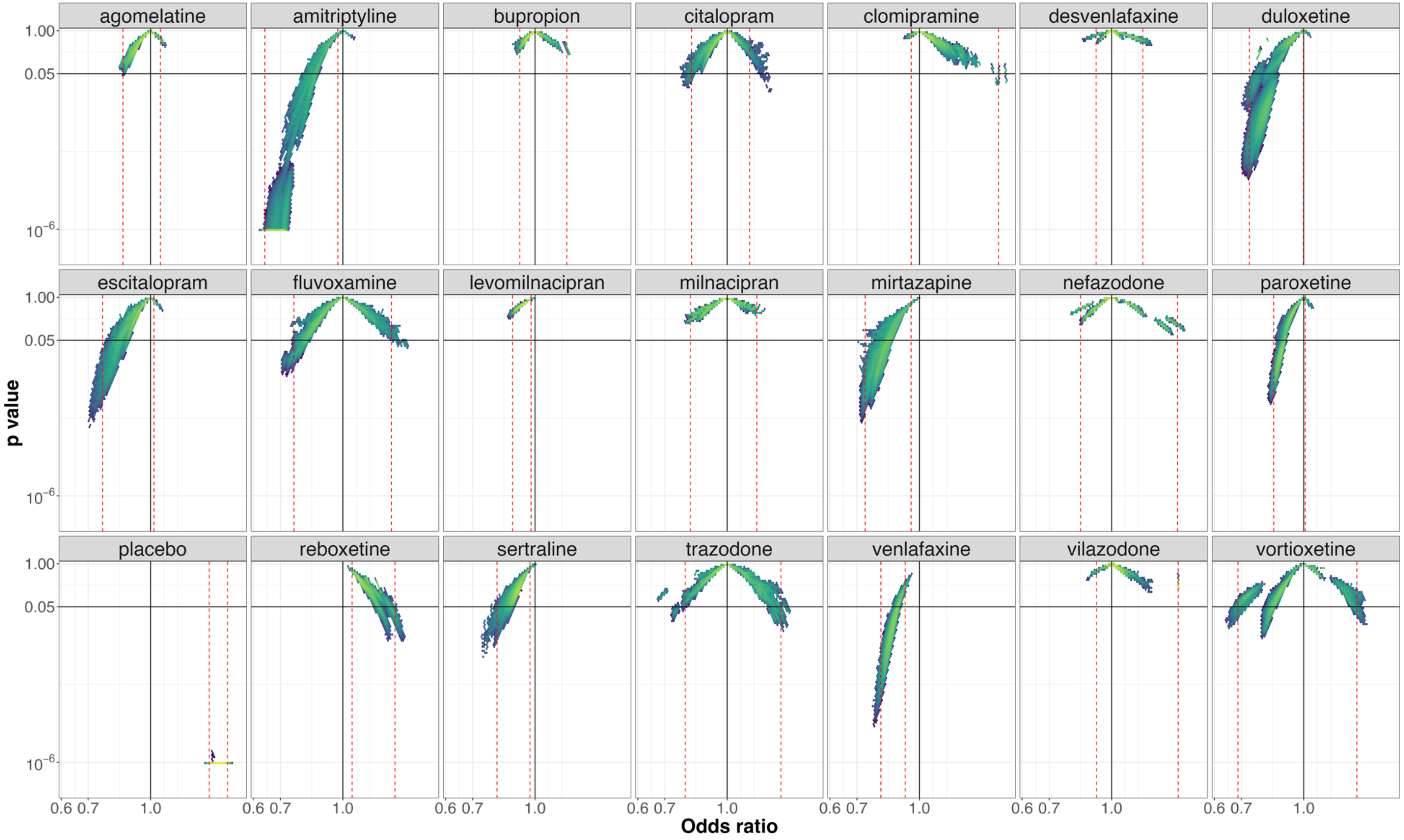


**e-figure 8:** *Vibration of effects for* treatment response *for the comparisons of fluoxetine with the 20 remaining antidepressants and placebo (with the number of patients included in the most complete network for this comparison). An Odds ratio >1 favors fluoxetine. The Colors indicate the log densities of network meta-analyses (yellow: high, green: moderate, blue: low) Dotted red lines illustrate the 1$^{st}$ and 99$^{th}$ percentiles.*

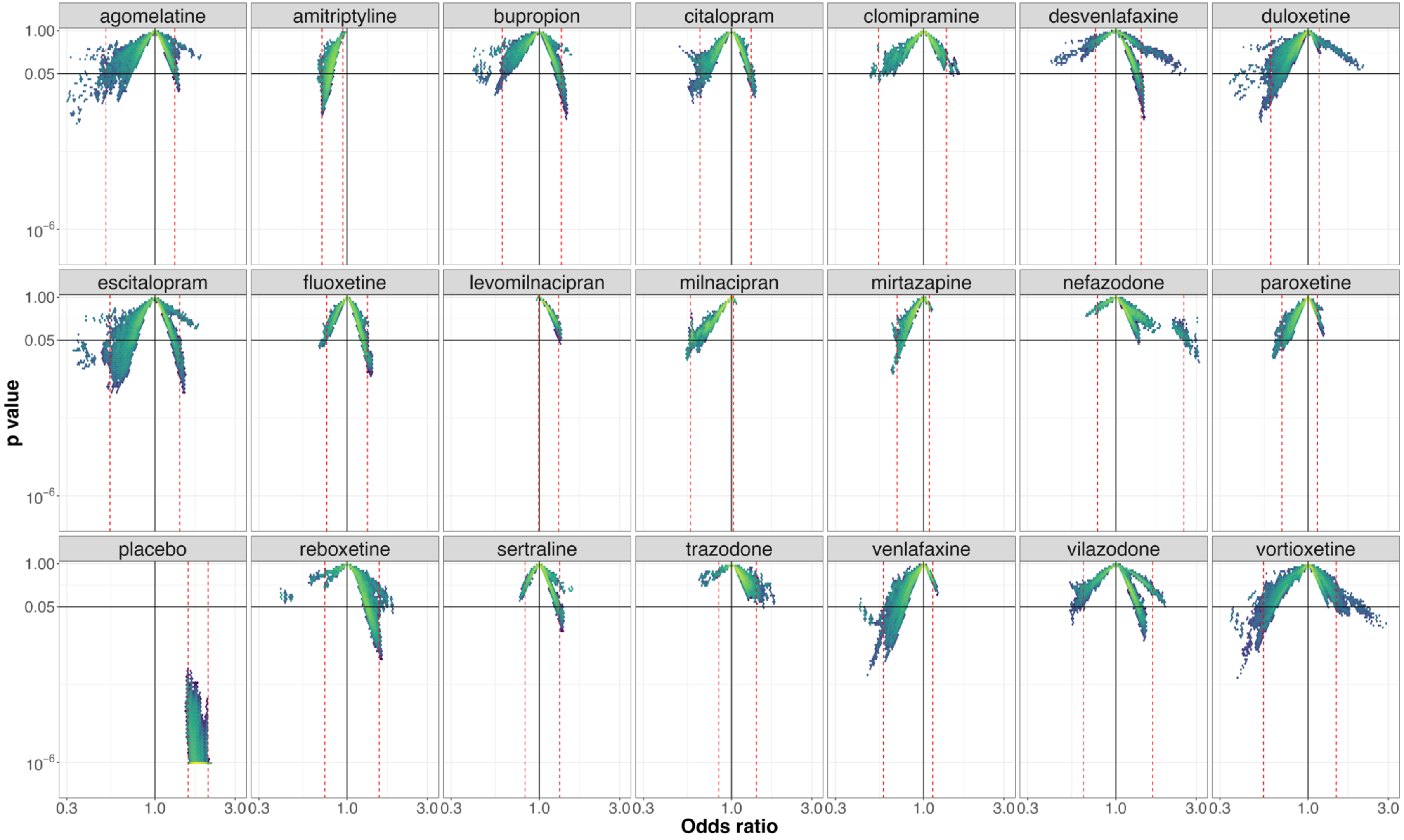


**e-figure 9:** *Vibration of effects for* treatment response *for the comparisons of fluvoxamine with the 20 remaining antidepressants and placebo (with the number of patients included in the most complete network for this comparison). An Odds ratio >1 favors fluvoxamine. The Colors indicate the log densities of network meta-analyses (yellow: high, green: moderate, blue: low) Dotted red lines illustrate the $1^{st}$ and $99^{th}$ percentiles.*

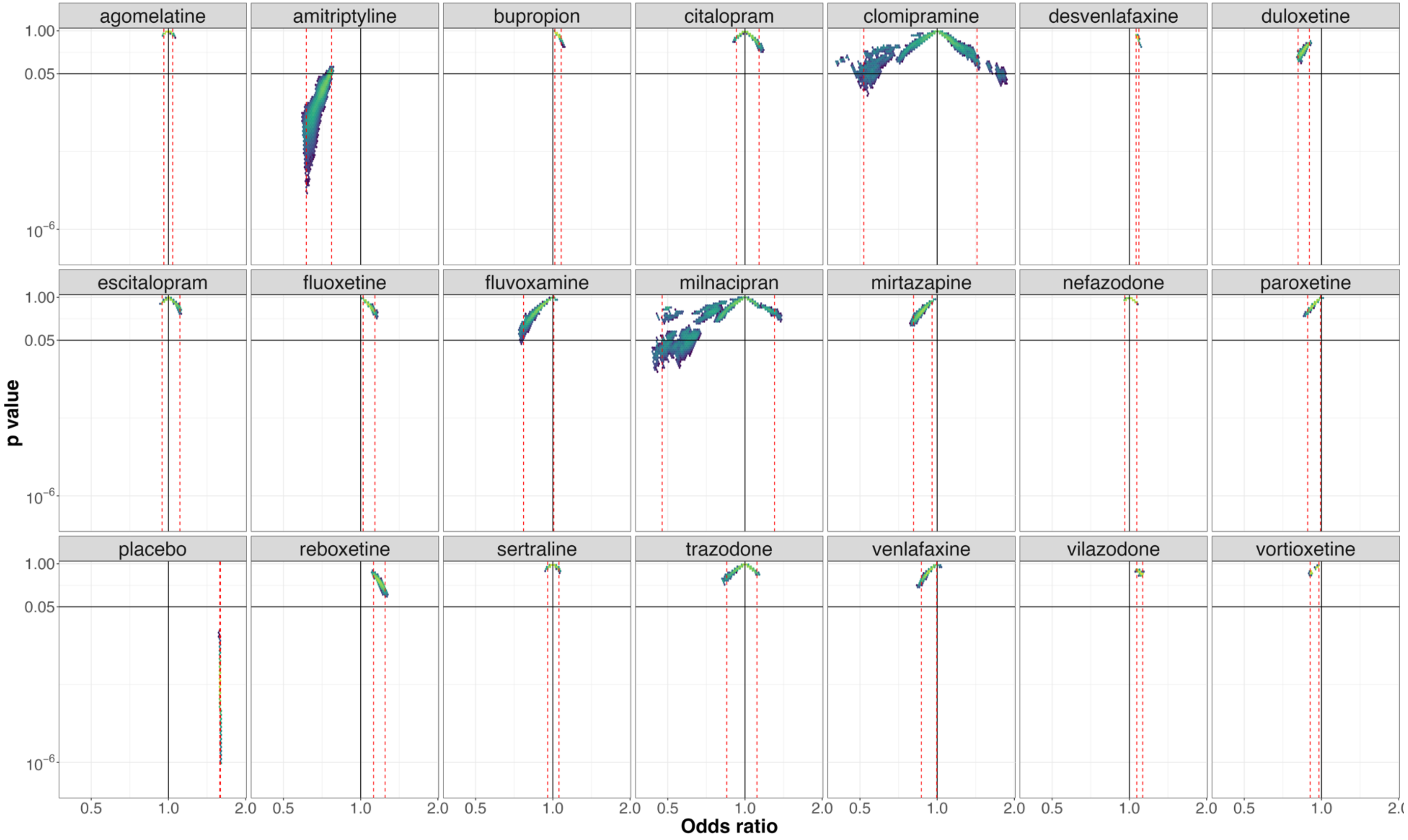


**e-figure 10:** *Vibration of effects for* treatment response *for the comparisons of levomilnacipran with the 20 remaining antidepressants and placebo (with the number of patients included in the most complete network for this comparison). An Odds ratio >1 favors levomilnacipran. The Colors indicate the log densities of network meta-analyses (yellow: high, green: moderate, blue: low) Dotted red lines illustrate the 1st and 99th percentiles.*

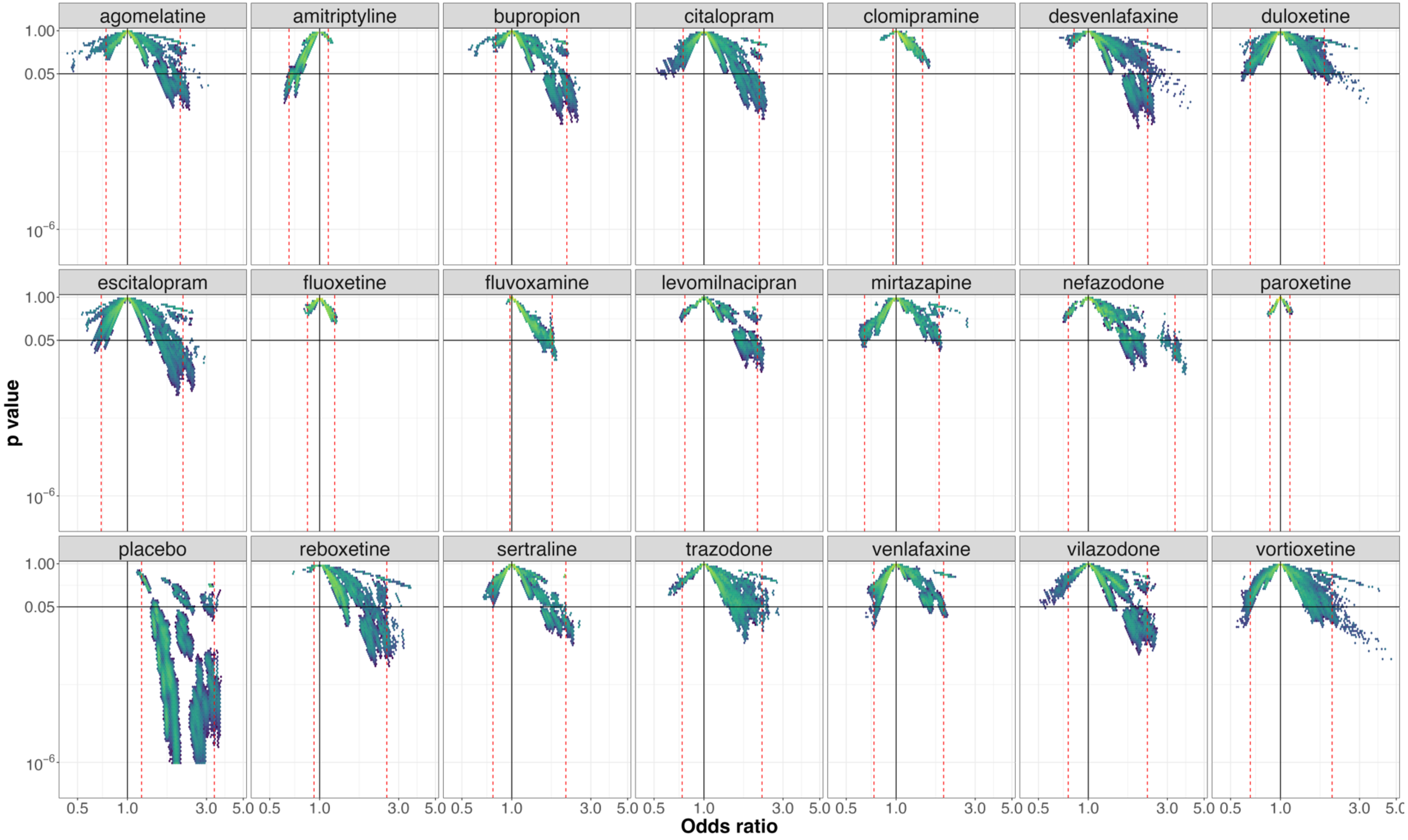


**e-figure 11:** *Vibration of effects for treatment response for the comparisons of milnacipran with the 20 remaining antidepressants and placebo (with the number of patients included in the most complete network for this comparison). An Odds ratio >1 favors milnacipran. The Colors indicate the log densities of network meta-analyses (yellow: high, green: moderate, blue: low) Dotted red lines illustrate the 1st and 99th percentiles.*

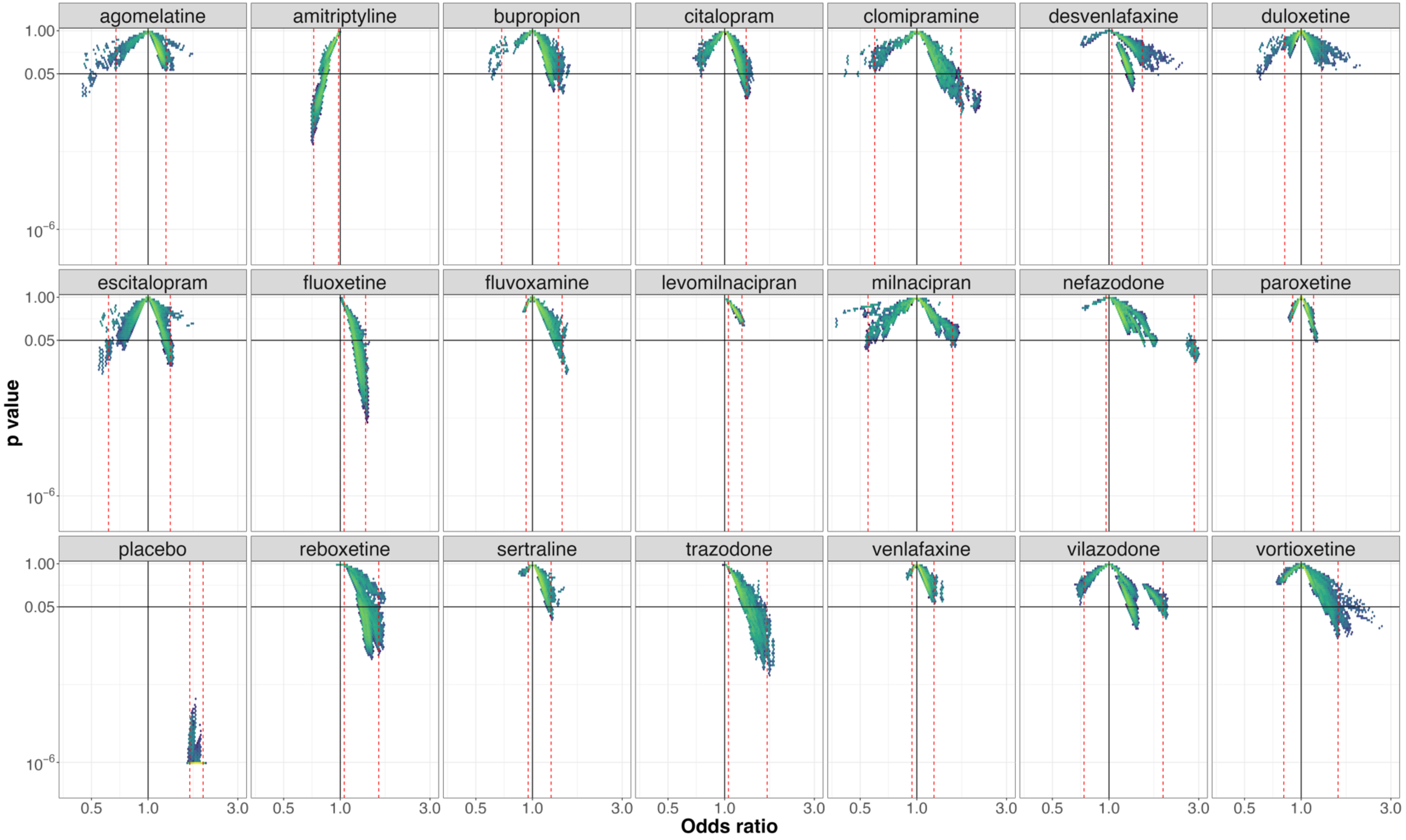


**e-figure 12:** *Vibration of effects for treatment response for the comparisons of mirtazapine with the 20 remaining antidepressants and placebo (with the number of patients included in the most complete network for this comparison). An Odds ratio >1 favors mirtazapine. The Colors indicate the log densities of network meta-analyses (yellow: high, green: moderate, blue: low) Dotted red lines illustrate the 1st and 99th percentiles.*

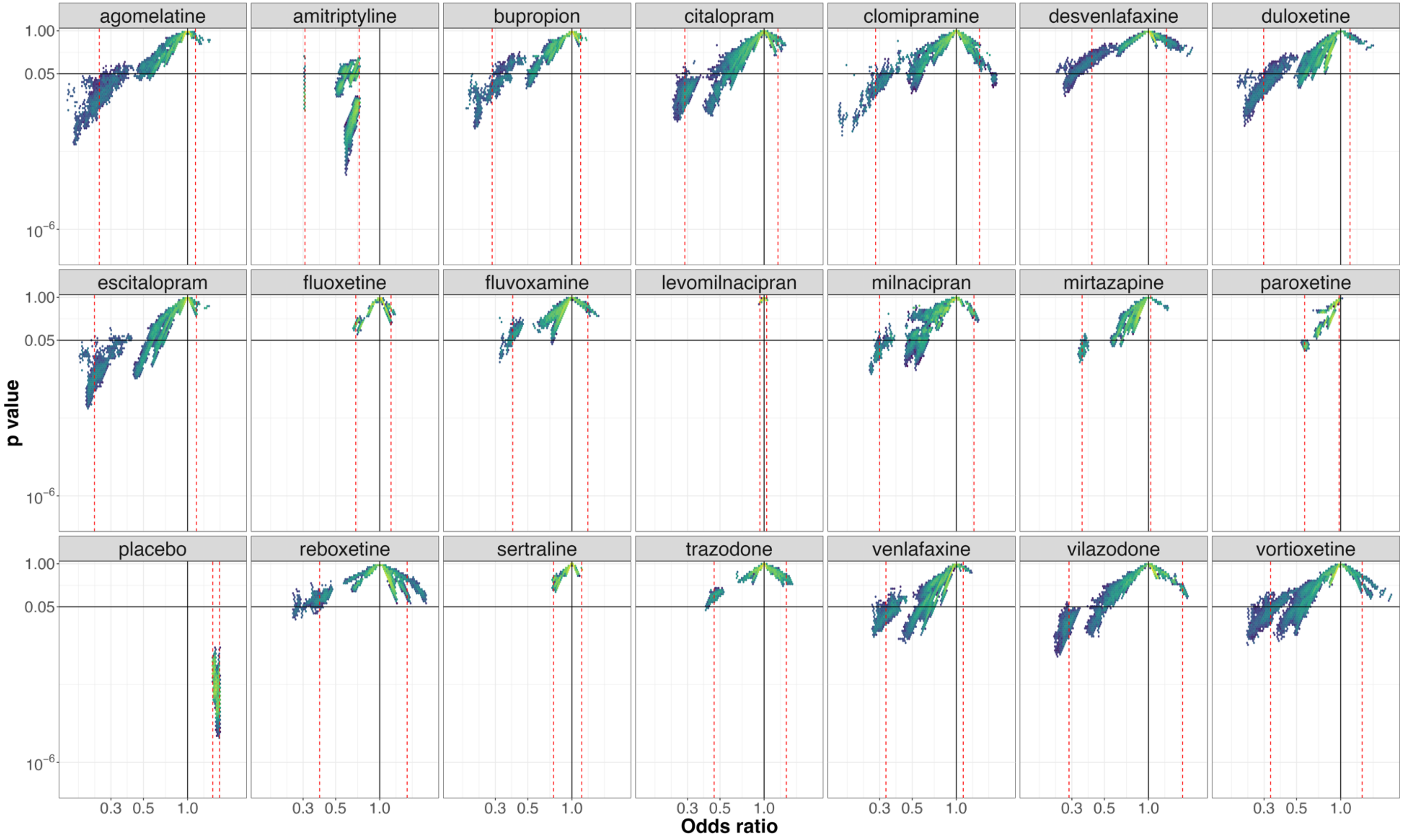


**e-figure 13:** *Vibration of effects for treatment response for the comparisons of nefazodone with the 20 remaining antidepressants and placebo (with the number of patients included in the most complete network for this comparison). An Odds ratio >1 favors nefazodone. The Colors indicate the log densities of network meta-analyses (yellow: high, green: moderate, blue: low) Dotted red lines illustrate the 1st and 99th percentiles.*

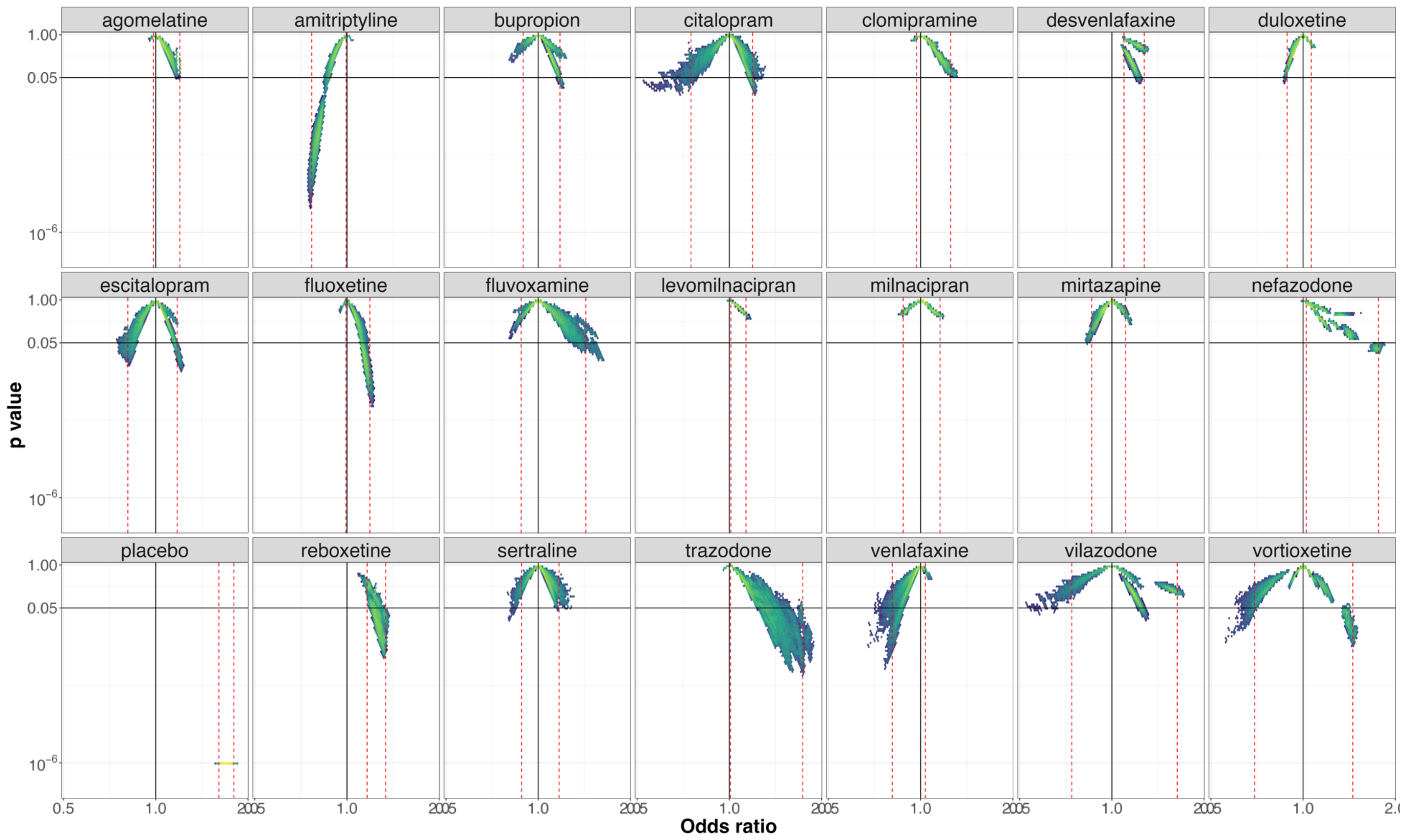


**e-figure 14:** *Vibration of effects for treatment response for the comparisons of paroxetine with the 20 remaining antidepressants and placebo (with the number of patients included in the most complete network for this comparison). An Odds ratio >1 favors paroxetine. The Colors indicate the log densities of network meta-analyses (yellow: high, green: moderate, blue: low) Dotted red lines illustrate the 1st and 99th percentiles.*

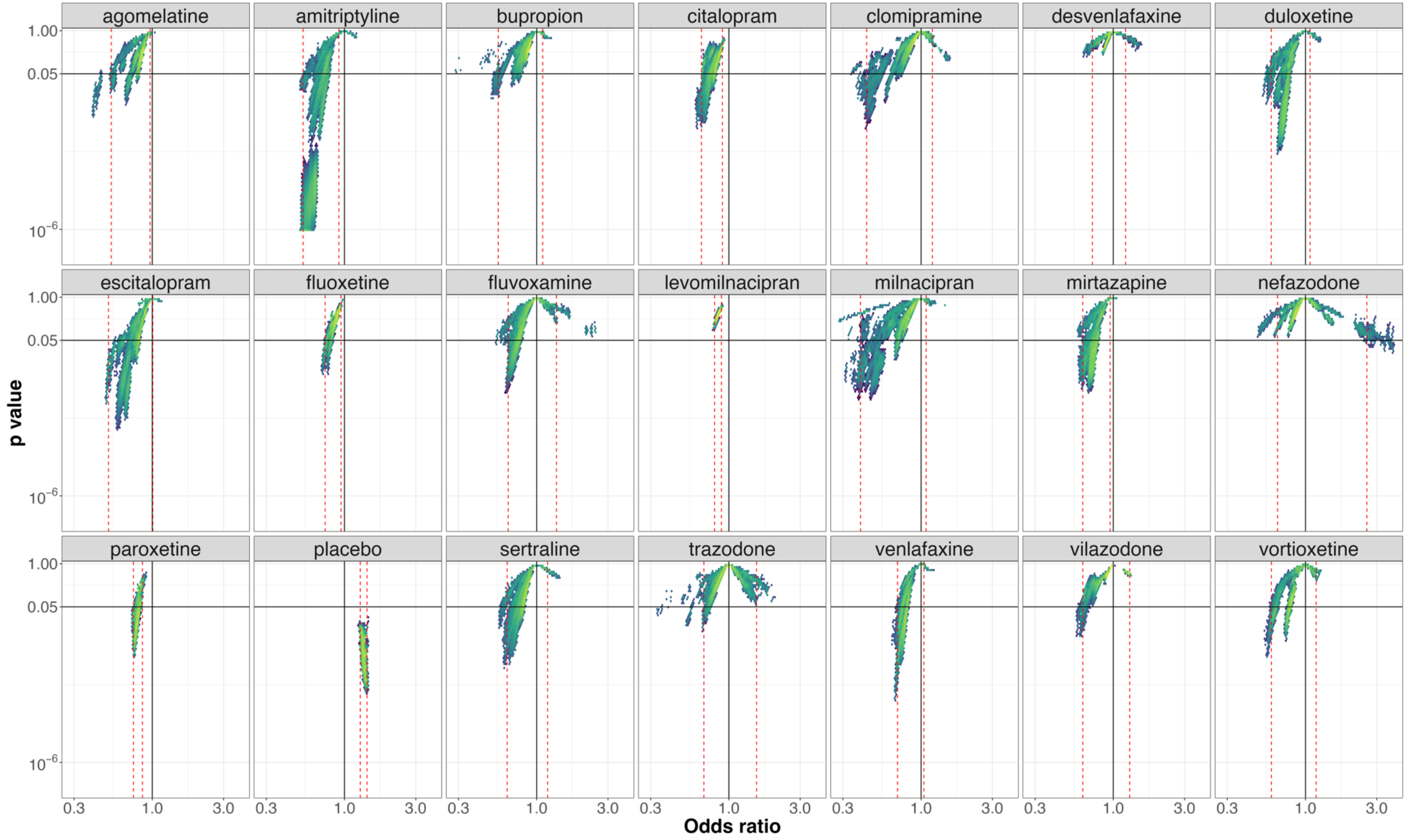


**e-figure 15:** *Vibration of effects for treatment response for the comparisons of reboxetine with the 20 remaining antidepressants and placebo (with the number of patients included in the most complete network for this comparison). An Odds ratio >1 favors reboxetine. The Colors indicate the log densities of network meta-analyses (yellow: high, green: moderate, blue: low) Dotted red lines illustrate the 1st and 99th percentiles.*

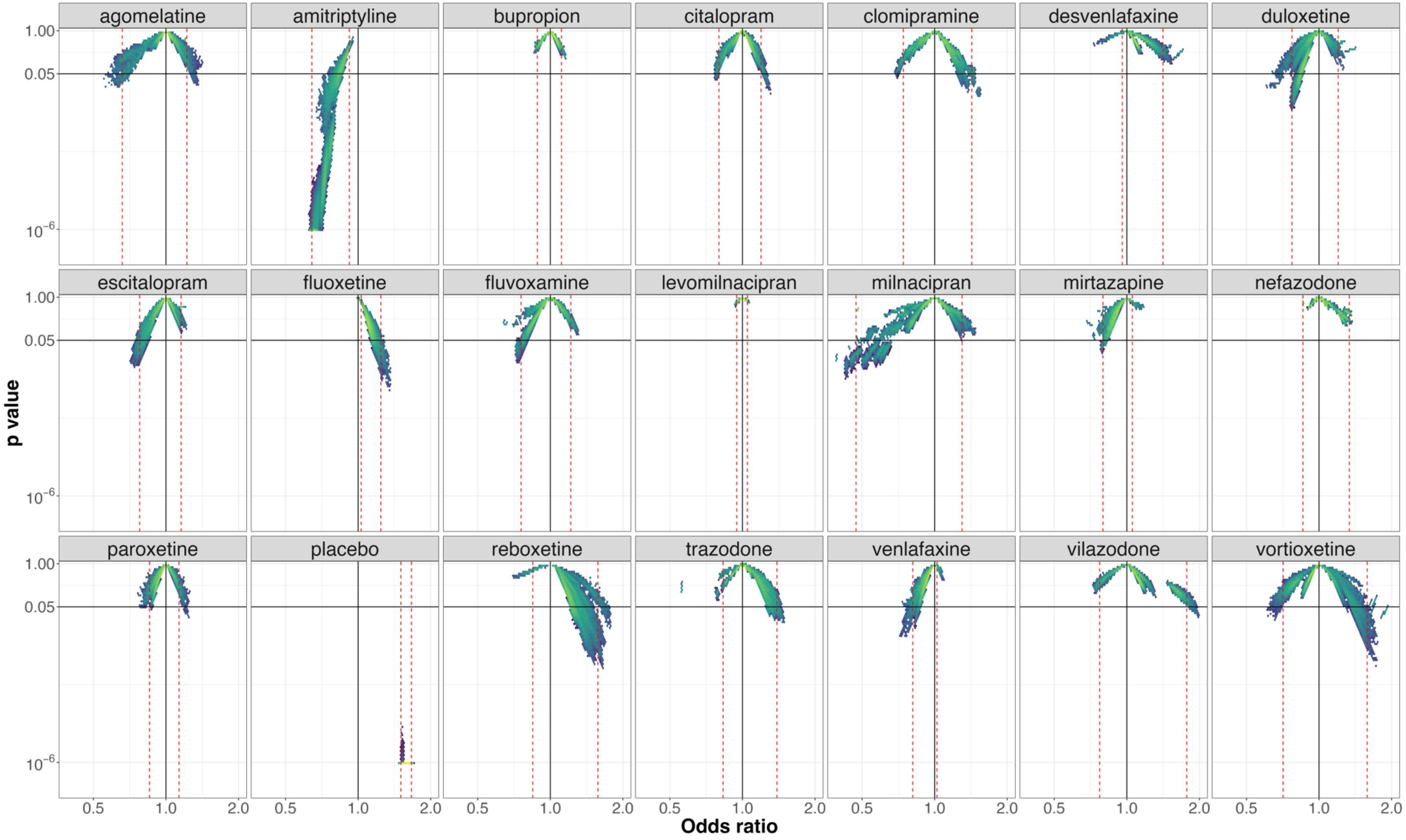


**e-figure 16:** *Vibration of effects for treatment response for the comparisons of sertraline with the 20 remaining antidepressants and placebo (with the number of patients included in the most complete network for this comparison). An Odds ratio >1 favors clomipramine. The Colors indicate the log densities of network meta-analyses (yellow: high, green: moderate, blue: low) Dotted red lines illustrate the 1st and 99th percentiles.*

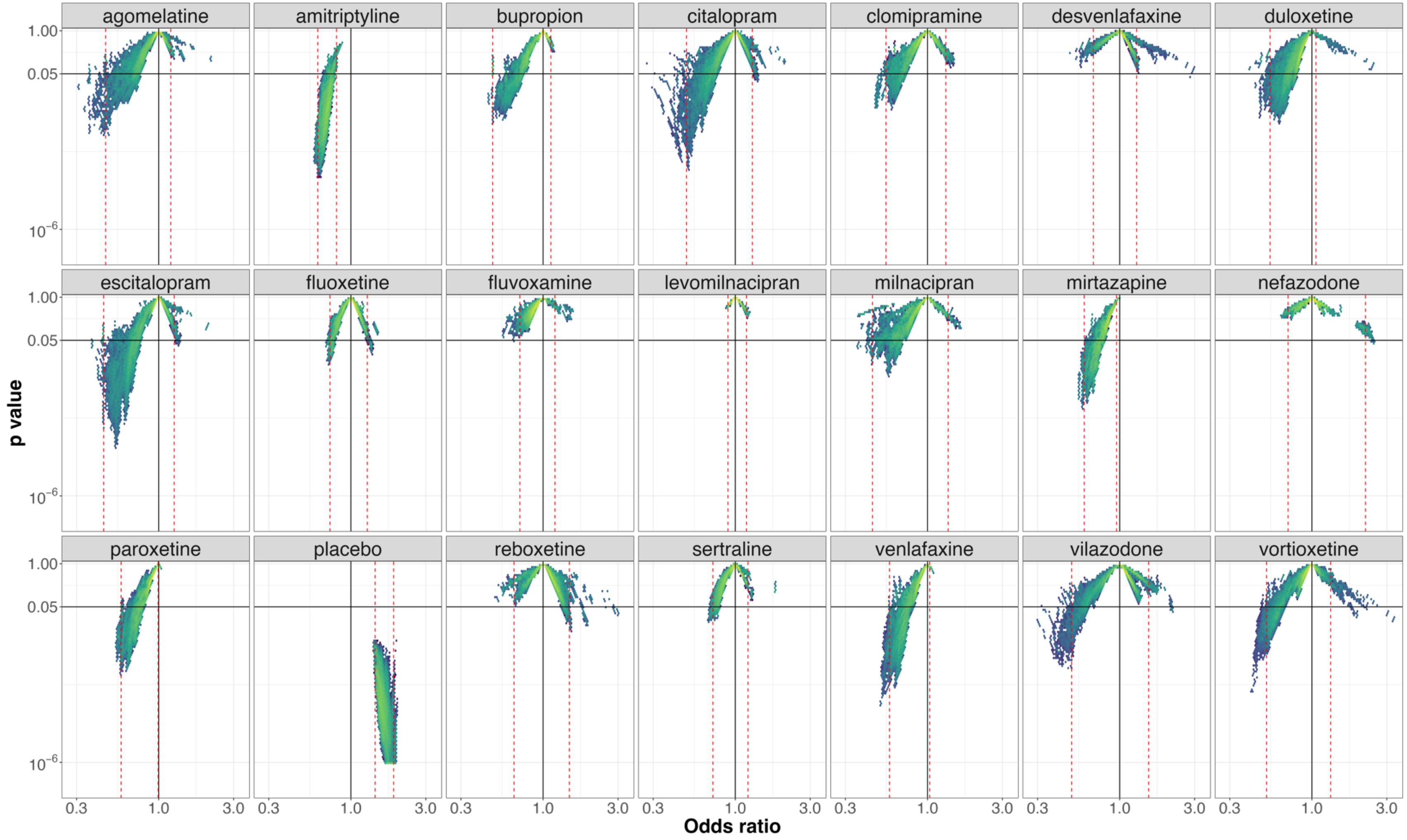


**e-figure 17:** *Vibration of effects for treatment response for the comparisons of trazodone with the 20 remaining antidepressants and placebo (with the number of patients included in the most complete network for this comparison). An Odds ratio >1 favors trazodone. The Colors indicate the log densities of network meta-analyses (yellow: high, green: moderate, blue: low) Dotted red lines illustrate the 1st and 99th percentiles.*

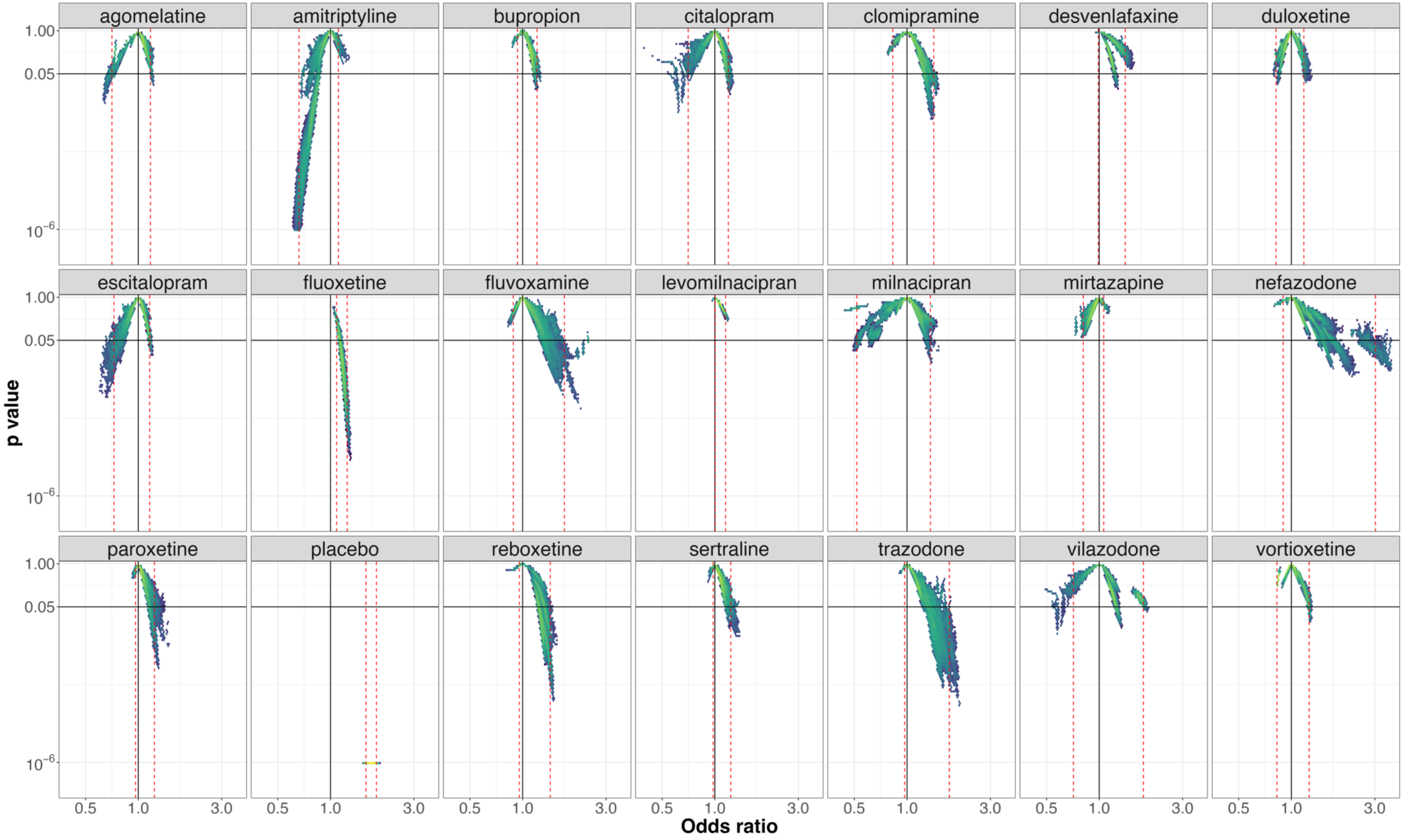


**e-figure 18:** *Vibration of effects for treatment response for the comparisons of venlafaxine with the 20 remaining antidepressants and placebo (with the number of patients included in the most complete network for this comparison). An Odds ratio >1 favors venlafaxine. The Colors indicate the log densities of network meta-analyses (yellow: high, green: moderate, blue: low) Dotted red lines illustrate the 1st and 99th percentiles.*

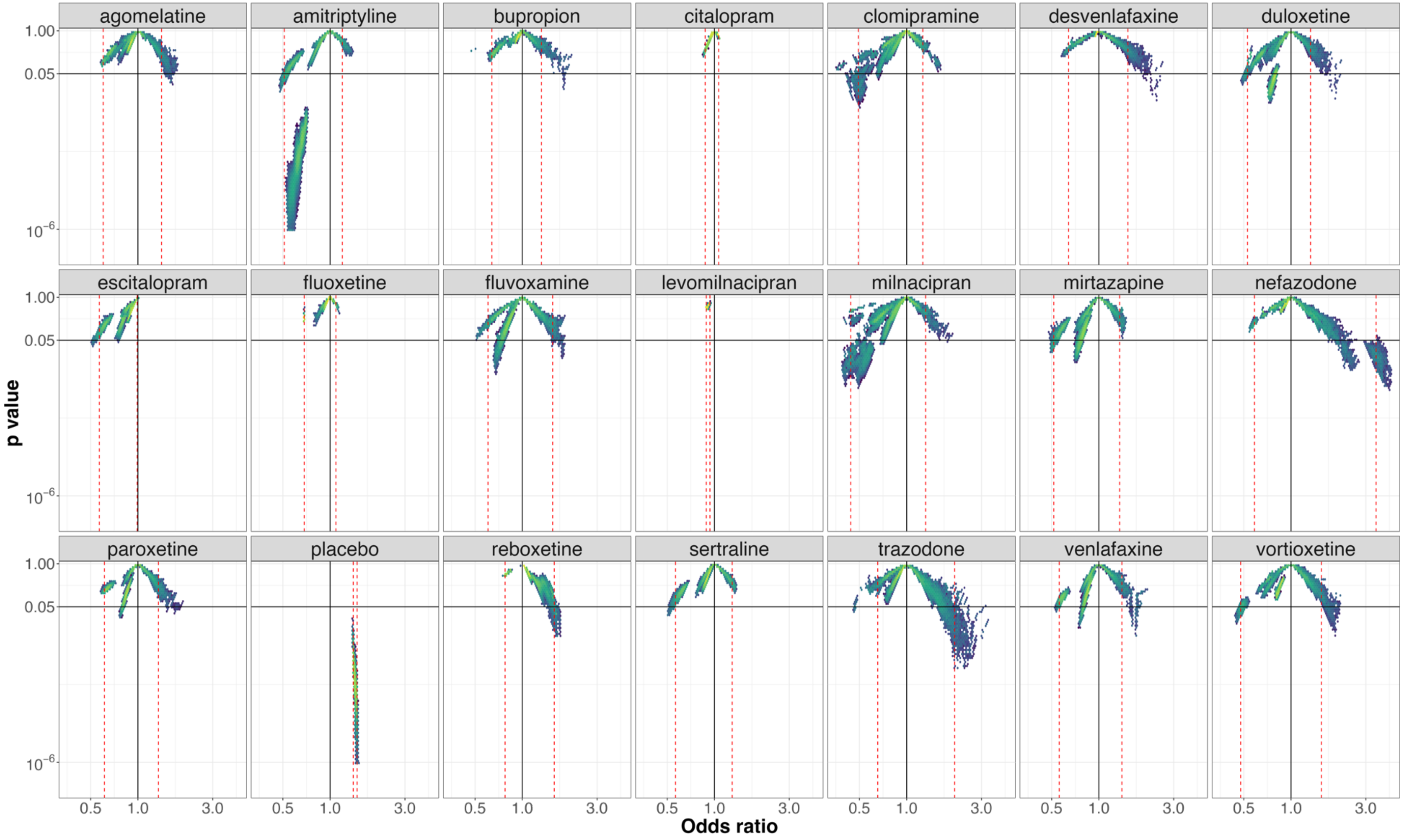


**e-figure 19:** *Vibration of effects for treatment response for the comparisons of vilazodone with the 20 remaining antidepressants and placebo (with the number of patients included in the most complete network for this comparison). An Odds ratio >1 favors vilazodone. The Colors indicate the log densities of network meta-analyses (yellow: high, green: moderate, blue: low) Dotted red lines illustrate the 1$^{st}$ and 99$^{th}$ percentiles.*

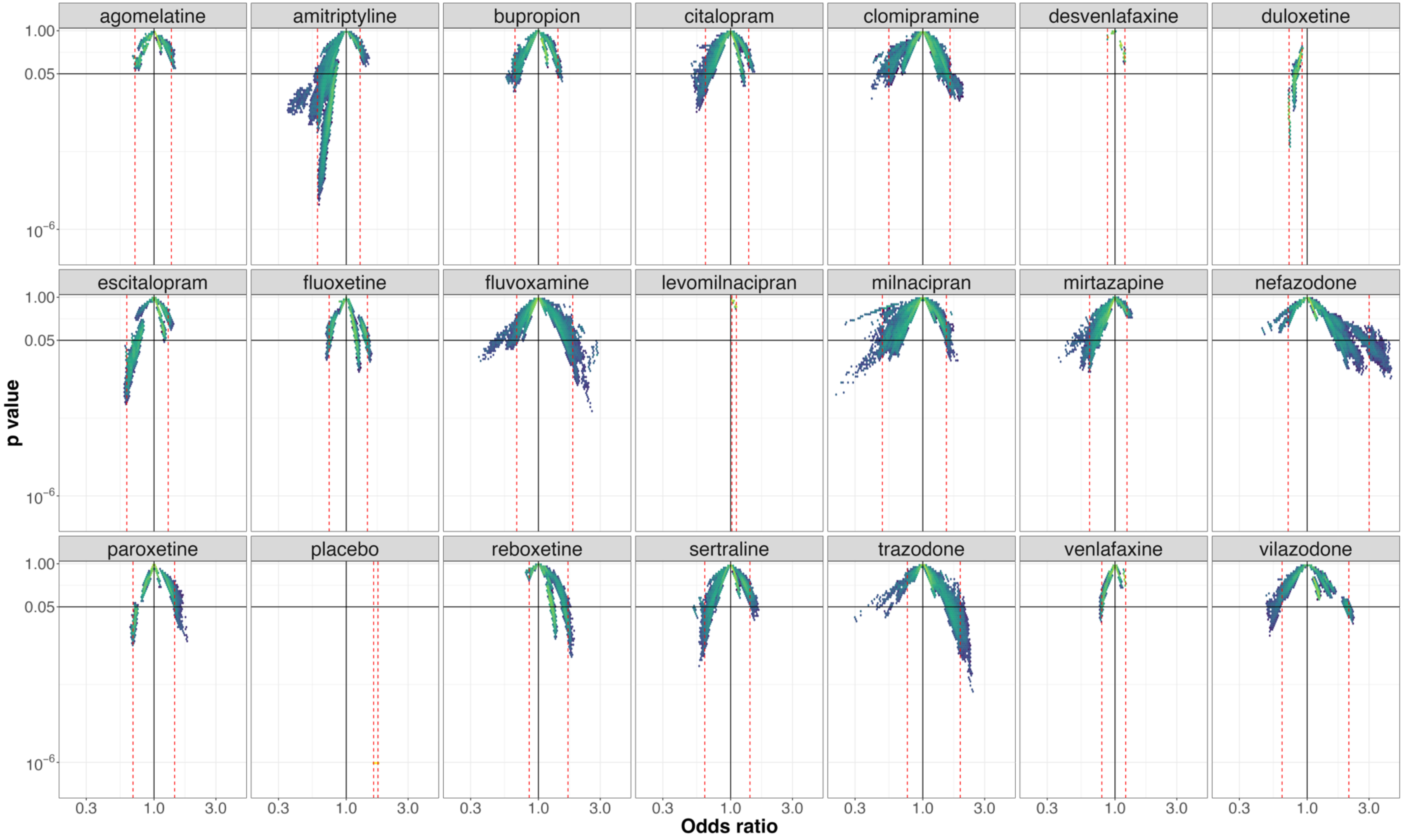


**e-figure 20:** *Vibration of effects for treatment response for the comparisons of vortioxetine with the 20 remaining antidepressants and placebo (with the number of patients included in the most complete network for this comparison). An Odds ratio >1 favors vortioxetine. The Colors indicate the log densities of network meta-analyses (yellow: high, green: moderate, blue: low) Dotted red lines illustrate the 1$^{st}$ and 99$^{th}$ percentiles.*

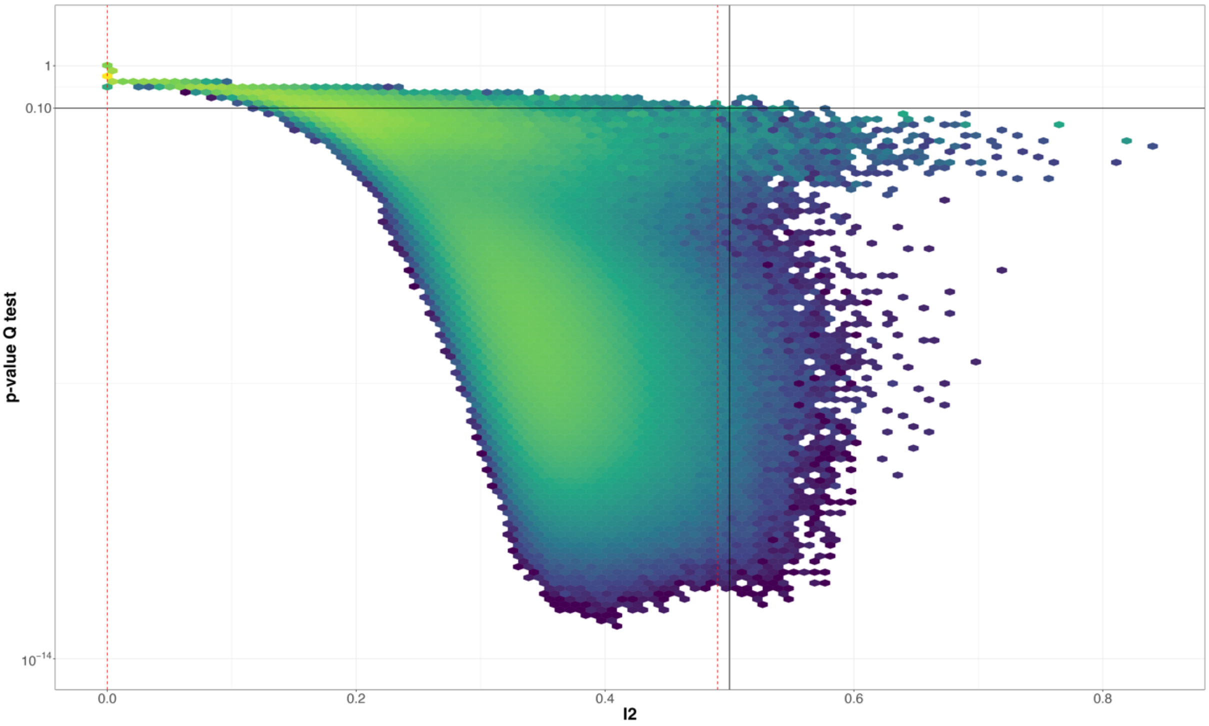


**e-figure 21:** *Vibration of effects for heterogeneity for NMAs. $I^2$ >0.5 indicates a high level of heterogeneity. The colors indicate the log densities of network meta-analyses (yellow: high, green: moderate, blue: low) for the outcome treatment response.*

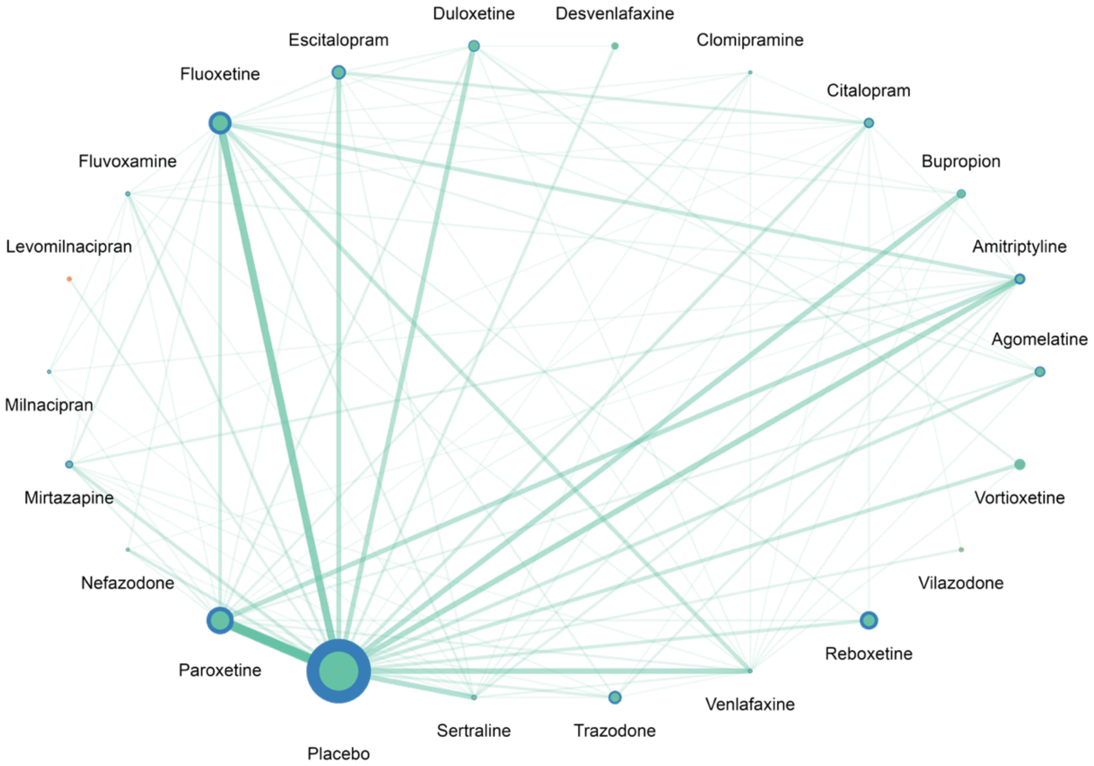


**e-figure 22:***Distribution of network geometries in the 4 121 590 NMAs on treatment discontinuation. The size of each dot indicates the number of patients allocated to the respective treatments. For each treatment, blue circles indicate the NMAs with the largest numbers of patients included, the green circles indicate the NMAs with the median number of patients included s, and the orange circles show the NMAs with the smallest number of patients included. The width of the lines is proportional to the number of trials comparing pairs of treatments in the complete NMA.*

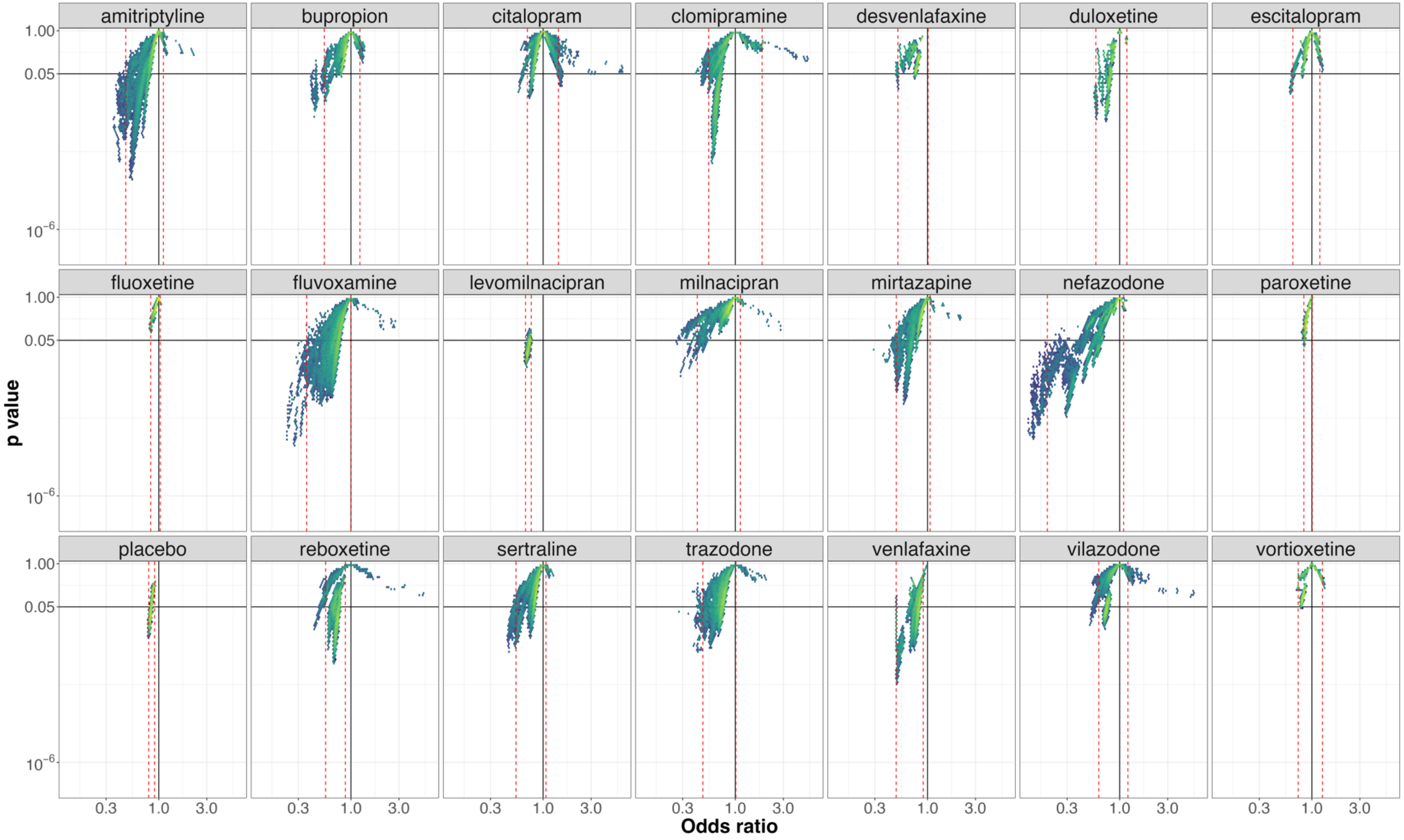


**e-figure 23**:*Vibration of effects for treatment discontinuation for the comparisons of agomelatine with the 20 remaining antidepressants and placebo (with the number of patients included in the most complete network for this comparison). An Odds ratio >1 favors agomelatine. The colors indicate the log densities of network meta-analyses (yellow: high, green: moderate, blue: low). Dotted red lines illustrate the 1st and 99th percentiles.*

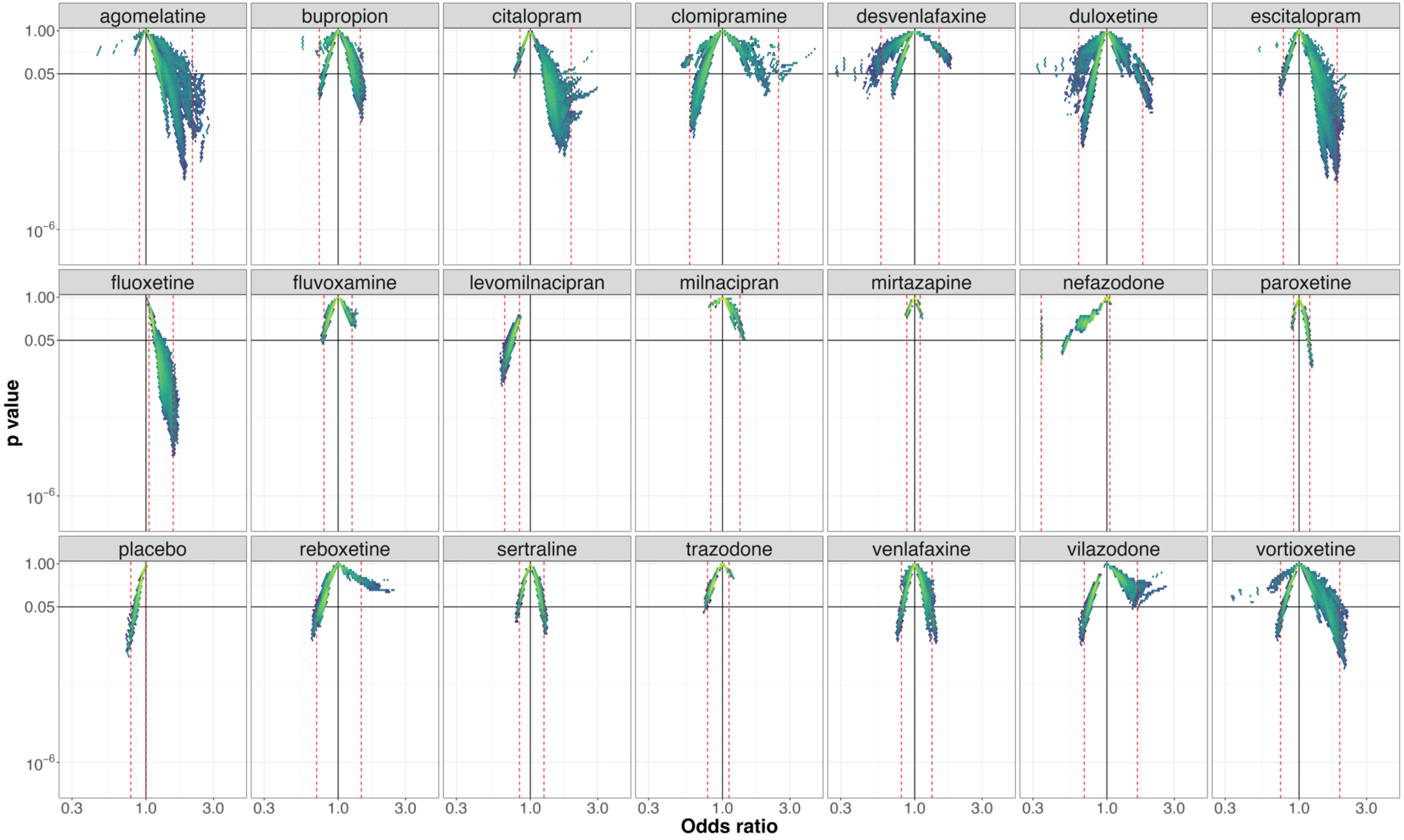


**e-figure 24**: *Vibration of effects for treatment discontinuation for the comparisons of amitriptyline with the 20 remaining antidepressants and placebo (with the number of patients included in the most complete network for this comparison). An Odds ratio >1 favors amitriptyline. The colors indicate the log densities of network meta-analyses (yellow: high, green: moderate, blue: low). Dotted red lines illustrate the 1st and 99th percentiles.*

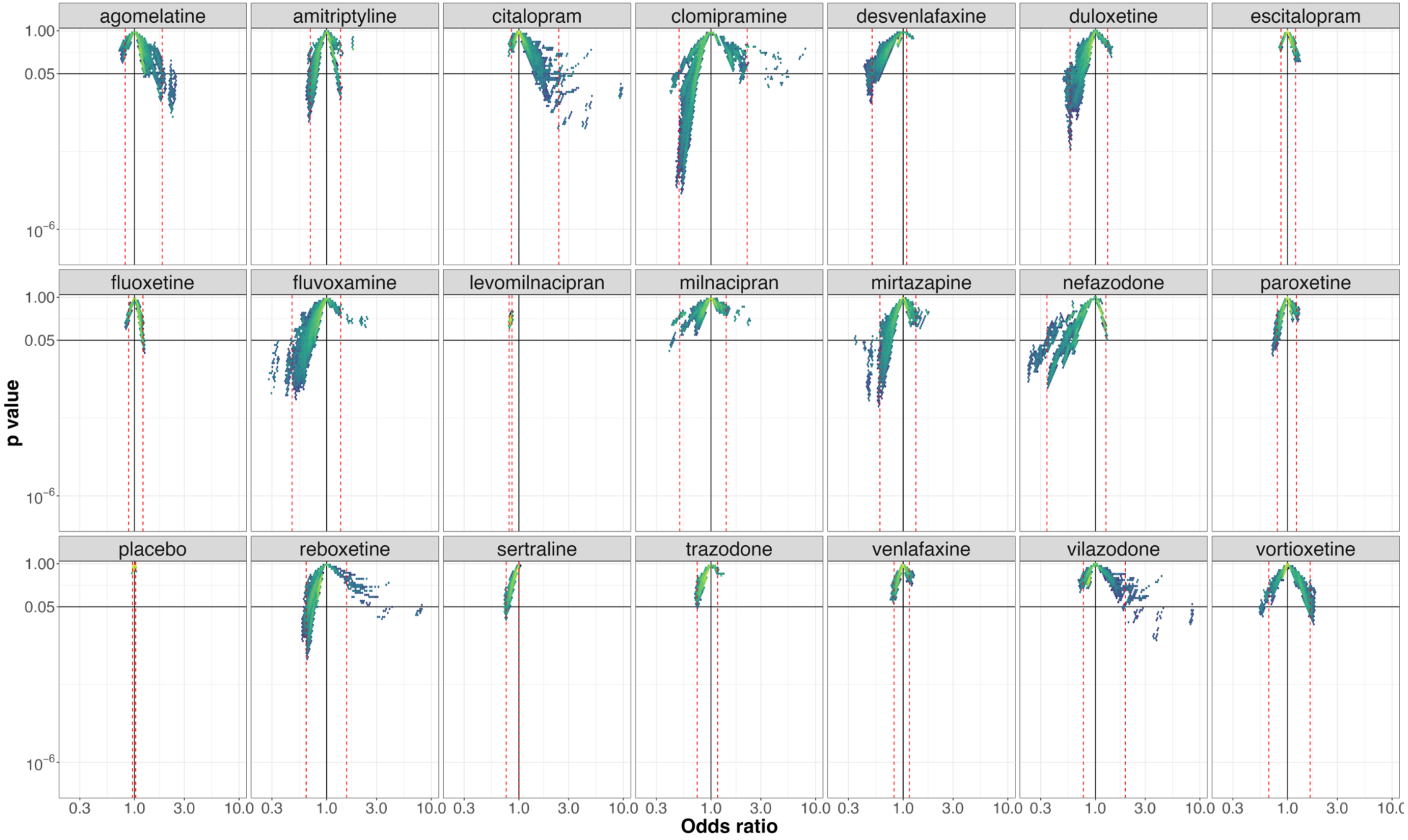


**e-figure 25**:*Vibration of effects for treatment discontinuation for the comparisons of bupropion with the 20 remaining antidepressants and placebo (with the number of patients included in the most complete network for this comparison). An Odds ratio >1 favors bupropion. The colors indicate the log densities of network meta-analyses (yellow: high, green: moderate, blue: low). Dotted red lines illustrate the 1st and 99th percentile.*

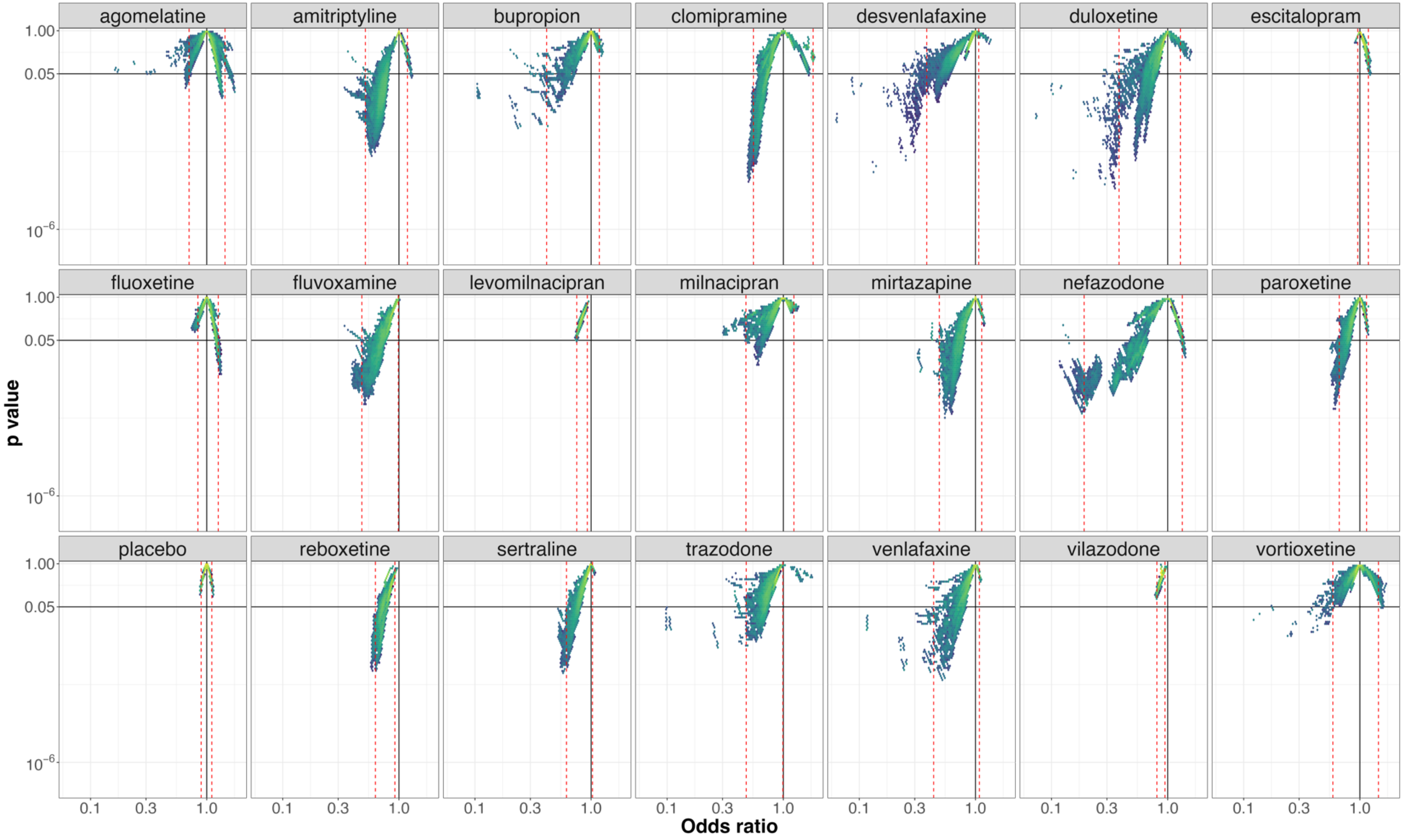


**e-figure 26**: *Vibration of effects for treatment discontinuation for the comparisons of citalopram with the 20 remaining antidepressants and placebo (with the number of patients included in the most complete network for this comparison). An Odds ratio >1 favors citalopram. The colors indicate the log densities of network meta-analyses (yellow: high, green: moderate, blue: low). Dotted red lines illustrate the 1st and 99th percentiles.*

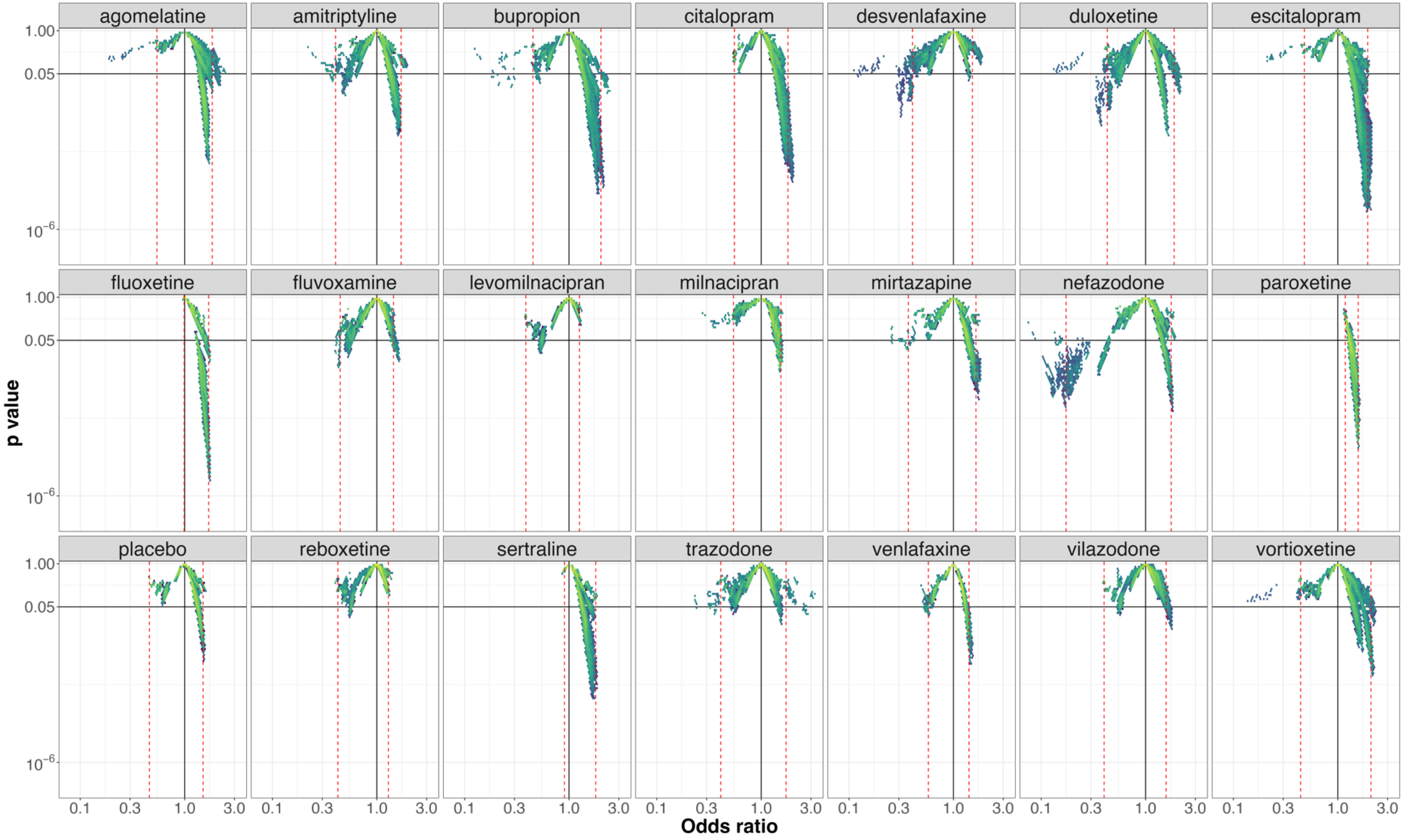


**e-figure 27**:*Vibration of effects for treatment discontinuation for the comparisons of clomipramine with the 20 remaining antidepressants and placebo (with the number of patients included in the most complete network for this comparison). An Odds ratio >1 favors clomipramine. The colors indicate the log densities of network meta-analyses (yellow: high, green: moderate, blue: low). Dotted red lines illustrate the 1st and 99th percentiles.*

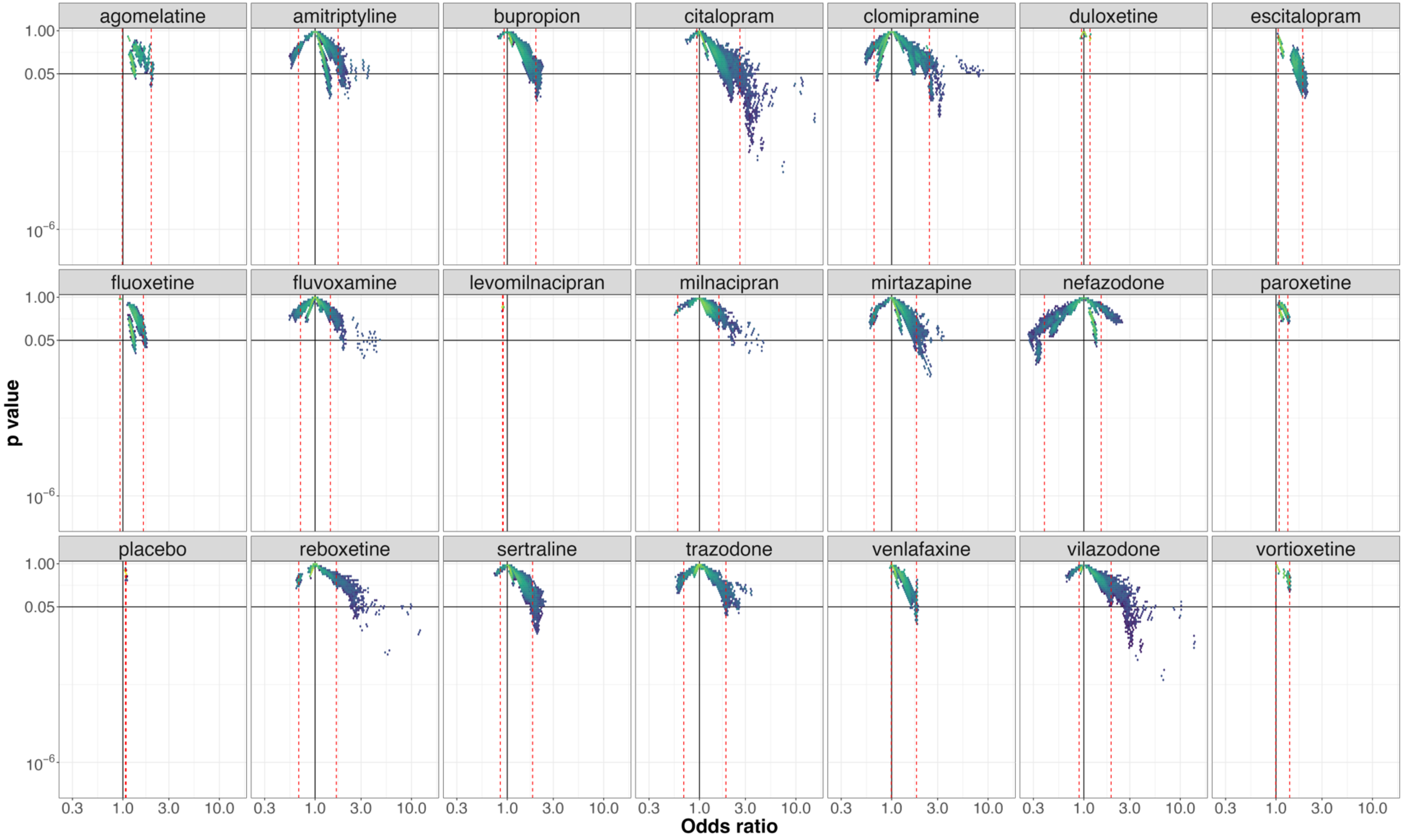


**e-figure 28**: *Vibration of effects for treatment discontinuation for the comparisons of desvenlafaxine with the 20 remaining antidepressants and placebo (with the number of patients included in the most complete network for this comparison). An Odds ratio >1 favors desvenlafaxine. The colors indicate the log densities of network meta-analyses (yellow: high, green: moderate, blue: low). Dotted red lines illustrate the 1st and 99th percentiles.*

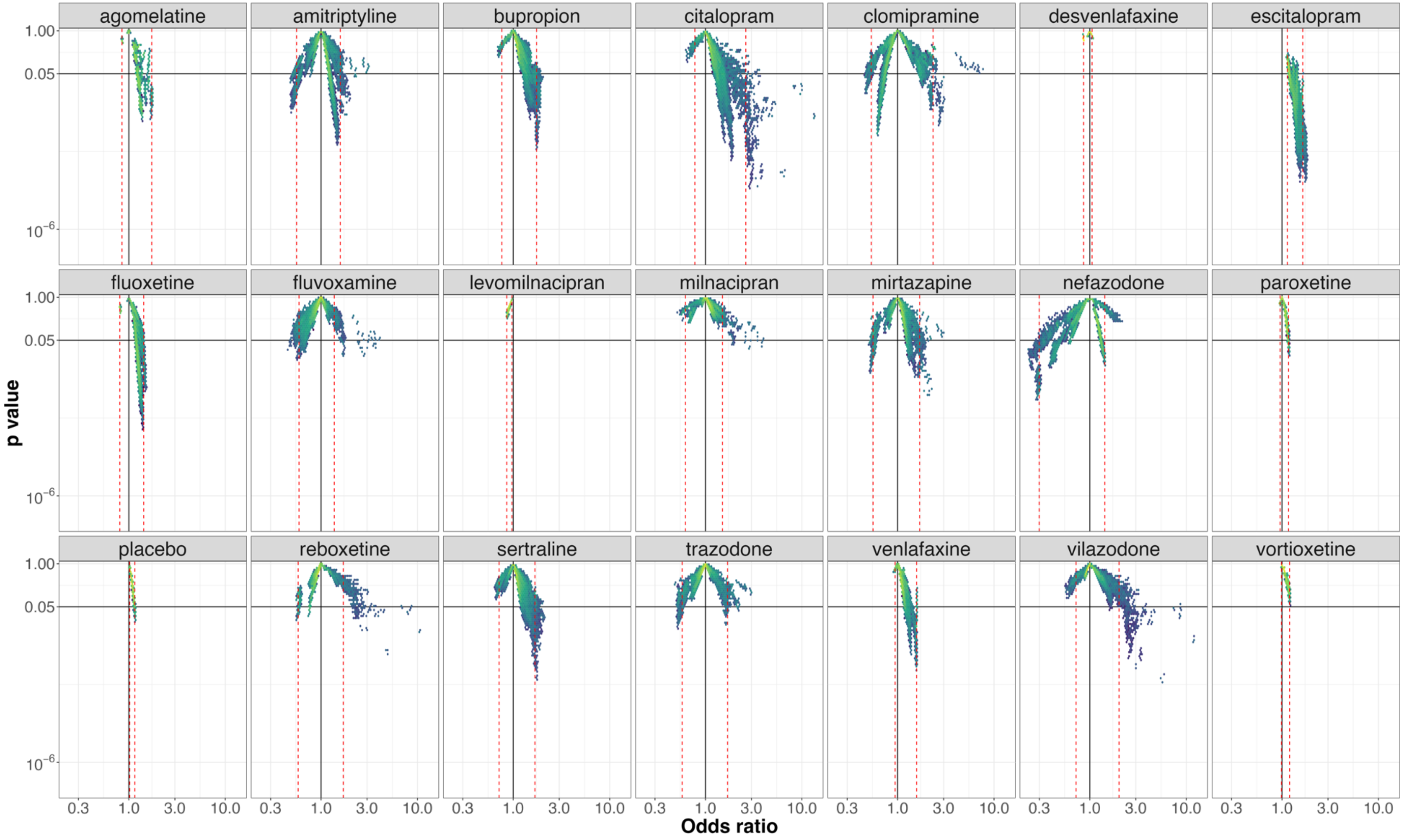


**e-figure 29**:*Vibration of effects for treatment discontinuation for the comparisons of duloxetine with the 20 remaining antidepressants and placebo (with the number of patients included in the most complete network for this comparison). An Odds ratio >1 favors duloxetine. The colors indicate the log densities of network meta-analyses (yellow: high, green: moderate, blue: low). Dotted red lines illustrate the 1st and 99th percentile.*

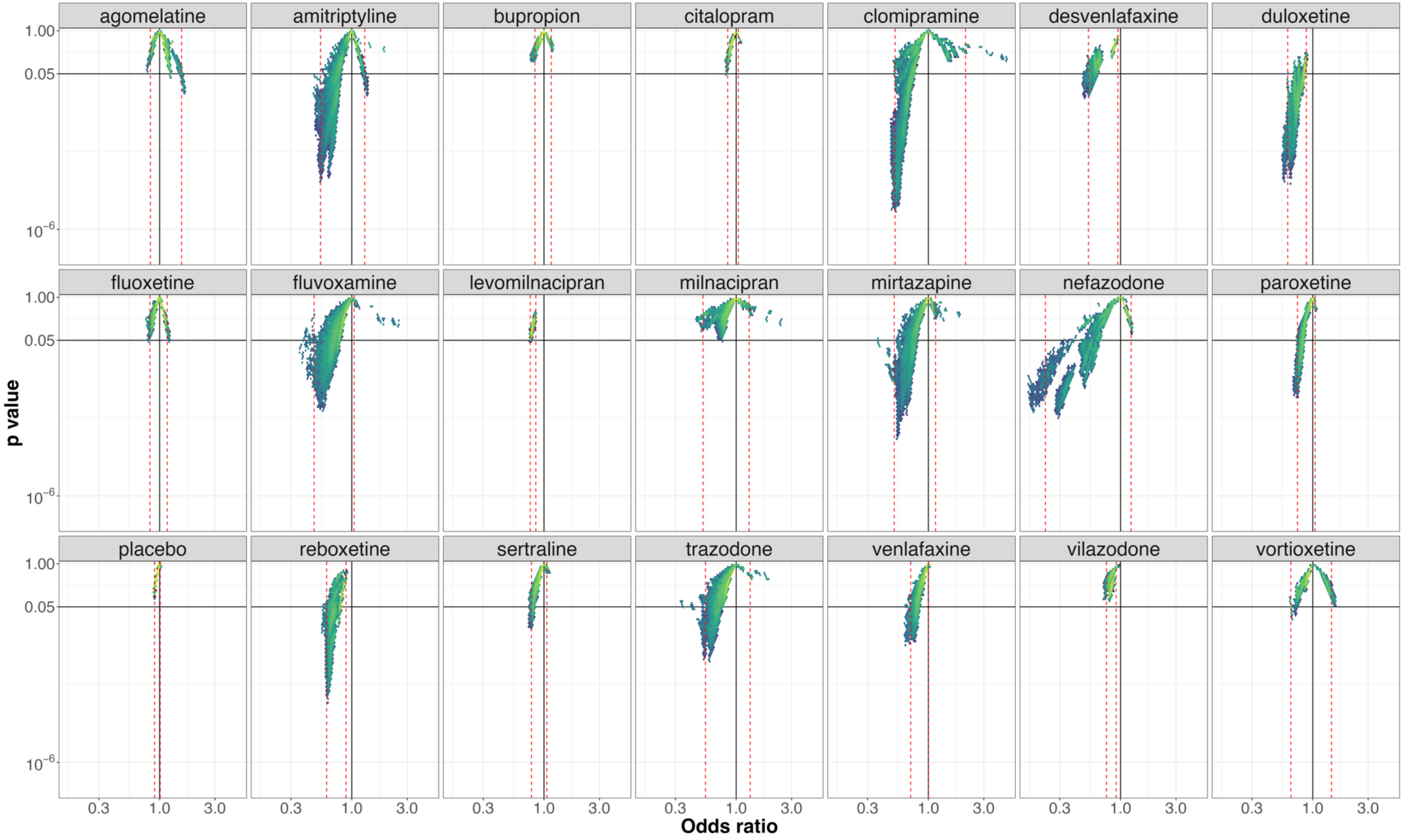


**e-figure 30**:*Vibration of effects for treatment discontinuation for the comparisons of escitalopram with the 20 remaining antidepressants and placebo (with the number of patients included in the most complete network for this comparison). An Odds ratio >1 favors escitalopram. The colors indicate the log densities of network meta-analyses (yellow: high, green: moderate, blue: low). Dotted red lines illustrate the* $1^{st}$ *and* $99^{th}$ *percentiles.*

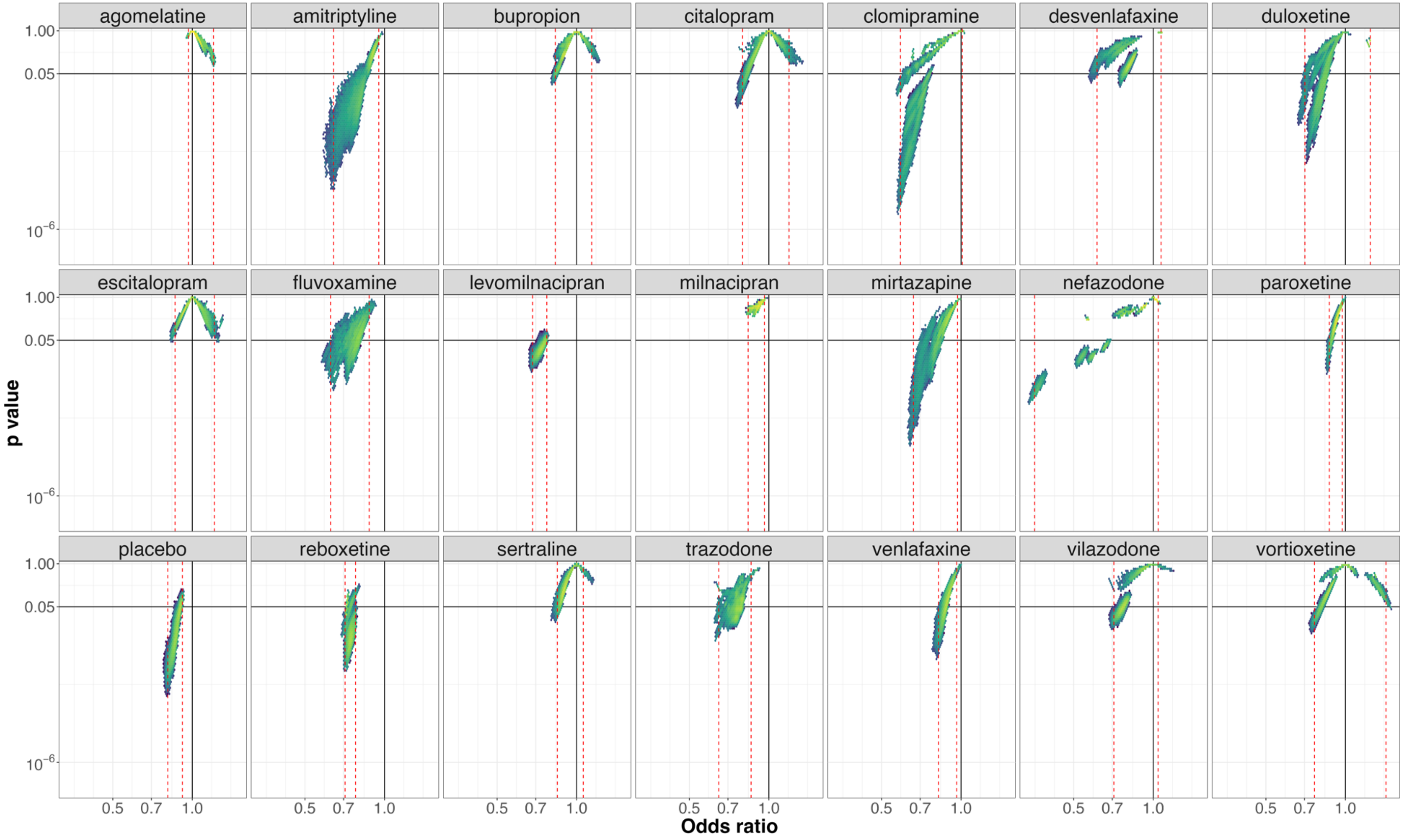


**e-figure 31**: *Vibration of effects for treatment discontinuation for the comparisons of fluoxetine with the 20 remaining antidepressants and placebo (with the number of patients included in the most complete network for this comparison). An Odds ratio >1 favors fluoxetine. The colors indicate the log densities of network meta-analyses (yellow: high, green: moderate, blue: low). Dotted red lines illustrate the 1st and 99th percentiles.*

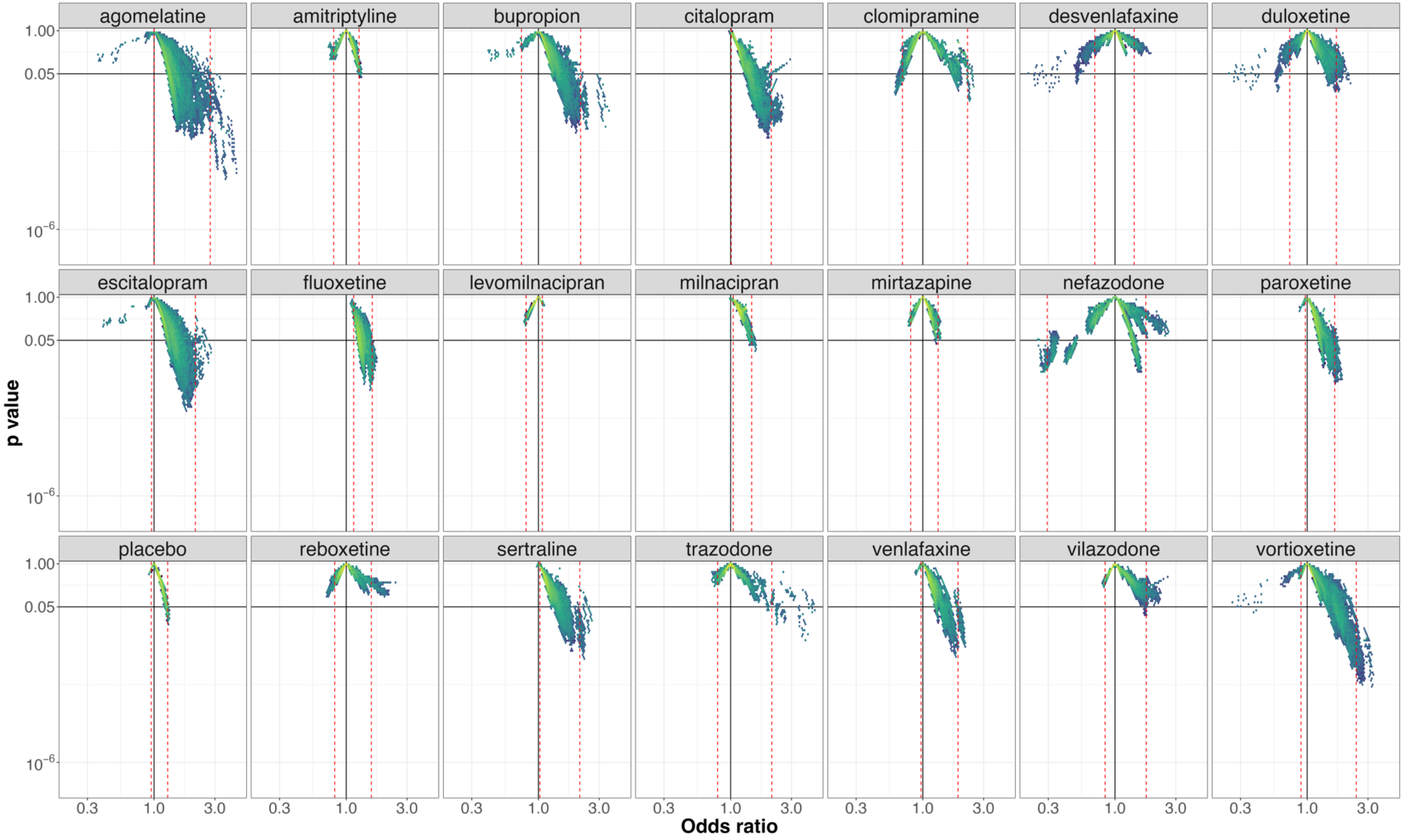


**e-figure 32**: *Vibration of effects for treatment discontinuation for the comparisons of fluvoxamine with the 20 remaining antidepressants and placebo (with the number of patients included in the most complete network for this comparison). An Odds ratio >1 favors fluvoxamine. The colors indicate the log densities of network meta-analyses (yellow: high, green: moderate, blue: low). Dotted red lines illustrate the 1st and 99th percentiles.*

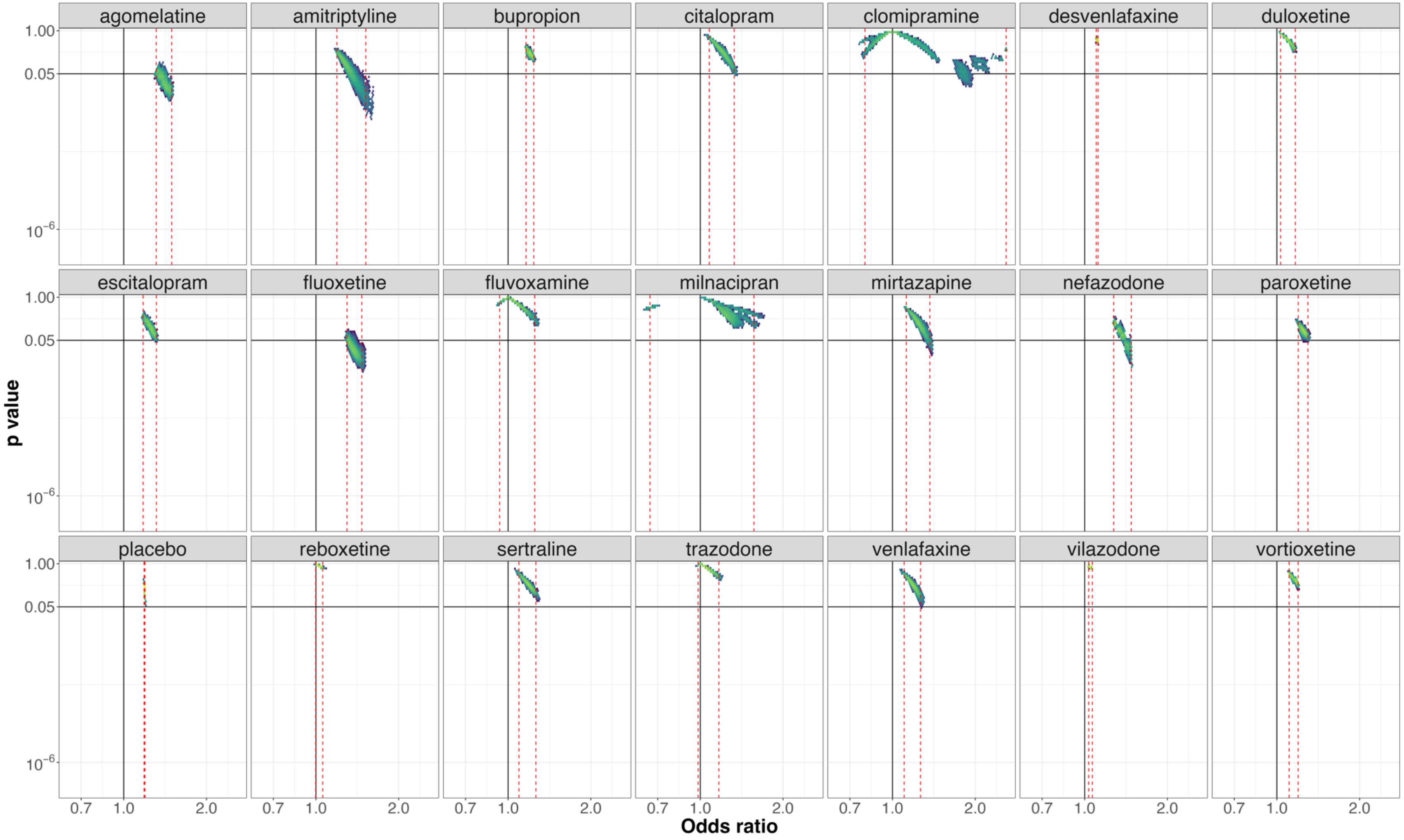


**e-figure 33**: *Vibration of effects for treatment discontinuation for the comparisons of levomilnacipran with the 20 remaining antidepressants and placebo (with the number of patients included in the most complete network for this comparison). An Odds ratio >1 favors levomilnacipran. The colors indicate the log densities of network meta-analyses (yellow: high, green: moderate, blue: low). Dotted red lines illustrate the 1st and 99th percentiles.*

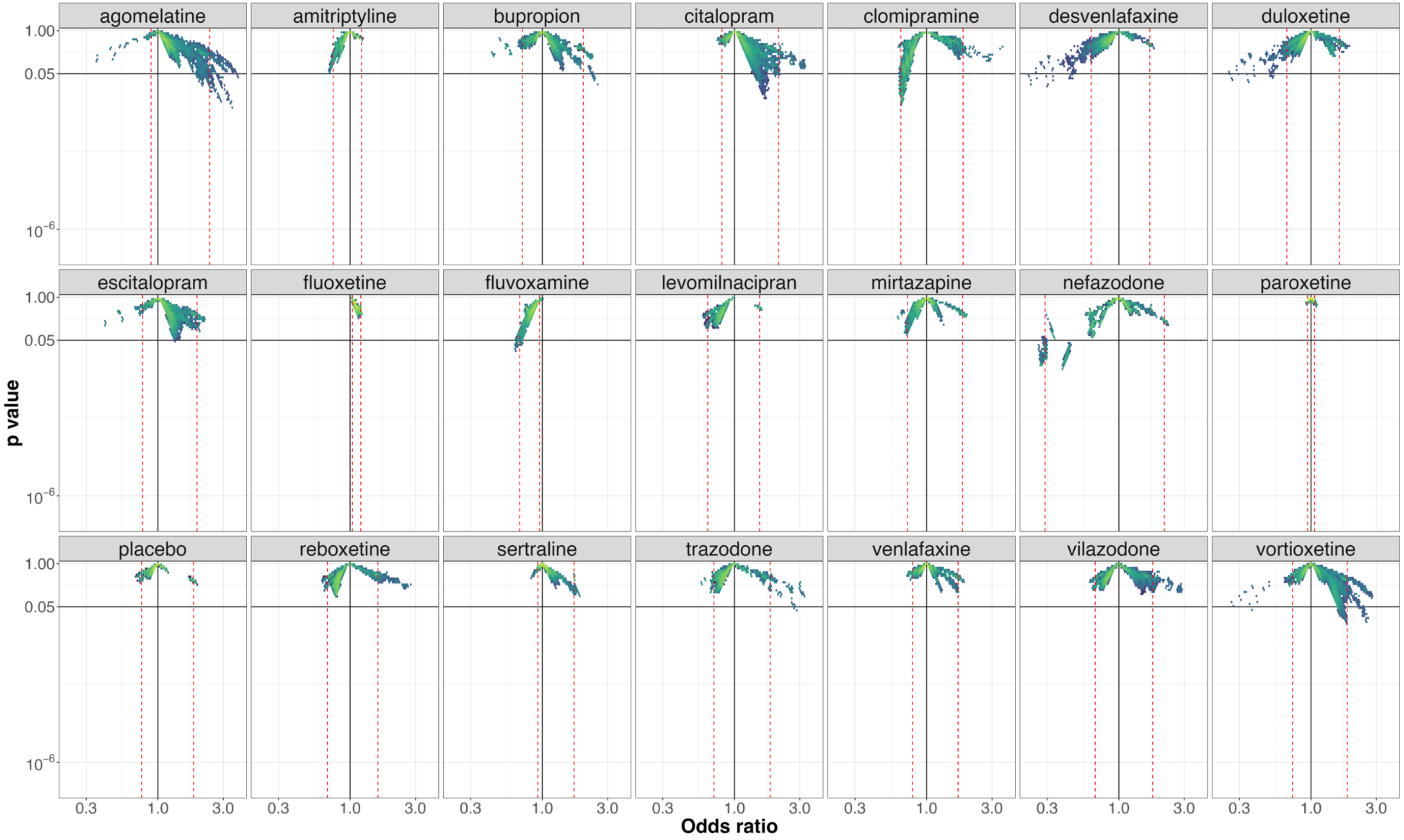


**e-figure 34**: *Vibration of effects for treatment discontinuation for the comparisons of milnacipran with the 20 remaining antidepressants and placebo (with the number of patients included in the most complete network for this comparison). An Odds ratio >1 favors milnacipran. The colors indicate the log densities of network meta-analyses (yellow: high, green: moderate, blue: low). Dotted red lines illustrate the 1st and 99th percentiles.*

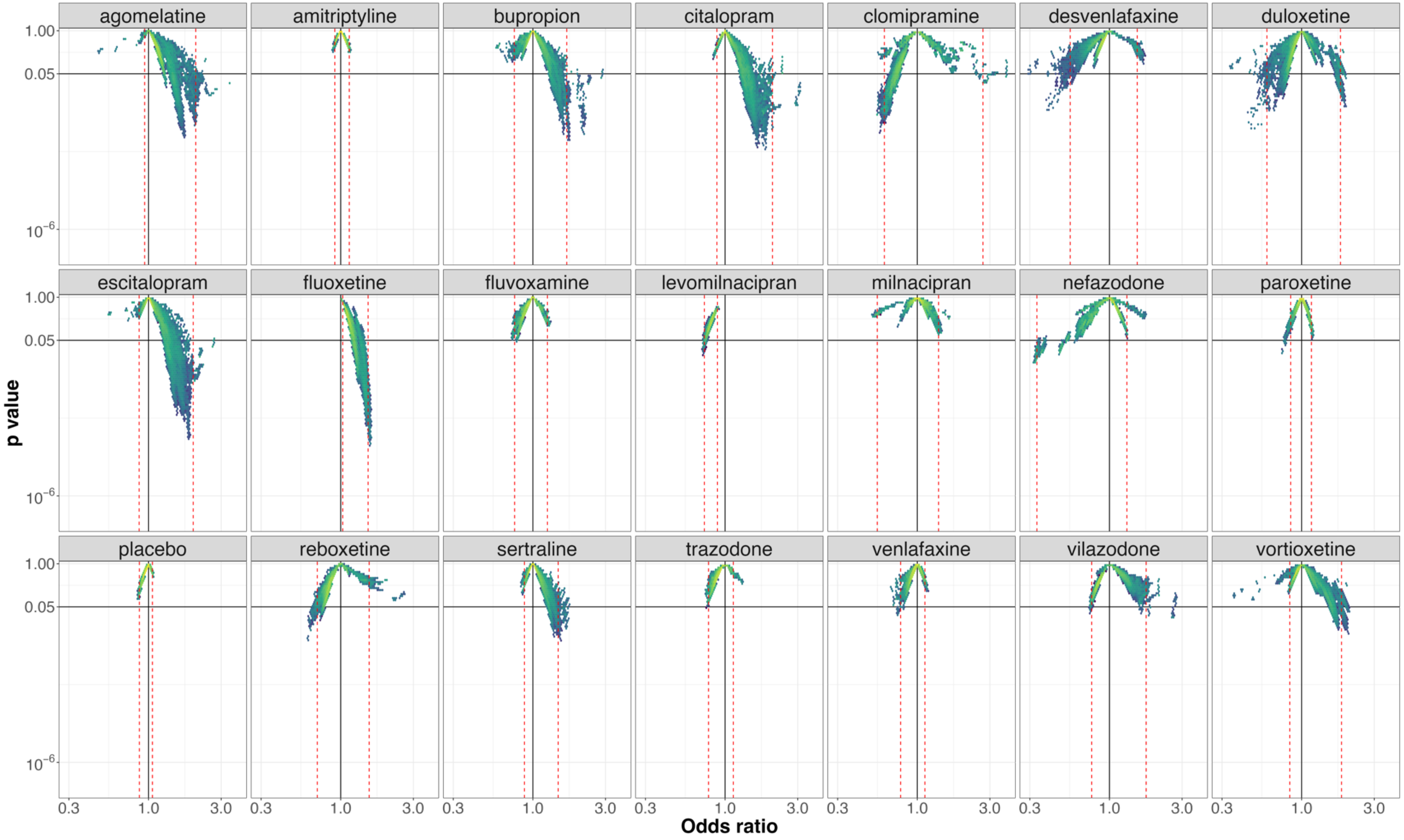


**e-figure 35**:*Vibration of effects for treatment discontinuation for the comparisons of mirtazapine with the 20 remaining antidepressants and placebo (with the number of patients included in the most complete network for this comparison). An Odds ratio >1 favors mirtazapine. The colors indicate the log densities of network meta-analyses (yellow: high, green: moderate, blue: low). Dotted red lines illustrate the 1st and 99th percentiles.*

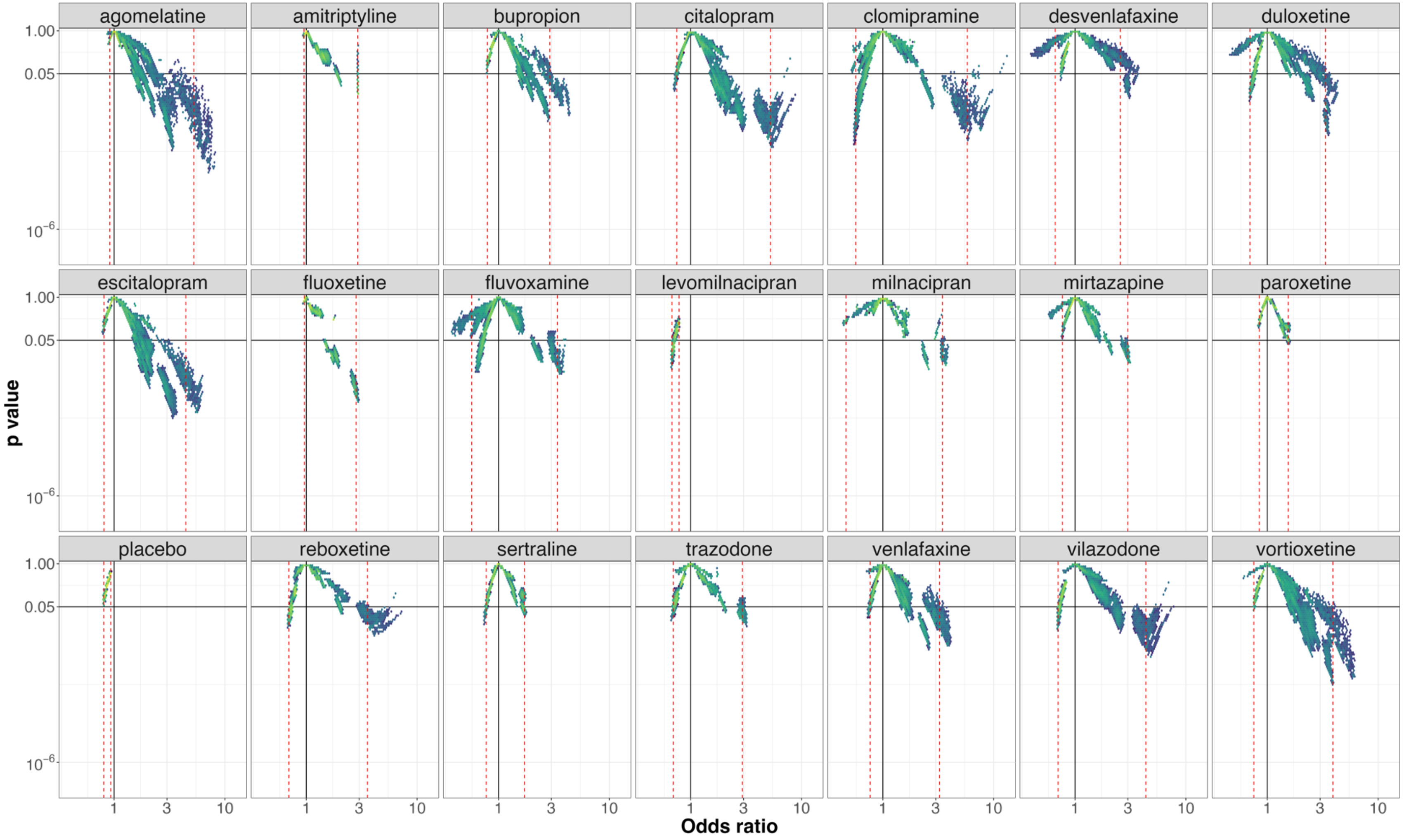


**e-figure 36**:*Vibration of effects for treatment discontinuation for the comparisons of nefazodone with the 20 remaining antidepressants and placebo (with the number of patients included in the most complete network for this comparison). An Odds ratio >1 favors nefazodone. The colors indicate the log densities of network meta-analyses (yellow: high, green: moderate, blue: low). Dotted red lines illustrate the 1st and 99th percentiles.*

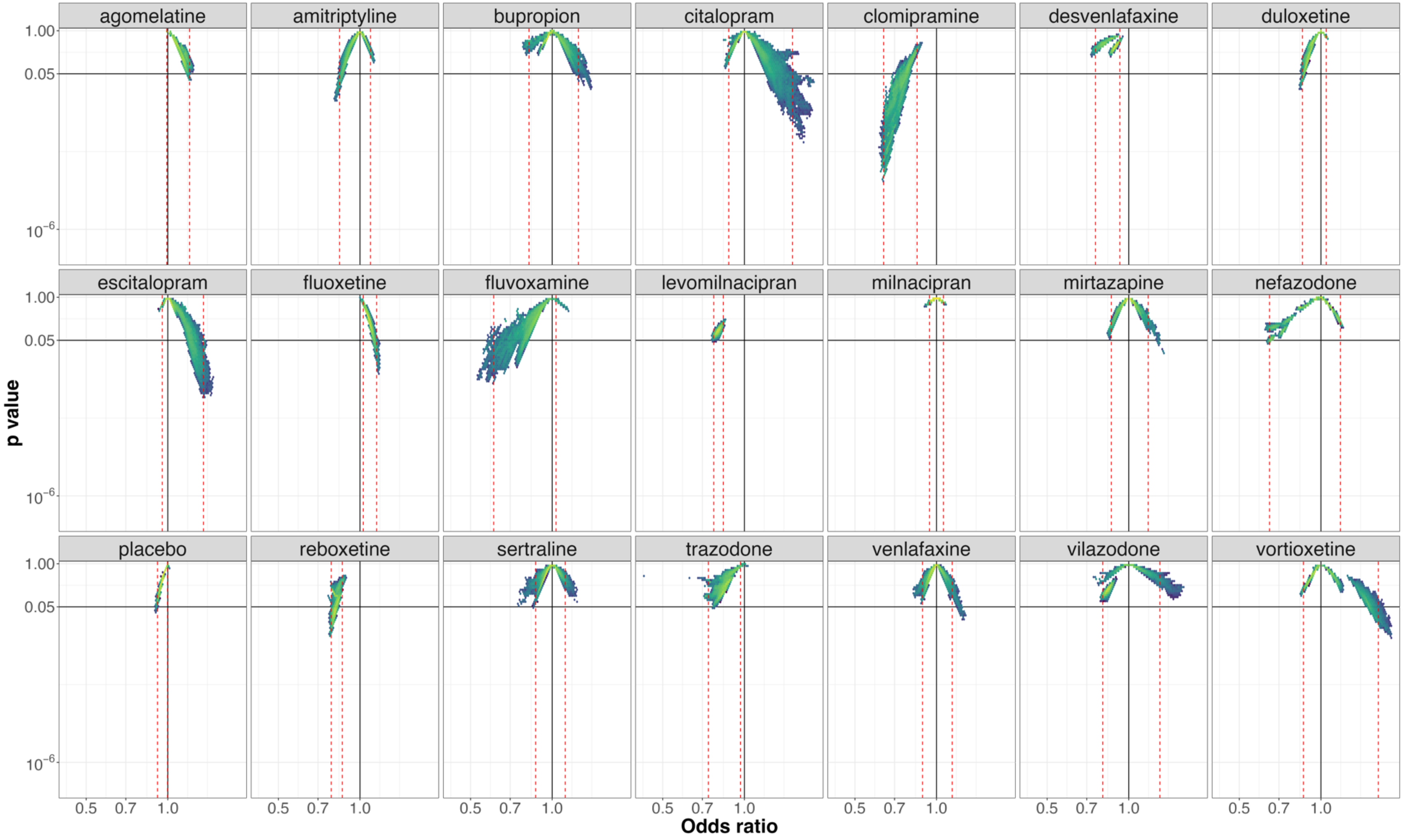


**e-figure 37**:*Vibration of effects for treatment discontinuation for the comparisons of paroxetine with the 20 remaining antidepressants and placebo (with the number of patients included in the most complete network for this comparison). An Odds ratio >1 favors paroxetine. The colors indicate the log densities of network meta-analyses (yellow: high, green: moderate, blue: low). Dotted red lines illustrate the 1$^{st}$ and 99$^{th}$ percentiles.*

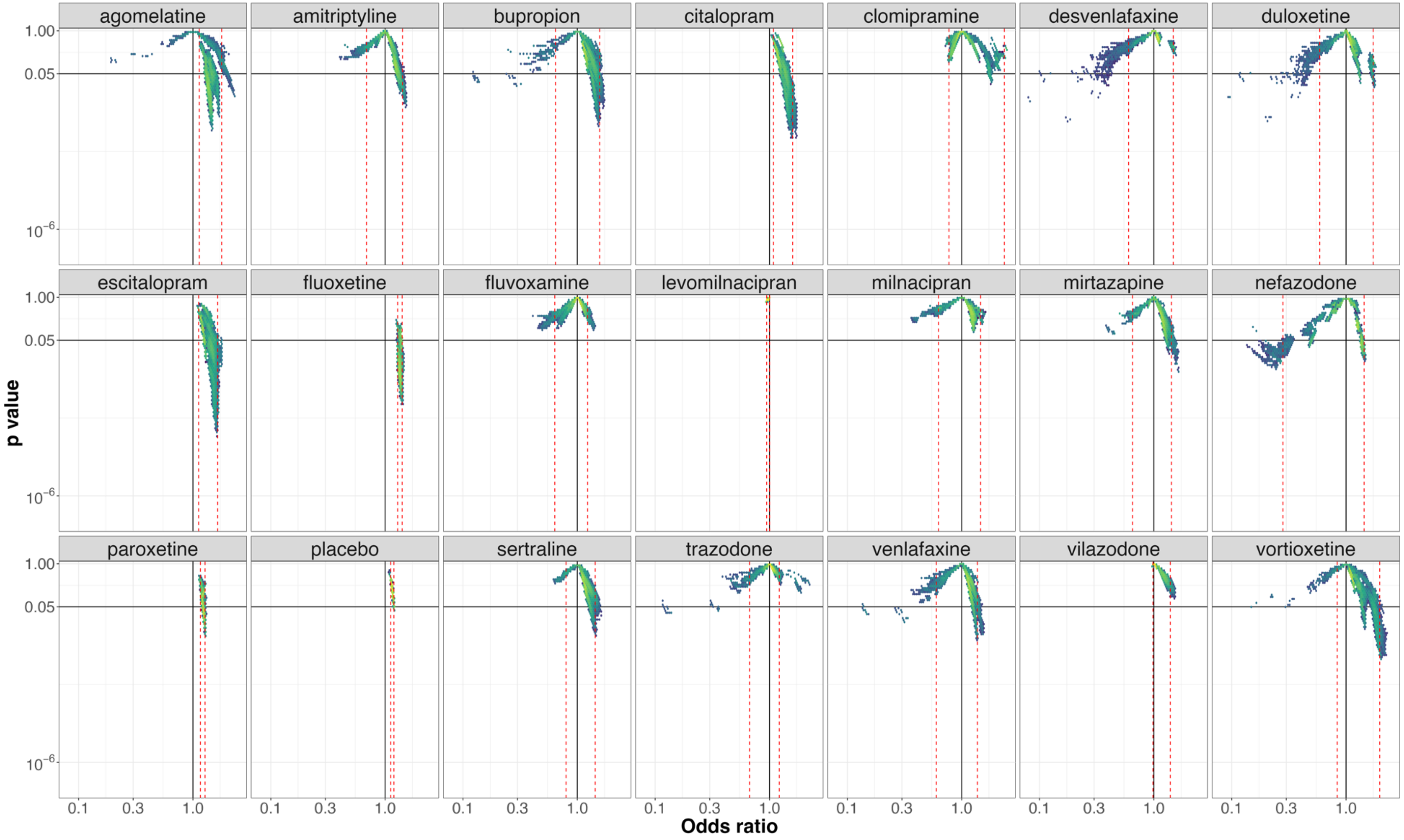


**e-figure 38**:*Vibration of effects for treatment discontinuation for the comparisons of reboxetine with the 20 remaining antidepressants and placebo (with the number of patients included in the most complete network for this comparison). An Odds ratio >1 favors reboxetine. The colors indicate the log densities of network meta-analyses (yellow: high, green: moderate, blue: low). Dotted red lines illustrate the 1st and 99th percentiles.*

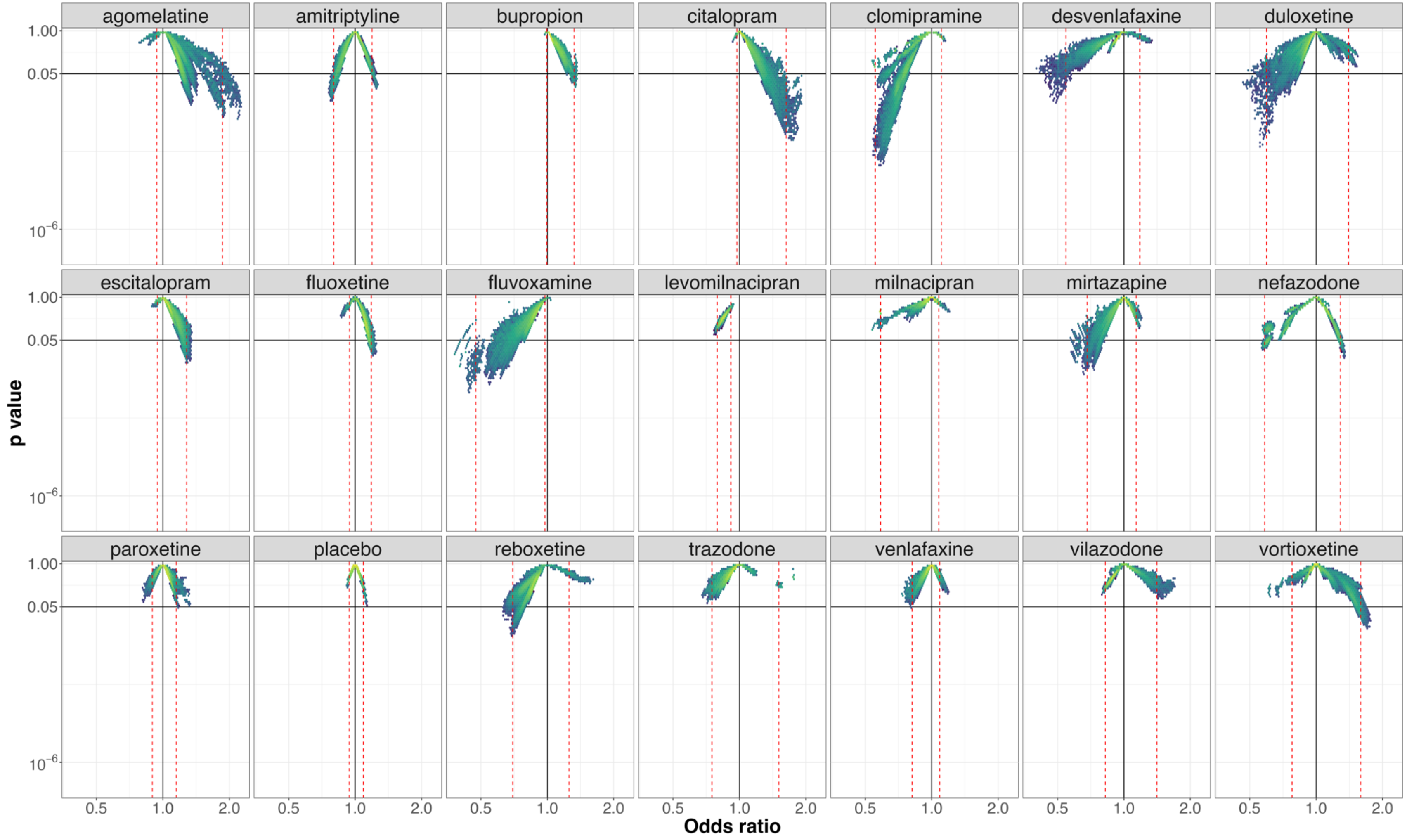


**e-figure 39**:*Vibration of effects for treatment discontinuation for the comparisons of sertraline with the 20 remaining antidepressants and placebo (with the number of patients included in the most complete network for this comparison). An Odds ratio >1 favors sertraline. The colors indicate the log densities of network meta-analyses (yellow: high, green: moderate, blue: low). Dotted red lines illustrate the 1st and 99th percentiles.*

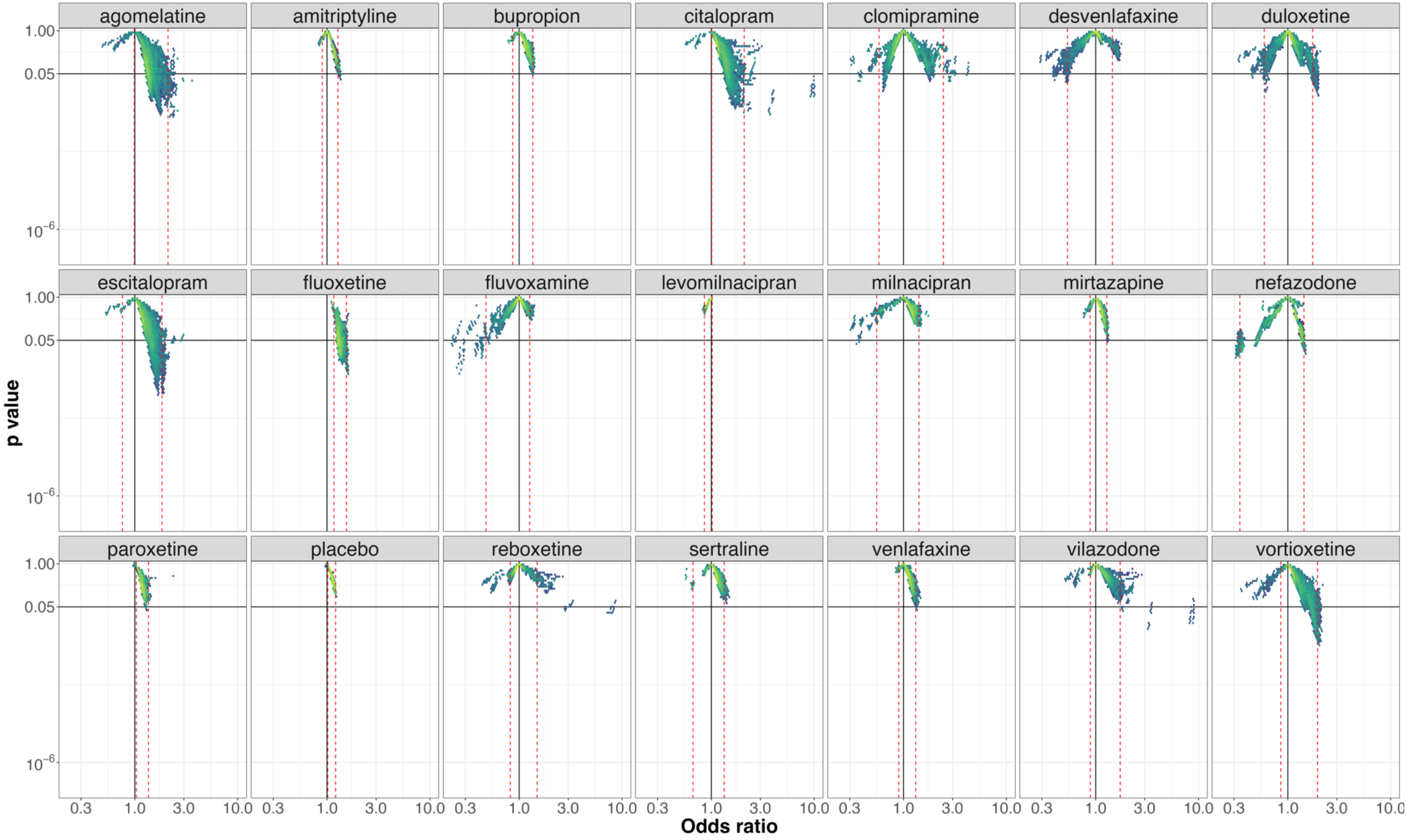


**e-figure 40**: *Vibration of effects for treatment discontinuation for the comparisons of trazodone with the 20 remaining antidepressants and placebo (with the number of patients included in the most complete network for this comparison). An Odds ratio >1 favors trazodone. The colors indicate the log densities of network meta-analyses (yellow: high, green: moderate, blue: low). Dotted red lines illustrate the 1st and 99th percentiles.*

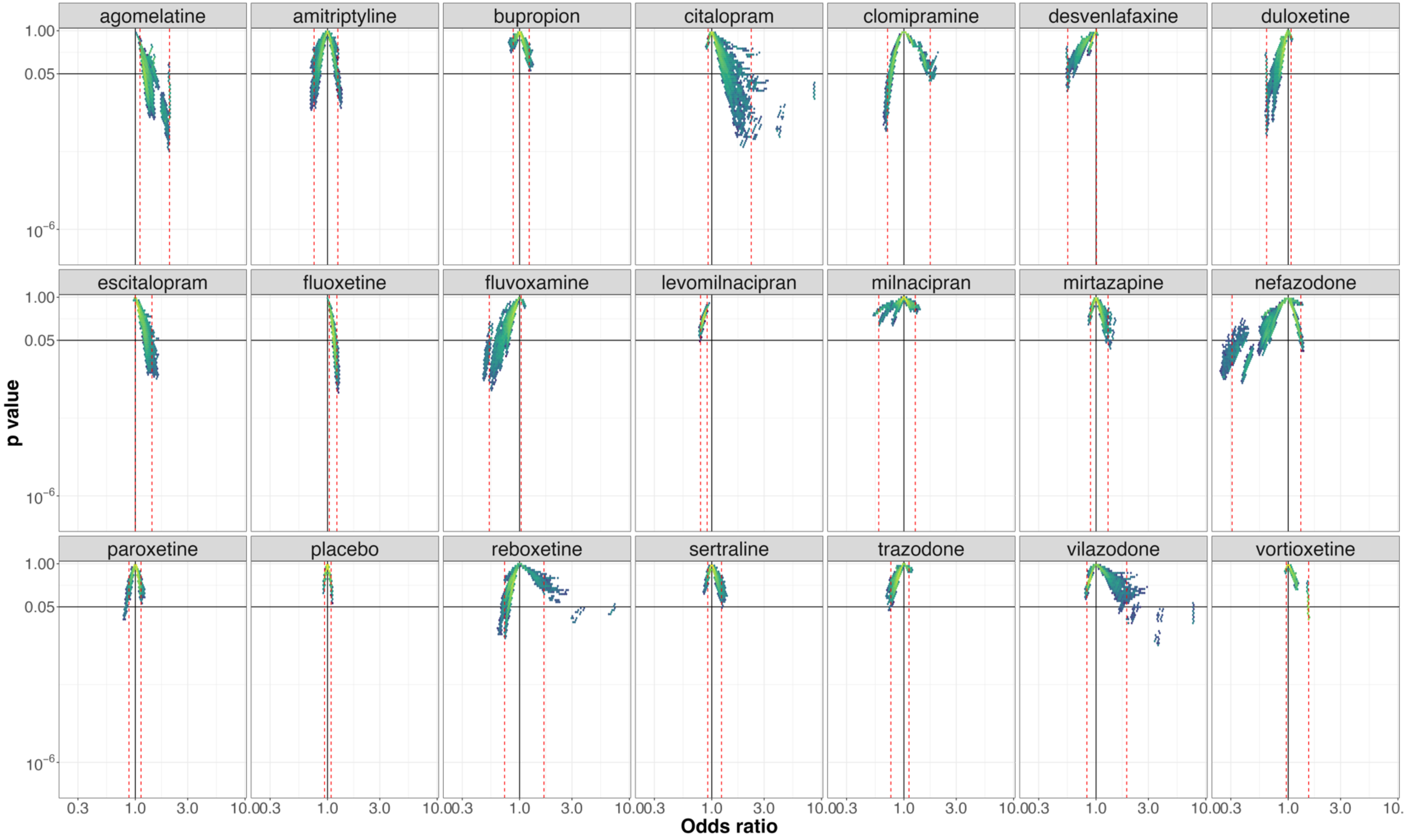


**e-figure 41**:*Vibration of effects for treatment discontinuation for the comparisons of venlafaxine with the 20 remaining antidepressants and placebo (with the number of patients included in the most complete network for this comparison). An Odds ratio >1 favors venlafaxine. The colors indicate the log densities of network meta-analyses (yellow: high, green: moderate, blue: low). Dotted red lines illustrate the 1st and 99th percentiles.*

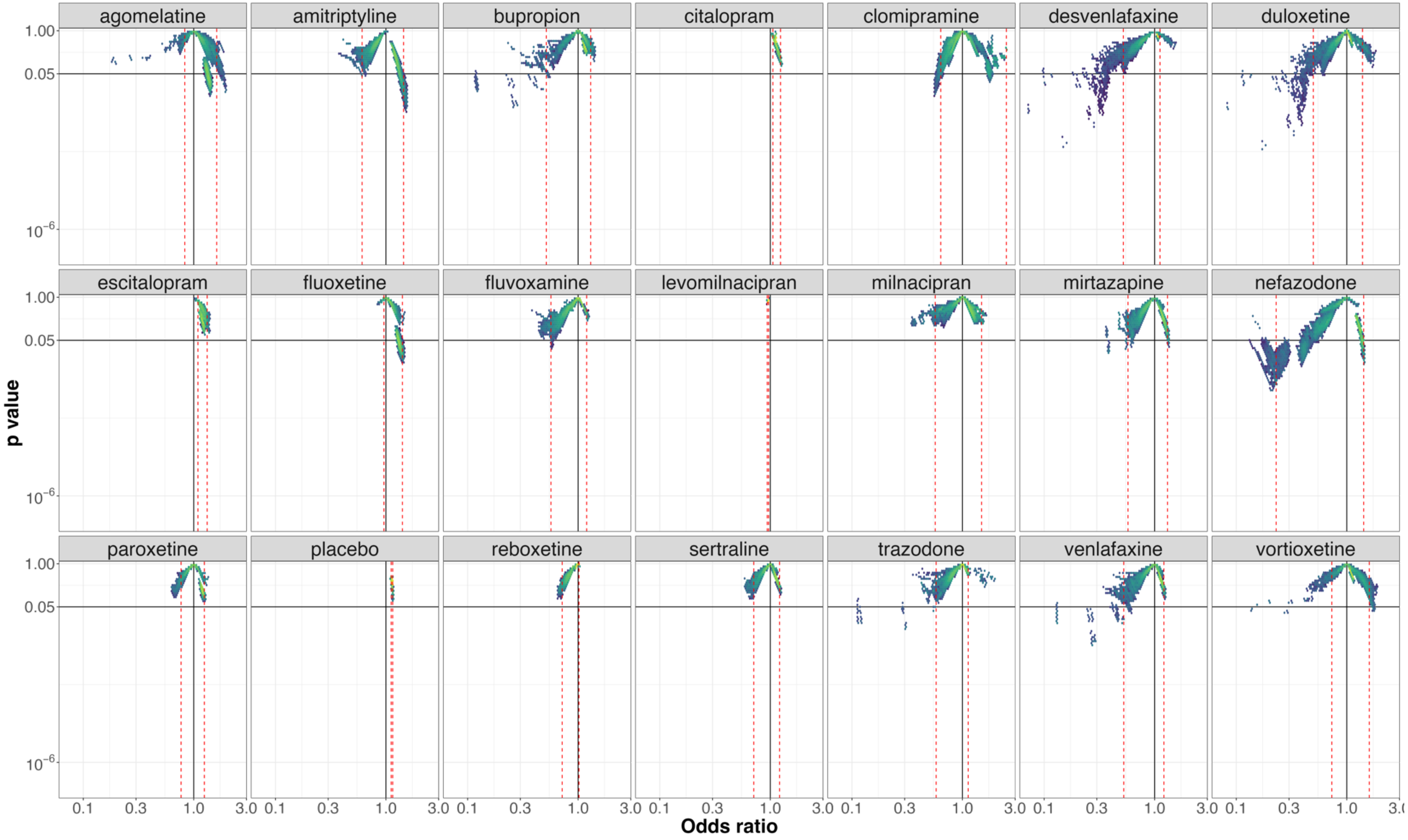


**e-figure 42**:*Vibration of effects for treatment discontinuation for the comparisons of vilazodone with the 20 remaining antidepressants and placebo (with the number of patients included in the most complete network for this comparison). An Odds ratio >1 favors vilazodone. The colors indicate the log densities of network meta-analyses (yellow: high, green: moderate, blue: low). Dotted red lines illustrate the 1st and 99th percentiles.*

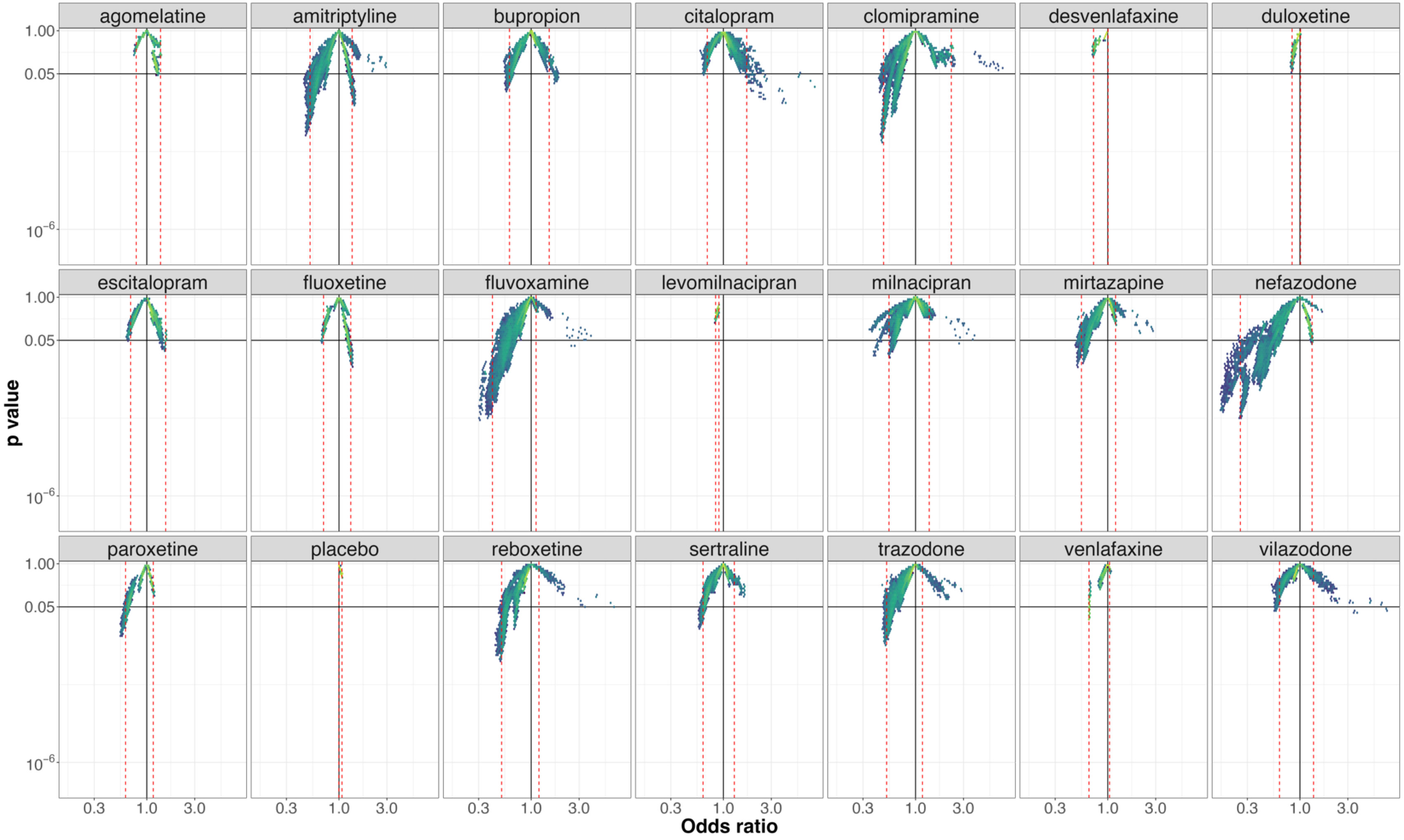


**e-figure 43**:*Vibration of effects for treatment discontinuation for the comparisons of vortioxetine with the 20 remaining antidepressants and placebo (with the number of patients included in the most complete network for this comparison). An Odds ratio >1 favors vortioxetine. The colors indicate the log densities of network meta-analyses (yellow: high, green: moderate, blue: low). Dotted red lines illustrate the 1st and 99th percentiles.*

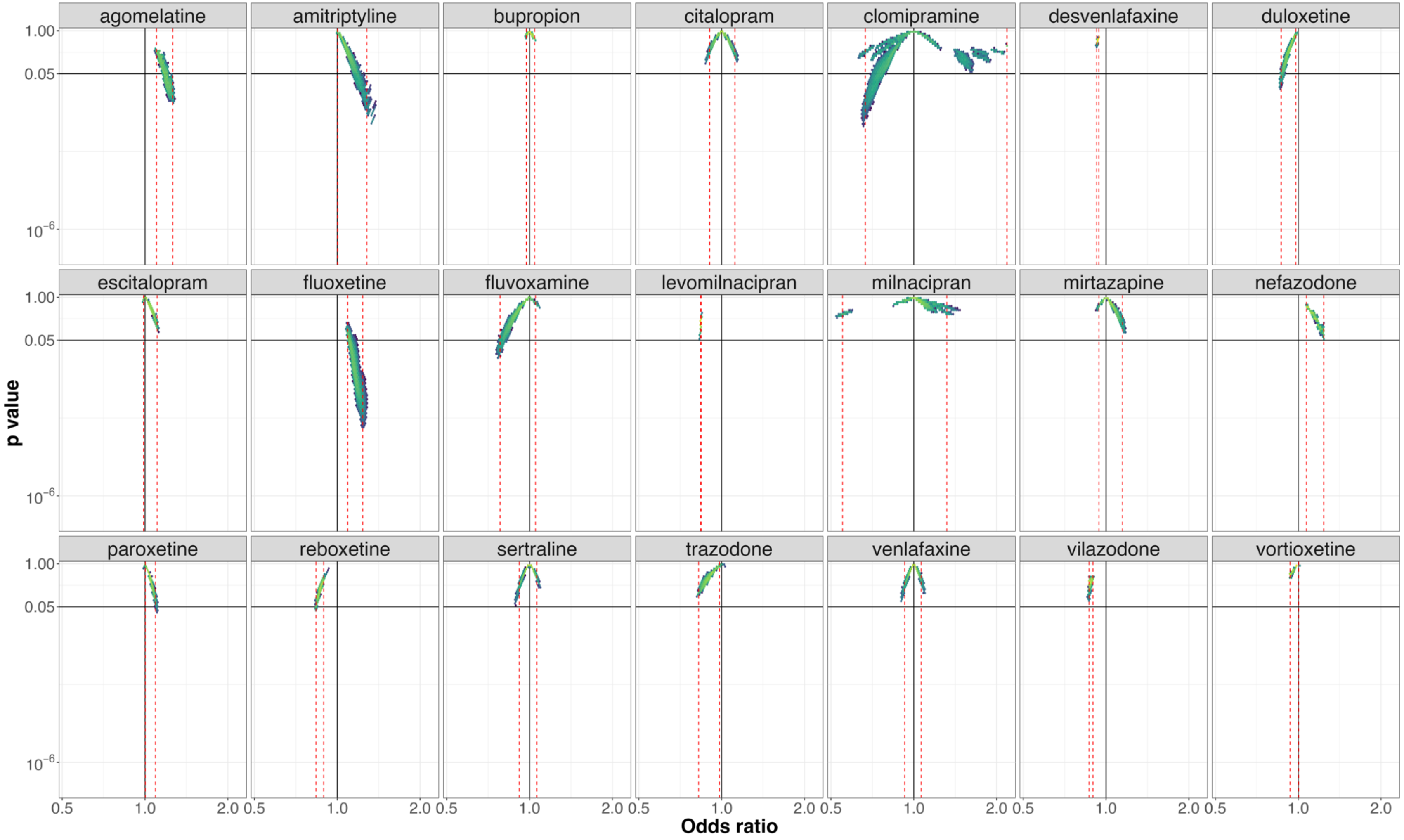


**e-figure 44**: *Vibration of effects for treatment discontinuation for the comparisons of placebo with the 21 antidepressants (with the number of patients included in the most complete network for this comparison). An Odds ratio >1 favors placebo. The colors indicate the log densities of network meta-analyses (yellow: high, green: moderate, blue: low). Dotted red lines illustrate the 1st and 99th percentiles.*

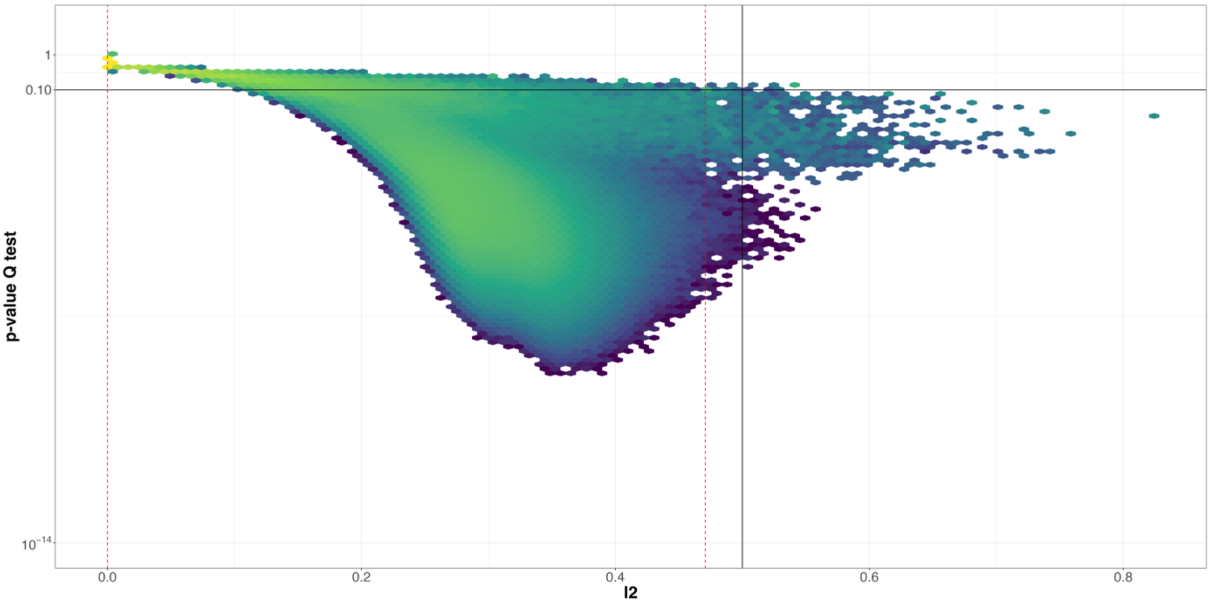


**e-figure 45:** *Vibration of effects for heterogeneity for NMAs. $I^2$ >0.5 indicates a high level of heterogeneity The colors indicate the log densities of network meta-analyses (yellow: high, green: moderate, blue: low) for the outcome treatment discontinuation.*
